# Characterizing the Uncertainty, Misclassification and Inconsistency of Polygenic Prediction

**DOI:** 10.64898/2026.05.11.26352897

**Authors:** Yingzhe Zhang, Rui Zhang, Tian Ge

## Abstract

Polygenic risk scores (PRSs) hold promise for precision medicine, yet their clinical translation is hindered by substantial uncertainty in individual risk estimates and often limited agreement in risk stratification across multiple PRSs for the same disease. We develop a unified inferential framework to calibrate PRS point estimates and uncertainties for both quantitative traits and binary phenotypes, and to characterize how PRS accuracy, uncertainty, pairwise correlation jointly determine misclassification and classification inconsistency. We show, both theoretically and empirically, that individual- and population-level misclassification and inconsistency rates are highly predictable in independent datasets. We further evaluate PRS integration and uncertainty-aware probabilistic thresholding strategies that reduce misclassification and improve concordance in risk stratification. Together, these results demonstrate that instability in PRS-based classification is a predictable statistical consequence of uncertainty and establish a principled foundation for incorporating uncertainty into PRS-based risk interpretation, communication, and clinical decision-making.

## Introduction

Polygenic risk scores (PRSs), which aggregate the effects of many common genetic variants across the genome to quantify the genetic predisposition of an individual to complex traits and diseases, hold promise for advancing precision medicine. Ongoing global efforts to expand the sample size and ancestral diversity of genome-wide association studies (GWASs)^1–9^, combined with continued statistical and computational innovation^10–18^, have improved the predictive performance of PRSs both within and across populations. Early translational studies have demonstrated the potential clinical utility of PRSs and informed best practices for their implementation and risk communication in healthcare settings^19–25^, while also highlighting key challenges that remain to be addressed^26–30^.

A central issue concerns the uncertainty inherent in PRSs^31,32^. Current practice typically stratifies individuals by thresholding PRS point estimates, largely ignoring the potentially large and individual-specific uncertainties associated with these estimates. Such uncertainty can lead to misclassification of high-risk individuals and instability in the selected high-risk group. These uncertainties further propagate to affect agreement among multiple PRSs for the same disease, derived from different GWASs or constructed using different methods^33–35^. As demonstrated in recent empirical studies, PRSs with similar population-level performance can yield highly variable individual-level risk estimates and identify divergent subsets of individuals as high risk^33–35^. These discrepancies complicate clinical interpretation and decision-making and may undermine the utility of PRSs.

To address these issues, recent work has developed parametric or nonparametric methods to quantify uncertainty in PRS-based predictions, for example, through the construction of confidence intervals on the phenotype scale^36–38^. However, a systematic framework for calibrating uncertainty on the genetic scale – and for understanding how this uncertainty, together with other factors, shapes misclassification rates of individual PRSs and classification inconsistency among multiple PRSs – remains lacking.

In this work, we ask: (1) How to properly calibrate PRS point estimates and their uncertainties for both quantitative traits and binary phenotypes? (2) How uncertainty in PRS estimates influences the probability of misclassifying individuals at high genetic risk? (3) What factors determine the degree of inconsistency in risk stratification across multiple PRSs for the same phenotype? (4) How to accurately estimate misclassification and inconsistency rates in independent datasets? (5) How to reduce misclassification and improve agreement across PRSs? We develop theoretical results and conduct simulations and empirical analyses in the *All of Us* Research Program^3^ to answer these questions.

## Results

### Polygenic model

We assume a standard linear model relating genotype to phenotype for individual 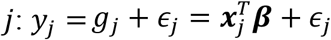, where *y*_*j*_ is standardized to have zero mean and unit variance; ***x***_*j*_ is an *M* × 1 vector of standardized genotypes; ***β*** is an *M* × 1 vector of causal genetic effects; 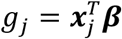 is the true genetic value; and *ϵ*_*j*_ is residual noise. Under a Bayesian polygenic prediction framework, a prior distribution is specified for ***β***, and the posterior mean given GWAS summary statistics ***D*** is estimated as 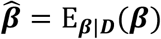. The PRS for an unseen individual *i* is then calculated as 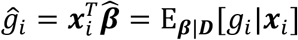. In practice, a threshold *t* may be applied to 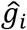 to identify individuals at elevated genetic risk.

The estimated effects 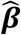 carry uncertainty that depends on factors such as the genetic architecture of the phenotype and the sample size of the training GWAS^31^. This uncertainty propagates to individual-level PRS estimates 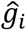. Following Ding et al.^31^, inferential uncertainty in 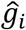 can be quantified by sampling ***β*** from its posterior distribution and computing the posterior variance of 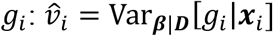. Assuming the posterior distribution of *g*_*i*_ is approximately normal, an individual-specific (1 − *α*)-level confidence interval (CI) is 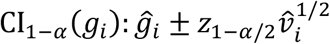, where *z*_1−*α*/2_ is the (1 − *α*/2)-quantile of the standard normal distribution.

### Overview of simulation design

Throughout this paper, we illustrate our theoretical results using extensive simulations. Individual-level genotypes for HapMap3^39^ variants with minor allele frequency (MAF) >1% were generated using HAPGEN2^40^, with the 1000 Genomes Project (1KGP)^41^ phase 3 European samples (*N* = 503) as the reference panel. In the primary simulation setting, we randomly designated 1% of variants as causal (polygenicity *π* = 1%) and drew their per-allele effect sizes from a normal distribution with homogeneous variance across the genome. These genetic effects jointly explained 50% of phenotypic variation (SNP heritability *h*^*2*^ = 50%). Training GWAS sample sizes ranged from 50,000 to 250,000. To investigate classification inconsistency across PRSs (using a top 10% high-risk threshold) and the integration of multiple PRSs, we considered two study designs: (i) Overlapping setting, comprising five nested training datasets of increasing size (50K, 100K, 150K, 200K, 250K), where each smaller dataset was a subset of the next larger dataset; and (ii) Independent setting, comprising five independent training datasets of size 50K each.

We additionally conducted secondary simulations, in which we varied polygenicity (*π* = 0.1% and 10%), SNP heritability (*h*^*2*^ = 20% and 80%), and percentile cutoffs (top 5% and top 2%). We also simulated binary phenotypes under the liability threshold model with prevalences of 10% and 20%.

In all analyses, the training datasets were used to generate GWAS summary statistics, which were input to PRS-CS-auto^12^ to obtain posterior samples of SNP effects. A separate validation dataset with individual-level genotypes and phenotypes was used for PRS calibration and for estimating quantities (e.g., PRS accuracy and pairwise correlations) required for theoretical predictions of misclassification and classification inconsistency. An independent testing dataset was used to evaluate calibration, misclassification, inconsistency, and PRS integration. Training, validation, and testing datasets were mutually non-overlapping.

### Slope and variance calibration

Slope calibration evaluates whether the regression slope of the true genetic value *g*_*i*_ on the predicted risk score 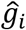 equals 1 (Figure 1a). A slope-calibrated PRS accurately preserves relative differences in genetic risk. When the slope is less than 1, the PRS overstates risk differences (i.e., the score is overly dispersed) and should be shrunken toward the mean; when the slope is greater than 1, the PRS underestimates risk differences (i.e., the score is too compressed) and should be stretched to match the magnitude of the true genetic signal.

**Figure 1:**
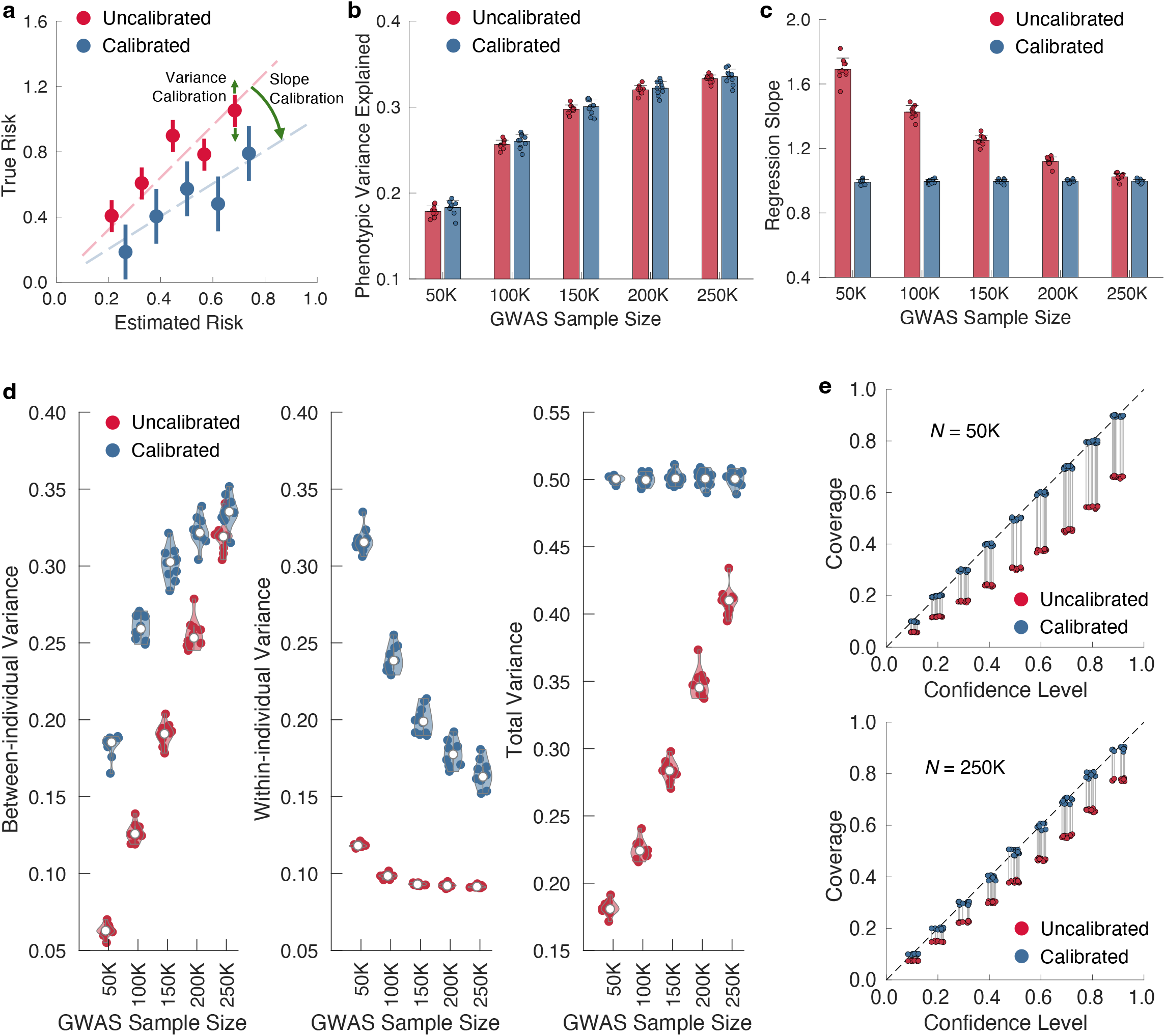
Calibration of PRSs in the primary simulation setting. **a**, Schematic illustration of slope and variance calibration. **b**, Comparison of prediction accuracy for calibrated and uncalibrated PRSs across GWAS training sample sizes. Bars and error bars indicate the mean and standard deviation across 10 simulation replicates, with individual estimates overlaid. **c**, Regression slope of the true genetic value *g*_*i*_ on the calibrated and uncalibrated posterior point estimate 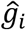 in the testing dataset across GWAS training sample sizes. Bars and error bars indicate the mean and standard deviation across 10 simulation replicates, with individual estimates overlaid. **d**, Between-individual variance of the PRS point estimate 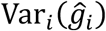 (left panel), average within-individual posterior variance 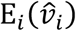 (middle panel), and total posterior predictive variance (right panel) for calibrated and uncalibrated PRSs in the testing dataset across GWAS training sample sizes. Each violin plot shows the median across 10 simulation replicates, with individual estimates overlaid. **e**, Coverage (the proportion of confidence intervals that contain the true genetic value) of calibrated and uncalibrated PRSs in the testing dataset across confidence levels for two GWAS training sample sizes, 50K (upper panel) and 250K (lower panel). Each dot represents a simulation replicate, with slight horizontal jitter for visualization.

Variance calibration assesses whether the uncertainty around the point estimate 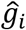 is accurately quantified (Figure 1a). A variance-calibrated PRS yields confidence intervals with correct frequentist coverage, neither systematically too wide (over-coverage, producing uninformative estimates) nor too narrow (under-coverage, giving false confidence). Calibration is often overlooked in PRS evaluation, in part because it does not affect population-level metrics such as variance explained or discrimination (e.g., area under the receiver operating characteristic curve, AUROC). However, calibration is tightly linked to uncertainty, misclassification, and inconsistency of PRS-based predictions, and is therefore critical for reliable risk stratification.

We developed methods to calibrate both slope and variance using a validation dataset with individual-level genotypes and phenotypes. For slope calibration, we rescale 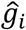 so that the regression slope of the observed phenotype on the scaled score equals 1 (Methods). For variance calibration, we show that 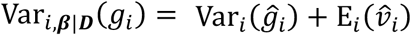, which decomposes the total posterior genetic variance into between-individual variation in PRS point estimates and average within-individual posterior uncertainty (Methods). We therefore rescale 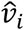 such that Var_*i*,***β***|***D***_(*g*_*i*_) matches the estimated SNP heritability 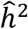, which approximates the total genetic variance captured by SNPs (Methods). For binary phenotypes, we extended both slope and variance calibration to the liability scale by appropriately rescaling 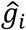 and 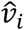 under the liability threshold model (Methods).

Bayesian PRS construction methods impose shrinkage on SNP effect sizes, which typically causes 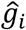 to underestimate the magnitude of the true genetic value *g*_*i*_ (Supplementary Methods). In simulations, (Supplementary Figure 1; Supplementary Table 1), calibration did not change overall prediction accuracy of the PRS, as expected (Figure 1b). However, the uncalibrated PRS exhibited a regression slope greater than 1, indicating systematic underestimation of genetic risk (Figure 1c). The total posterior predictive variance was also smaller than the true heritability (Figure 1d), leading to under-coverage of confidence intervals (Figure 1e). Calibration and coverage improved with increasing GWAS training sample size, consistent with reduced shrinkage imposed by PRS-CS as SNP effect size estimates became more precise (Figure 1c-e).

After calibration, the regression slope was consistently close to 1 across GWAS sample sizes (Figure 1c). As GWAS power increased, a larger proportion of total genetic variation was captured by between-individual variation in PRS point estimates relative to within-individual posterior uncertainty, corresponding to improved prediction accuracy (Figure 1d). At the same time, total posterior variance matched the underlying heritability (Figure 1d), and calibrated confidence intervals achieved accurate coverage across confidence levels in the testing dataset (Figure 1e).

In secondary simulations, the slope and variance calibration procedures performed well across a range of genetic architectures, as well as for binary phenotypes with varying prevalences (Supplementary Figures 2-6; Supplementary Tables 2-4). For calibrated PRSs, we further show that the traditional frequentist definition of prediction accuracy, i.e., the squared correlation between the true and predicted genetic values across individuals, 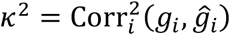, coincides with the Bayesian definition of prediction accuracy, 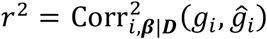, which quantifies the proportion of between-individual variance in PRS estimates, 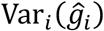, relative to total posterior genetic variance, Var_*i*,***β***|***D***_(*g*_*i*_) (Supplementary Methods). Consequently, after calibration, the explained genetic variance 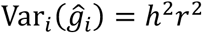 reflects prediction accuracy, whereas the remaining unexplained variance 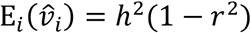 represents the residual uncertainty in individual risk estimates (Supplementary Methods). These two quantities are complementary components of the same variance decomposition.

### Misclassification

Next, we examined misclassification when thresholding PRS point estimates 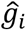 to identify individuals in the top-*q* percentile of genetic risk. Misclassification occurs when the PRS-based classification disagrees with the true class determined by the latent genetic value *g*_*i*_.

For calibrated PRSs under normality, the top-*q* percentile cutoffs for 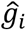 and *g*_*i*_ are *hrt*_*q*_ and *ht*_*q*_, respectively, where *t*_*q*_ is the (1 − *q*)-quantile of the standard normal distribution. The misclassification event for individual *i* can thus be represented as 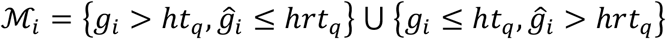. We show in Methods that the conditional posterior probability of misclassification for individual *i* given 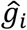 and 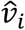 is:

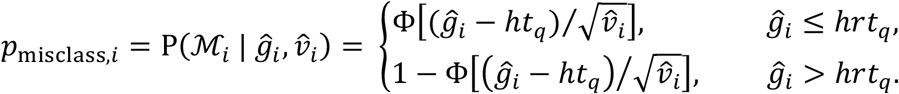

This conditional misclassification probability depends on the normalized distance between the point estimate 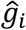 and the cutoff *ht*_*q*_. It increases monotonically for 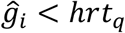 and decreases monotonically for 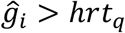, reaching its maximum at the decision boundary *hrt*_*q*_. The peak value is (Methods):

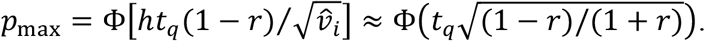

As expected, *p*_max_ → 0.5 for a perfect PRS (*r*^2^ → 1) and *p*_max_ → 1 − *q* for an uninformative PRS (*r*^2^ → 0). Intuitively, individuals near the cutoff have substantial posterior mass on both sides of the decision boundary, whereas individuals far from the cutoff are classified with high confidence.

Figure 2a compares the predicted conditional misclassification probability across PRS percentiles (upper panel) with the empirically observed misclassification rate (lower panel) in the testing dataset for three GWAS training sample sizes (50K, 100K and 250K), using the top 10% as the high-risk threshold. As expected, misclassification rates decreased with increasing GWAS sample size – reflecting improved PRS accuracy and reduced uncertainty – and peak near the 90^th^ percentile cutoff. Across all percentile bins and GWAS training sample sizes, the predicted probabilities closely aligned with observed misclassification rates, demonstrating excellent calibration of the theoretical model (Figure 2a; Supplementary Tables 5-6).

**Figure 2:**
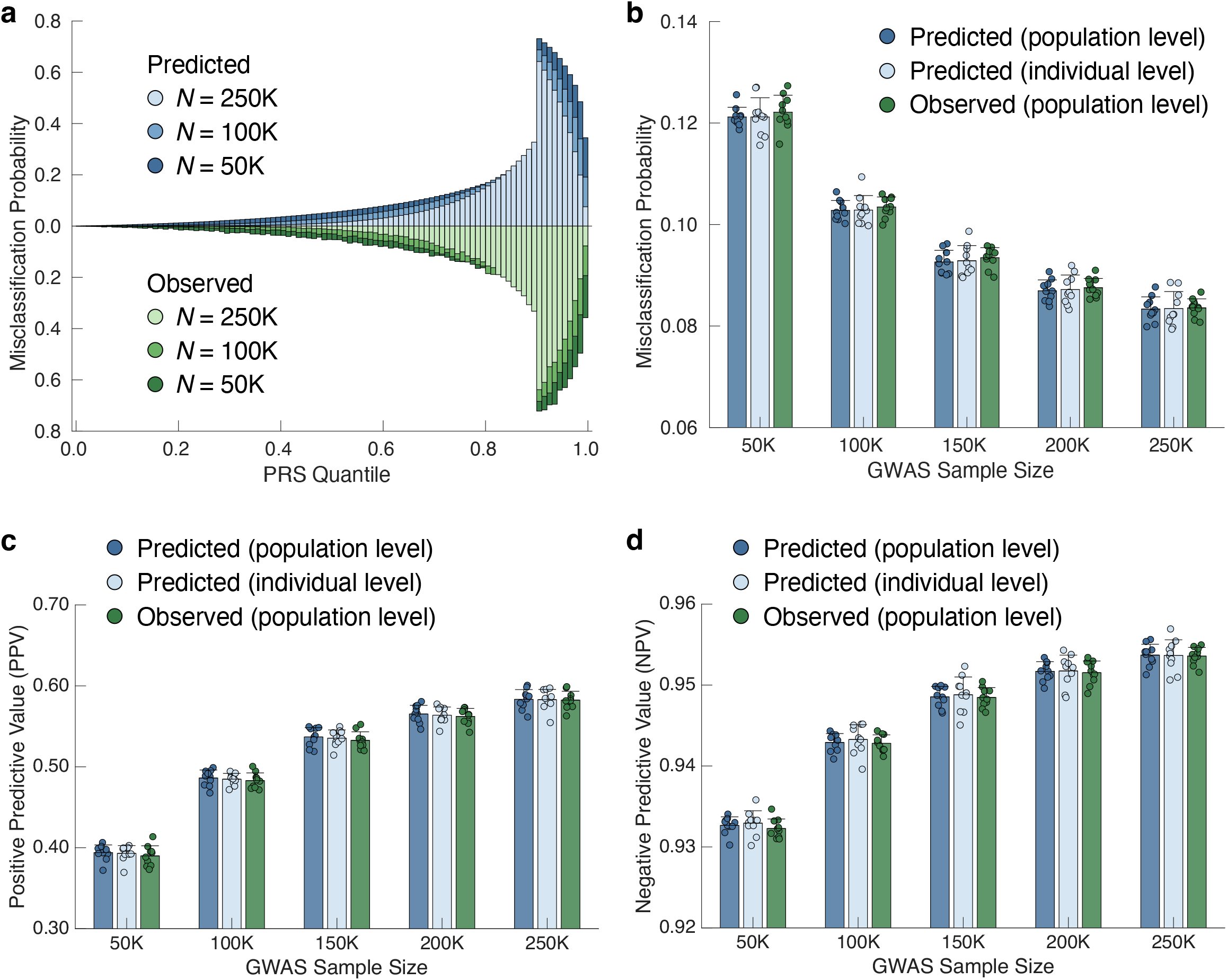
Misclassification of PRSs in the primary simulation setting. **a**, Distribution of predicted (upper panel) and empirically observed (lower panel) conditional (individual-level) misclassification probabilities across PRS percentiles in the testing dataset for three GWAS training sample sizes (50K, 100K and 250K), using the top 10% as the risk threshold. **b**, Comparison of (i) the predicted population-level misclassification probability, (ii) the average predicted individual-level conditional misclassification probability, and (iii) the empirically observed overall misclassification rate in the testing dataset across GWAS training sample sizes. **c**, Comparison of predicted and empirically observed positive predictive values (PPV) in the testing dataset across GWAS training sample sizes. **d**, Comparison of predicted and empirically observed negative predictive values (NPV) in the testing dataset across GWAS training sample sizes. In **b-d**, bars and error bars indicate the mean and standard deviation across 10 simulation replicates, with individual estimates overlaid.

At the population level, we show in Methods that the overall misclassification probability can be estimated as:

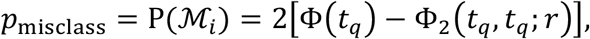

where Φ(*t*_*q*_) is the cumulative distribution function (CDF) of the standard normal distribution evaluated at *t*_*q*_, and Φ_2_(*t*_*q*_, *t*_*q*_; *r*) is the standard bivariate normal CDF with correlation *r*, evaluated at (*t*_*q*_, *t*_*q*_). Thus, population-level misclassification depends only on PRS accuracy and decreases monotonically as accuracy increases (equivalently, as PRS uncertainty decreases; Supplementary Methods). Population-level positive and negative predictive values (PPV and NPV) can be derived analogously (Methods).

Figure 2b compares (i) the predicted population-level misclassification probability, (ii) the average predicted individual-level conditional misclassification probability, and (iii) the empirically observed overall misclassification rate in the testing dataset. These three quantities were highly consistent across GWAS training sample sizes and decreased with increasing GWAS sample size (Supplementary Table 7). Corresponding PPV and NPV were also accurately predicted (Figure 2c-d; Supplementary Tables 8-9). Secondary simulations further confirmed that the theoretical predictions closely tracked misclassification risk at both individual and population levels across a range of genetic architectures, binary phenotype prevalences, and percentile cutoffs (Supplementary Figures 7-16; Supplementary Tables 5-9).

### Classification inconsistency

We next consider an individual *i* who has *K* slope- and variance-calibrated PRSs, with point estimates and posterior variances 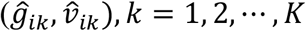. Let the prediction accuracy of the *k*-th PRS be 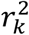, and ***C*** = [*c*_*kl*_]_*K*×*K*_ denote the correlation matrix among PRSs. The joint correlation matrix between the true genetic value *g*_*i*_ and the *K* PRSs is **Σ** = [1, ***r***^*T*^; ***r, C***], where ***r*** = (*r*_1_, *r*_2_, ⃛, *r*_*K*_)^*T*^.

Using the top-*q* percentile cutoff for each PRS, classification inconsistency is defined as any instance in which at least two classifications among 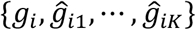 disagree. This definition depends on the true class determined by *g*_*i*_ and is equivalent to requiring that at least one PRS misclassifies individual *i*: 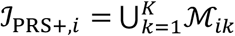, where ℳ_*ik*_ denotes the misclassification event for the *k*-th PRS. As a result, inconsistency is driven by the same uncertainty that governs misclassification and is maximized near the decision threshold.

We show in Methods that, under multivariate normal assumptions, the overall probability of classification inconsistency at the top-*q* percentile cutoff is:

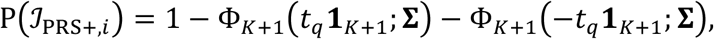

where Φ_*K*+1_[*t*_*q*_**1**_*K*+1_; **Σ**) is the CDF of a (*K* + 1)-dimensional standard multivariate normal (MVN) distribution with correlation matrix **Σ**, evaluated at the vector *t*_*q*_**1**_*K*+1_, and **1**_*K*+1_ is a (*K* + 1)-dimensional vector of ones. Thus, inconsistency is jointly determined by PRS accuracy and pairwise correlations. We further show in Supplementary Methods that this probability decreases monotonically as PRS accuracy improves or as the correlation between any pair of PRSs strengthens.

In practice, 𝒥_PRS+,*i*_ cannot be directly evaluated because the true genetic value *g*_*i*_ is unobserved. A commonly used alternative defines inconsistency based solely on agreement among the *K* PRSs^33,34^. Under this PRS-only definition, the inconsistency probability can be analogously estimated as (Methods):

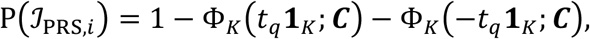

which depends only on the correlation structure ***C*** and decreases monotonically as PRSs become more strongly correlated (Supplementary Methods).

We further show that 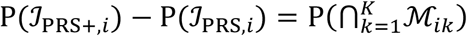 (Methods), which implies that P(𝒥_PRS,*i*_) is always smaller than P(𝒥_PRS+,*i*_). The difference arises from scenarios in which all PRSs misclassify the individual – events considered consistent under the PRS-only definition but inconsistent when the true latent genetic class is taken into account.

We assessed classification inconsistency among five PRSs under two simulation designs: (i) an overlapping setting, in which training datasets of increasing sample size partially overlapped, and (ii) an independent setting, in which training datasets of equal size were mutually independent. Individuals exceeding the top 10% of each PRS distribution were classified as high risk.

As expected, PRSs in the overlapping setting were both more accurate and more strongly correlated than those in the independent setting (Figure 3a; Supplementary Tables 10-11), resulting in lower observed inconsistency rates across percentile bins (Figure 3b). In both settings, the distribution of the with-truth inconsistency rate, P(𝒥_PRS+,*i*_) (lighter color), closely mirrored the misclassification pattern of individual PRSs (Figure 2a) and peaked near the 90^th^ percentile (Figure 3b). The PRS-only inconsistency rate, P(𝒥_PRS,*i*_) (darker color), was uniformly lower than P(𝒥_PRS+,*i*_), with the largest difference observed near the decision threshold, reflecting the increased probability that all PRSs simultaneously misclassify individuals in this region (Figure 3b).

**Figure 3:**
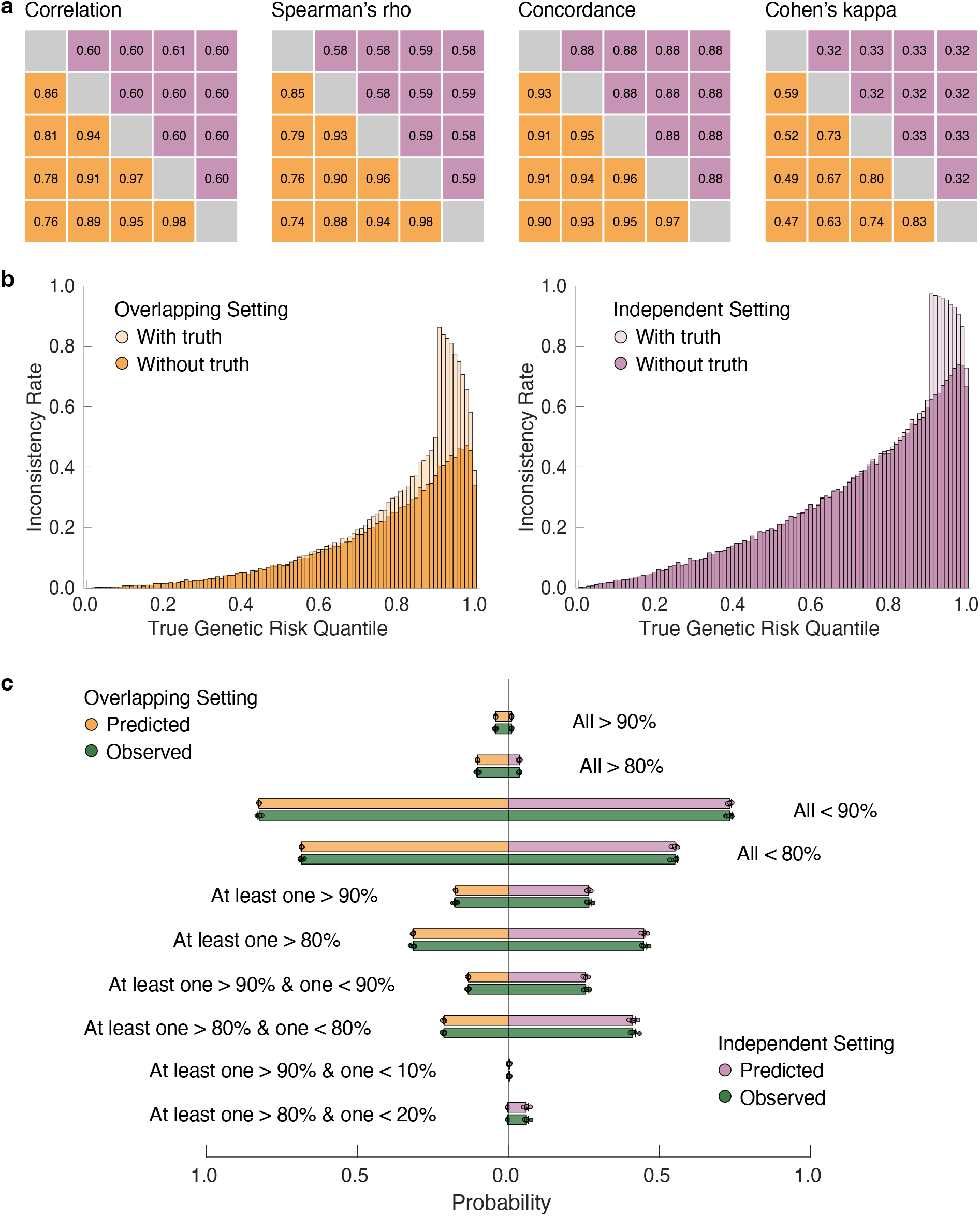
Inconsistency of PRS-based risk stratification in the primary simulation setting. **a**, Pairwise Pearson correlation and Spearman’s rho between the five PRSs, and pairwise concordance and Cohen’s kappa for high-risk classification (top 10%) between each pair of PRSs, under the overlapping (lower triangle) and independent (upper triangle) simulation settings. **b**, Distribution of empirically observed with-truth (lighter color) and PRS-only (darker color) inconsistency rates across genetic risk percentiles in the testing dataset, under the overlapping (left panel) and independent (right panel) simulation settings. **c**, Comparison of predicted and empirically observed probabilities of PRS classification profiles in the testing dataset under the overlapping (left panel) and independent (right panel) simulation settings. Bars and error bars indicate the mean and standard deviation across 10 simulation replicates, with individual estimates overlaid.

Figure 3c shows that, in both simulation settings, the predicted inconsistency probabilities closely matched the empirically observed rates in the testing dataset. Secondary simulations further demonstrated that the theoretical predictions remained robust across diverse genetic architectures, binary phenotype prevalences, and percentile cutoffs (Supplementary Figures 17-19; Supplementary Tables 12-15).

The MVN framework further enables prediction of the probability of arbitrary classification profile across PRSs (Supplementary Methods). Figure 3c illustrates several examples, including (i) the probability that all PRSs exceeded (or fell below) the 80^th^ or 90^th^ percentiles (consistent classifications); (ii) the probability that at least one PRS lay above and another below the 80^th^ or 90^th^ percentiles (inconsistent classifications); and (iii) the probability that at least one PRS exceeded the 80^th^ (or 90^th^) cutoff while another fell below the 20^th^ (or 10^th^) cutoff (extreme inconsistency). Across all scenarios and secondary simulation settings, predicted probabilities of consistency and inconsistency closely aligned with empirical estimates in the testing dataset (Supplementary Figures 20-22; Supplementary Tables 16-17).

While both the accuracy of individual PRSs and their pairwise correlations contribute to with-truth classification inconsistency, these quantities are intrinsically constrained (Methods):

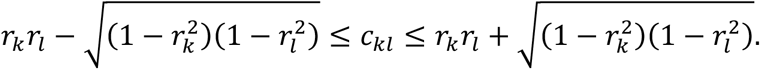

If *c*_*kl*_ = 0, the constraint reduces to 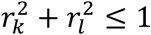, implying that two highly accurate PRSs cannot be mutually uncorrelated. Conversely, if both PRSs are near-perfect (*r*_*k*_ ≈ *r*_1_ ≈ 1), then *c*_*kl*_ ≈ 1, indicating that two highly accurate PRSs must be strongly correlated. More generally, as *r*_*k*_ and *r*_1_ increase, the feasible range of *c*_*kl*_ narrows, limiting the degree of independence among accurate PRSs.

### PRS integration and uncertainty-aware classification

Lastly, we explored strategies to reduce misclassification and between-PRS inconsistency. A widely used approach to improve PRS accuracy, thereby lowering misclassification and inconsistency, is to integrate multiple PRSs through a weighted linear combination^11,42–44^. Existing methods primarily focus on estimating weights that maximize predictive accuracy, but typically do not provide uncertainty quantification of the resulting integrated score. Given a weight vector, 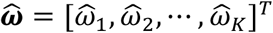 applied to *K* PRSs with individual-level point estimates 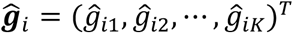, the integrated score is 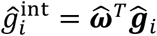 and its posterior variance would be 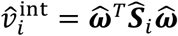, where 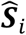 is the individual-level posterior covariance matrix among PRSs. In practice, however, 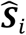 is typically unavailable because PRSs are trained independently on different datasets.

We show in Methods that, for calibrated PRSs, 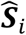 can be approximated using the prediction accuracies of individual PRSs along with their pairwise correlations. To evaluate this approach, we considered both overlapping and independent simulation designs, assuming that five PRSs became available sequentially. We then applied a non-negative regression model to iteratively integrate the available PRSs (Methods). Both the point estimates and posterior variances of the integrated PRSs were well calibrated (Supplementary Figures 23-24; Supplementary Tables 18-20). As the effective training sample size increased, a larger proportion of total genetic variation was attributed to between-individual variation in PRS estimates, reflecting improved prediction accuracy (Figure 4a-b).

**Figure 4:**
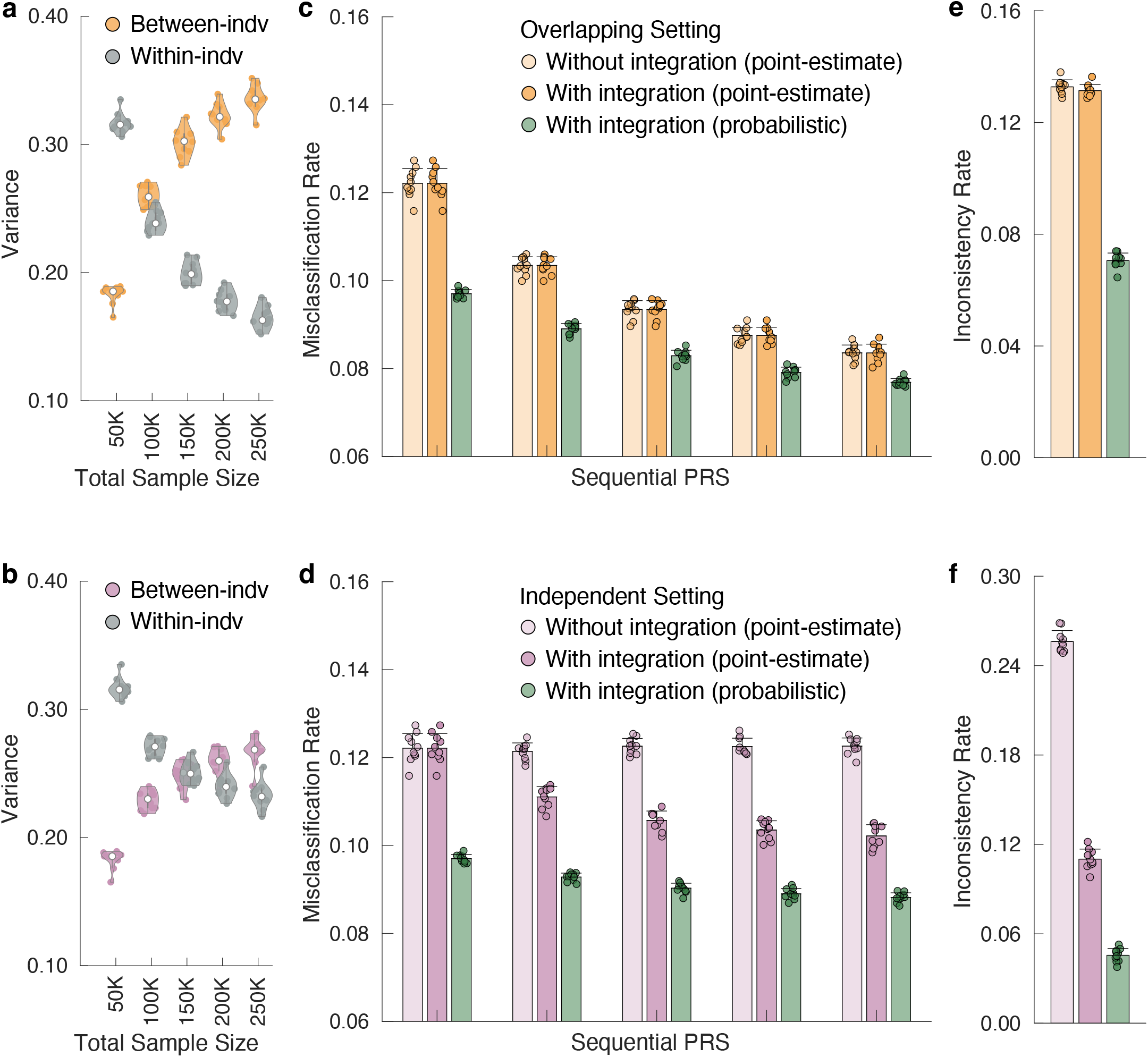
Calibration, misclassification, and classification inconsistency of integrated PRSs in the primary simulation setting. **a-b**, Between-individual variance of the PRS point estimate and average within-individual posterior variance for sequentially integrated PRSs in the testing dataset under the overlapping (upper panel) and independent (lower panel) simulation settings. Each violin plot shows the median across 10 simulation replicates, with individual estimates overlaid. **c-d**, Comparison of (i) the empirically observed misclassification rate without PRS integration using point-estimate thresholding, (ii) the empirically observed misclassification rate for sequentially integrated PRSs using point-estimate thresholding, and (iii) the empirically observed misclassification rate for sequentially integrated PRSs using uncertainty-aware probabilistic thresholding in the testing dataset under the overlapping (upper panel) and independent (lower panel) simulation settings. Bars and error bars indicate the mean and standard deviation across 10 simulation replicates, with individual estimates overlaid. **e-f**, Comparison of empirically observed classification inconsistency rates with and without PRS integration, using either point-estimate thresholding or uncertainty-aware probabilistic thresholding, in the testing dataset under the overlapping (upper panel) and independent (lower panel) simulation settings. Bars and error bars indicate the mean and standard deviation across 10 simulation replicates, with individual estimates overlaid.

In the overlapping setting, where larger GWASs subsumed samples from smaller studies, the integrated PRS provided little additional information beyond the most recent and most powerful PRS. Accordingly, misclassification and inconsistency rates in the testing dataset were nearly unchanged with integration (Figure 4c,e; Supplementary Tables 21, 23). In contrast, in the independent setting, integration increased the effective training sample size by leveraging complementary information across PRSs, resulting in lower misclassification rates and reduced classification inconsistency (Figure 4d,f; Supplementary Tables 22, 24). Similar patterns were observed across all secondary simulation settings (Supplementary Figures 25-30; Supplementary Tables 21-24).

Standard PRS applications typically identify high-risk individuals by thresholding the point estimates 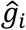, ignoring individual-specific uncertainty 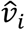. We evaluated an uncertainty-aware alternative that ranks and selects individuals based on the posterior probability that their genetic value exceeds a prespecified top-*q* percentile threshold^31^: 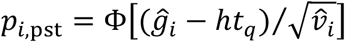. Under 0-1 loss, the Bayes-optimal decision rule classifies individual *i* as high risk if *p*_*i*,pst_ > 0.5. Unlike point-estimate thresholding, which by construction selects exactly *q*% of individuals, the probabilistic approach selects a smaller, accuracy-dependent subset (Methods).

In the overlapping setting, applying probabilistic thresholding to the five sequentially integrated PRSs selected, on average, 1.7%-5.9% of individuals for each PRS and 8.0% across any PRS, compared with 17.4% under conventional point-estimate thresholding. Despite selecting fewer individuals, uncertainty-aware stratification substantially reduced both misclassification and inconsistency rates (Figure 4c,e; Supplementary Tables 25-27). Among individuals classified as high risk across any PRS under probabilistic thresholding, 60.2% truly fell within the top 10% of genetic risk, compared with 40.6% under point-estimate thresholding, demonstrating that the probabilistic approach identifies high-risk individuals with higher confidence. Similar gains were observed in the independent setting and across secondary simulations (Figure 4d,f; Supplementary Figures 25-30; Supplementary Tables 25-27).

### Application in *All of Us*

We applied the framework of PRS calibration and uncertainty quantification to five representative complex traits and diseases in the *All of Us* Research Program (AoU): body mass index (BMI), total cholesterol (TC), coronary artery disease (CAD), type 2 diabetes (T2D), and major depressive disorder (MDD) (Supplementary Tables 28). We randomly partitioned the ancestrally-diverse AoU cohort into 10 folds, iteratively using 1 fold for validation – where PRSs were calibrated and integrated and key quantities (e.g., prediction accuracy and pairwise correlations) were estimated – and the remaining 9 folds for testing, where calibration, misclassification, and classification inconsistency were evaluated.

For each trait and disease, we curated recent large-scale GWASs in chronological order for PRS construction, yielding three to six PRSs per phenotype (Supplementary Table 29). SNP-based heritability estimates from the largest European-ancestry GWAS ranged from 7.97% (MDD) to 31.21% (T2D). Prediction accuracy varied across phenotypes and generally increased with GWAS sample size (Supplementary Table 29).

As the true genetic value is unobserved in real data, we developed methods to evaluate PRS calibration and misclassification using observed phenotypes (Methods). After slope and variance calibration, predicted disease probabilities closely matched empirically observed case proportions, whereas uncalibrated PRSs underestimated disease risk (Figure 5a-b; Supplementary Figure 31; Supplementary Table 30). As expected, calibrated PRSs exhibited larger within-individual posterior variances in East Asian and African individuals and smaller variances in European individuals, reflecting greater PRS uncertainty and lower predictive accuracy in non-European populations (Figure 5c-d; Supplementary Figures 32-33; Supplementary Table 31).

**Figure 5:**
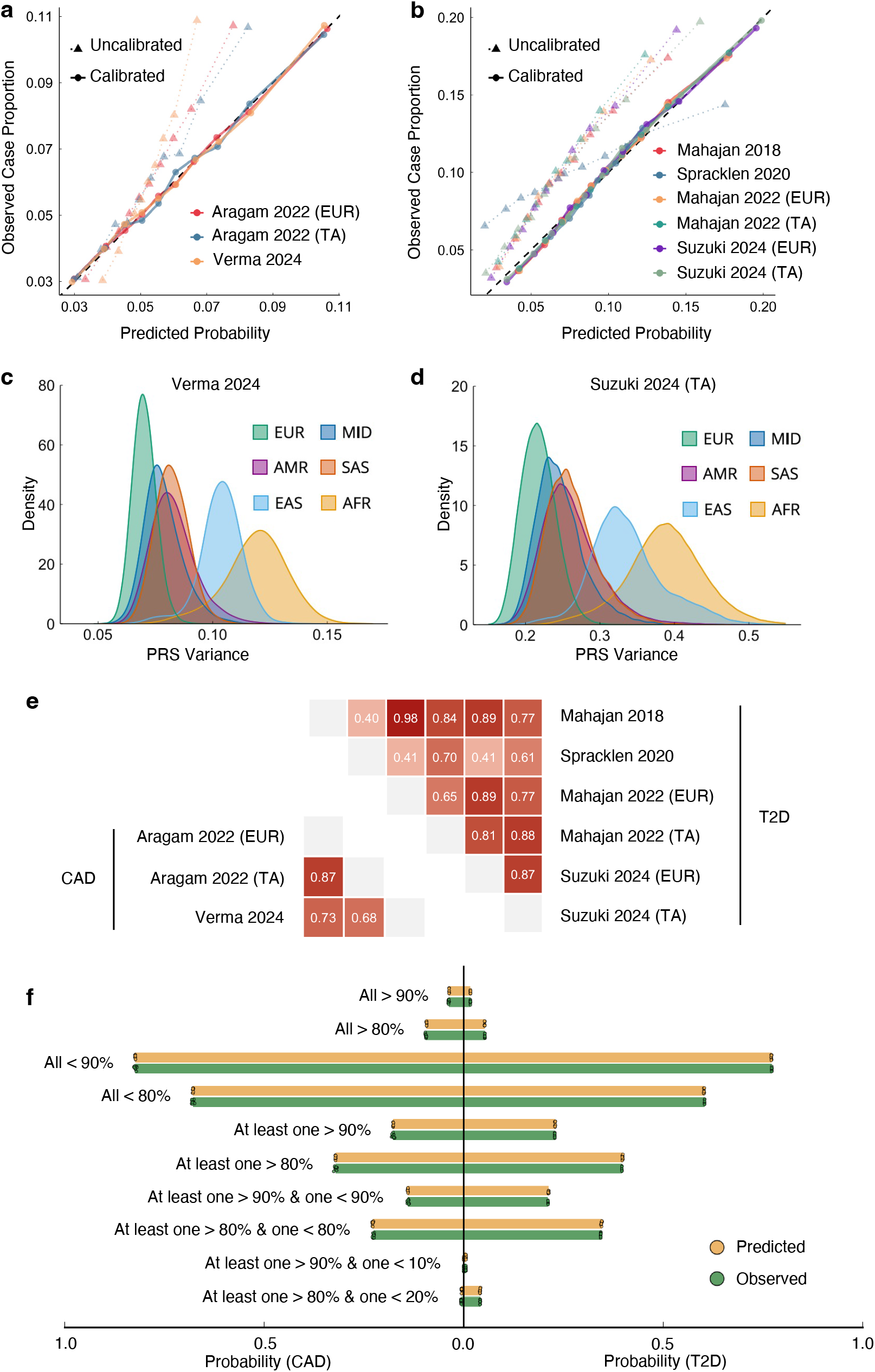
Calibration and classification inconsistency of PRSs in the AoU testing set across representative complex traits and diseases. **a-b**, Calibration of CAD and T2D PRSs, comparing observed case proportions against predicted disease probabilities. **c-d**, Distribution of within-individual posterior variance (uncertainty), stratified by ancestry, for the most recent CAD and T2D PRSs after calibration. **e**, Pairwise Pearson correlations between CAD (lower triangle) and T2D PRSs (upper triangle). **f**, Comparison of predicted and empirically observed probabilities of PRS classification profiles for CAD (left panel) and T2D (right panel). Bars indicate the mean across 10 data splits, with individual estimates overlaid.

Predicted misclassification rates, PPV, and NPV from calibrated PRSs were well aligned with empirical estimates across phenotypes and high-risk thresholds (top 10%, 5%, and 2%), supporting the validity of the framework (Supplementary Tables 32-33). Misclassification rates were higher for phenotypes that were more challenging to predict (e.g., MDD) and lower for phenotypes with more accurate PRSs (e.g., CAD). Within each phenotype, PRSs derived from larger and more recent GWASs generally exhibited lower misclassification and higher PPV, consistent with improved predictive performance (Supplementary Tables 32-33).

Pairwise correlations among calibrated PRSs were higher when derived from the same study or from GWASs with similar ancestry composition (e.g., *r* = 0.68-0.87 for CAD), but were substantially lower when incorporating GWASs from predominantly non-European ancestries^45,46^ (e.g., *r* = 0.40-0.70 for the Spracklen 2020 East Asian T2D GWAS; Figure 5e; Supplementary Table 34). Accordingly, between-PRS classification inconsistency was lower for CAD, where a small number of tightly correlated PRSs were evaluated, relative to T2D, which involved a larger number of less correlated PRSs (Figure 5f; Supplementary Table 35). Across PRS classification profiles, predicted probabilities closely matched empirically observed rates (Figure 5f; Supplementary Figure 34; Supplementary Table 36).

Finally, sequential integration of PRSs in chronological order progressively improved prediction accuracy and PPV while reducing between-PRS classification inconsistency (Figure 6a-b; Supplementary Figures 35-36; Supplementary Table 37). Gains were more pronounced for phenotypes with complementary GWAS sources (e.g., T2D: PPV increased from 0.173 to 0.197; inconsistency decreased from 0.211 to 0.097) and more modest for phenotypes with highly correlated PRSs (e.g., CAD: PPV increased from 0.108 to 0.113; inconsistency decreased from 0.139 to 0.068). Using the integrated PRSs, we further applied uncertainty-aware probabilistic thresholding to refine high-risk strata identified by top 10% point-estimate thresholds. Increasingly stringent posterior probability cutoffs substantially improved PPV and reduced between-PRS classification inconsistency, at the cost of selecting fewer individuals (Figure 6c-d; Supplementary Figures 37-38; Supplementary Table 37). As probabilistic thresholds increased, individuals of non-European ancestries, particularly African and East Asian, became progressively underrepresented among those selected, reflecting higher PRS uncertainty in these groups and the preferential selection of individuals with higher-confidence risk estimates under probabilistic thresholding (Figure 6e; Supplementary Methods; Supplementary Figures 39-40; Supplementary Table 38).

**Figure 6:**
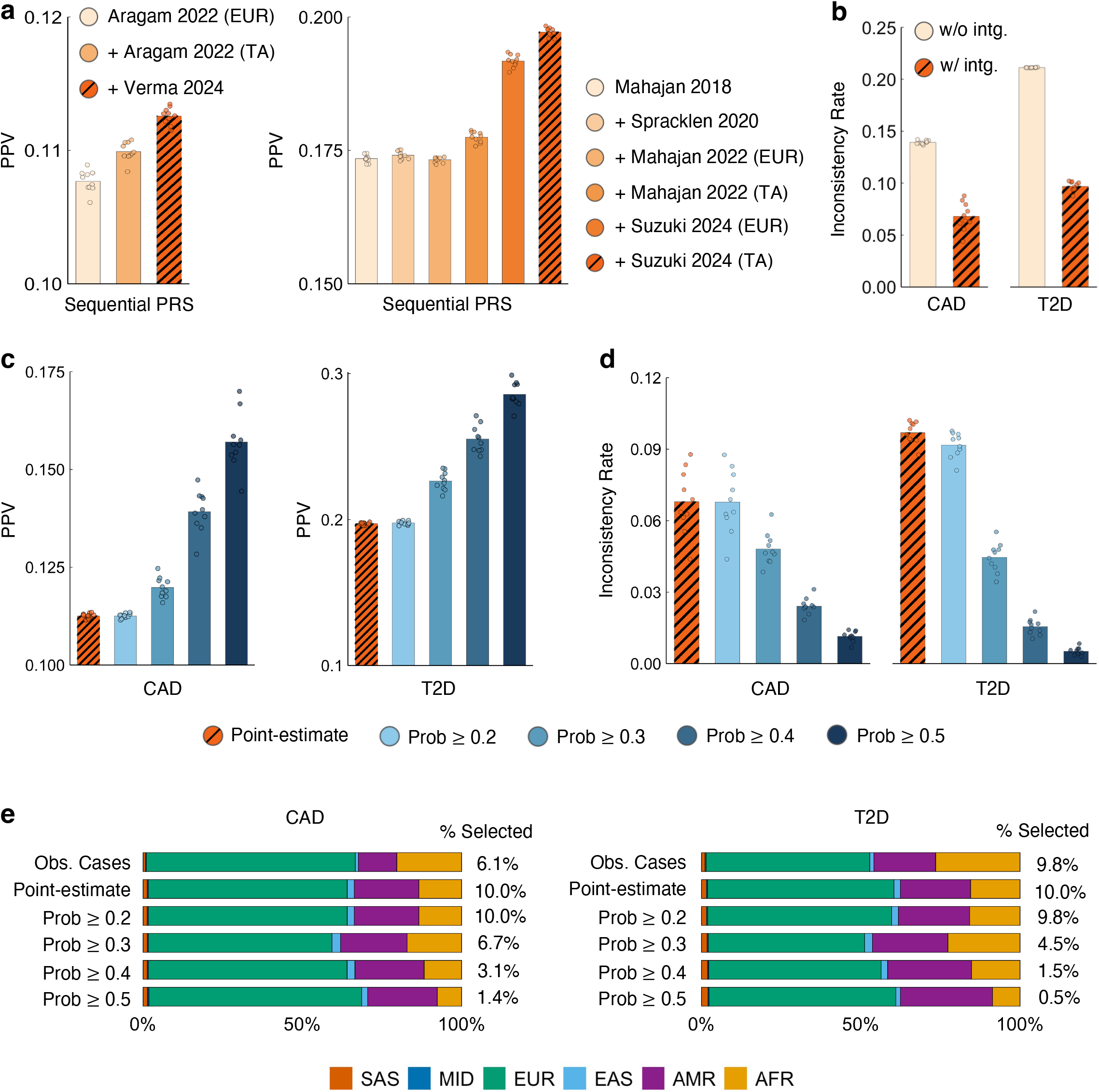
Performance of PRS integration and probabilistic thresholding in the AoU testing set for CAD and T2D. **a**, Positive predictive value (PPV) of sequentially integrated CAD and T2D PRSs using a top 10% point-estimate threshold. **b**, Comparison of empirically observed classification inconsistency rates with and without PRS integration using a top 10% point-estimate threshold. **c**, Comparison of PPV for the fully integrated CAD and T2D PRSs under point-estimate thresholding versus uncertainty-aware probabilistic thresholding across a range of posterior probability cutoffs. **d**, Comparison of observed classification inconsistency rates for the integrated CAD and T2D PRSs under point-estimate thresholding versus uncertainty-aware probabilistic thresholding across a range of posterior probability cutoffs. **e**, Genetically inferred ancestry composition of high-risk individuals selected under point-estimate and uncertainty-aware probabilistic thresholding (across varying cutoffs), compared with the ancestry composition of observed cases. In **a-d**, bars indicate the mean across 10 data splits, with individual estimates overlaid.

## Discussion

In this work, we theoretically and empirically characterized how uncertainty in PRSs shapes individual- and population-level risk stratification. We developed methods to calibrate PRS point estimates and their posterior variances for both quantitative traits and binary phenotypes, and showed that prediction accuracy and uncertainty are complementary components of total genetic variance. We further demonstrated that the probability of misclassifying high-risk individuals decreases monotonically with PRS accuracy, and that (with-truth) classification inconsistency across multiple PRSs decreases monotonically with both their accuracies and pairwise correlations. At the population level, we showed that misclassification and inconsistency rates are highly predictable in independent datasets. Supplementary Figure 41 summarizes the relationships linking PRS accuracy, uncertainty, correlation, misclassification, and inconsistency. Finally, we evaluated PRS integration and uncertainty-aware probabilistic thresholding as principled strategies to reduce misclassification and improve concordance in PRS-based risk stratification.

Calibration is often overlooked in PRS evaluation because it does not affect rank-based metrics or population-level predictive performance. However, we showed that calibration possesses important frequentist and Bayesian statistical properties and is essential for constructing valid confidence intervals for genetic risk, estimating individual-level misclassification probabilities, and enabling uncertainty-aware PRS integration and thresholding. In practice, miscalibration is pervasive and can arise from statistical overfitting and shrinkage in PRS construction, ancestry mismatch, differences in sample characteristics or disease prevalence between training and target datasets, and other sources of model misspecification. Our calibration framework provides a foundation for incorporating uncertainty into PRS-based risk stratification and improving the transparency of risk communication.

A growing literature has documented limited agreement among PRSs and raised concerns about instability in individual-level risk stratification^33–35^. These studies have largely treated discordance as an empirical pathology or a limitation of current PRS methodologies. In contrast, we showed that a substantial fraction of observed discordance is statistically expected, given the achievable prediction accuracies of individual PRSs and their attainable correlations. From this perspective, instability is not necessarily a failure of PRSs, but a predictable consequence of uncertainty interacting with hard classification thresholds.

A range of factors can reduce correlation between PRSs and contribute to classification inconsistency. Genetic architectures may differ across ancestries, demographic groups, or environmental contexts^47–49^. Phenotyping strategies may vary in specificity or emphasize distinct disease subtypes^50,51^. As a result, GWASs conducted in different samples may capture different facets of genetic risk and may be influenced by distinct sources of sampling variability. Even when PRSs are derived from the same GWAS, different algorithms rely on different modeling assumptions and can thus produce scores with similar population-level performance but divergent individual-level predictions^33^. Consequently, inconsistency between PRSs cannot be fully eliminated, as it reflects both complementary biological signals and heterogeneous sources of statistical uncertainty. Nevertheless, our theoretical framework enables accurate prediction of the expected degree of inconsistency for a given set of PRSs, offering practical guidance for PRS selection and integration. As PRS accuracy increases, correlations among PRSs are expected to strengthen and classification inconsistency will tend to decrease.

Integrating multiple PRSs through a weighted linear combination can improve prediction accuracy, thereby reducing both misclassification and inconsistency. Existing integration methods focus on estimating optimal weights via supervised or unsupervised learning^11,15,42–44^, but do not quantify uncertainty in the combined PRS. We developed a general method to approximate the posterior variance of integrated PRSs for arbitrary weight vectors. This enables direct application of our prediction theory for misclassification and inconsistency to integrated PRSs and provides a principled way to quantify and incorporate uncertainty in multi-PRS integration.

In contrast to traditional point-estimate thresholding at fixed percentiles, we examined a probabilistic thresholding approach that explicitly incorporates uncertainty in PRS estimates^31^. Under the Bayes-optimal rule for 0-1 loss, individuals are classified as high risk if the posterior probability of exceeding the genetic risk threshold is greater than 0.5. As expected, probabilistic thresholding identifies more high-risk individuals as PRS accuracy increases, reflecting greater confidence in classification. Although for a fixed confidence level probabilistic thresholding selects fewer individuals than point-estimate thresholding, we showed that the precision of selection is improved – that is, the proportion of truly high-risk individuals among those selected increases. This distinction reflects a fundamental difference in decision criteria: point-estimate thresholding fixes coverage by design, whereas probabilistic thresholding fixes confidence and prioritizes precision, selecting individuals with sufficient posterior evidence of exceeding the risk threshold.

The advantages of probabilistic thresholding are most pronounced when posterior variances vary substantially across individuals. When uncertainties are relatively homogeneous, rankings based on point estimates and posterior probabilities become highly concordant, and similar high-risk sets can be obtained by tuning thresholds for either method. Nonetheless, probabilistic thresholding provides a principled, uncertainty-aware decision rule with a clear probabilistic interpretation, and can flexibly accommodate asymmetric costs of false positives and false negatives^31^, allowing for alternative decision thresholds tailored to specific clinical contexts where the consequences of over- and under-classification differ.

This study has several limitations. First, many theoretical results rely on the assumption that PRSs and their joint distribution with the latent genetic value are approximately normal. This assumption is generally reasonable for highly polygenic traits and diseases, where PRSs aggregate small effects of many variants, but may be less accurate for phenotypes influenced by a small number of large-effect loci or in cohorts where nonrandom ascertainment skews the PRS distribution. Second, our framework relies on Bayesian inference to obtain individual-specific posterior variance estimates and therefore cannot be directly applied when only pretrained variant weights (e.g., from the PGS Catalog^52^) are available. Nevertheless, under assumptions such as constant variance across individuals, our calibration procedure can still provide population-level calibration of uncertainty. Although we focused on PRS-CS^12^, the framework extends to other Bayesian PRS construction methods^10,11,13,31,53^ that provide reliable posterior variance estimates.

Third, unlike approaches that construct confidence intervals for phenotype prediction^36–38^, we focused on uncertainty and calibration of genetic values, which are commonly used for risk stratification. As shown in the AoU analyses, our framework can be extended to phenotype prediction by incorporating residual variance. Fourth, recent studies have shown that polygenic prediction accuracy can vary across demographic, environmental, and clinical contexts^36,48,54^. While we focused on marginal calibration across contexts, the methods can be applied within specific contexts or their intersections to enable context-aware calibration and prediction. Lastly, although our theoretical results extend to prediction in ancestry-diverse cohorts such as AoU and cross-ancestry settings, this requires a validation dataset that is well matched to the genetic ancestry and sample characteristics of the target population. Challenges may arise when heritability differs across ancestry groups and reliable estimates are not available for the target population.

In summary, we developed methods to calibrate PRS point and variance estimates, quantify misclassification and inconsistency probabilities, and incorporate uncertainty into PRS integration and stratification. By establishing a unified inferential framework broadly applicable to Bayesian PRSs, we showed that individual-level misclassification and between-PRS inconsistency are not arbitrary or idiosyncratic phenomena, but predictable statistical consequences of PRS accuracy, uncertainty, and correlation structure. As translational efforts of PRSs continue to expand, explicitly accounting for uncertainty will be essential for robust risk interpretation, communication, and clinical decision-making.

## Methods

### Polygenic model

For quantitative traits, we assume a standard linear model relating genotype to phenotype for individual 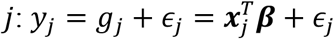, where *y*_*j*_ is standardized to have zero mean and unit variance; ***x***_*j*_ is an *M* × 1 vector of standardized genotypes; ***β*** is an *M* × 1 vector of causal genetic effects; 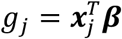 is the true genetic value; and *ϵ*_*j*_ is residual noise independent of *g*_*j*_. We assume that the heritability of the trait is *h*^2^, and thus Var_*j*_(*g*_*j*_) = *h*^2^ and Var_*j*_(*ϵ*_*j*_) = 1 − *h*^2^.

For binary phenotypes, we assume that the prevalence of the disease is *π*. Let *τ* = Φ^−1^(1 − *π*), where Φ is the CDF of the standard normal distribution. Under the liability threshold model, 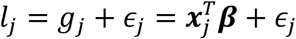 and 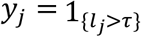, where *l*_*j*_ is the liability for individual *j* with zero mean and unit variance. Individual *j* is considered a case (*y*_*j*_ = 1) if their liability exceeds the threshold *τ*, and as a control (*y*_*j*_ = 0) otherwise. We assume that the heritability on the liability scale is *h*^2^.

### PRS estimation and uncertainty

Under a Bayesian polygenic prediction framework, a prior distribution is specified for ***β***, and the posterior distribution of SNP effect sizes is often approximated using Markov chain Monte Carlo (MCMC) methods, given GWAS summary statistics and an LD reference panel, collectively denoted by ***D***. Posterior samples of SNP effect sizes, 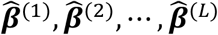, are drawn from this posterior distribution. For an unseen individual *i*, posterior samples of the PRS are calculated as 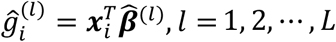. The posterior mean and variance of the PRS, 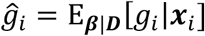 and 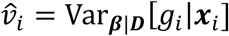, are then estimated as the empirical mean and variance of 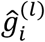 across MCMC samples.

Throughout this work, to obtain stable estimates of 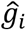 and 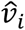, PRS-CS-auto^12^ was run for 5,000 MCMC iterations, with 2,500 burn-in steps and a thinning factor of 5, yielding 500 posterior samples per individual. Assuming that the posterior distribution of *g*_*i*_ is approximately normal, an individual-specific (1 − *α*)-level confidence interval (CI) for *g*_*i*_ can be constructed as 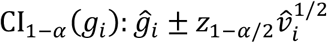, where *z*_1−*α*/2_ = Φ^−1^(1 − *α*/2).

### Slope calibration

Slope calibration evaluates whether the regression slope of the true genetic value *g*_*i*_ on the predicted score 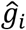 equals 1. For quantitative traits, assume the conditional mean model 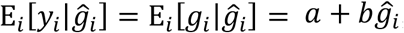, where *a* is the intercept and *b* is the slope. When both *y*_*i*_ and 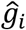 are mean-centered, *a* = 0 and 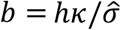, where 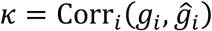 is the correlation between *g*_*i*_ and 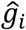 across individuals (i.e., prediction accuracy), and 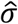 is the standard deviation of 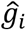 across individuals (Supplementary Methods). In practice, the slope can be estimated by fitting a linear regression: 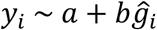. An estimate of the prediction accuracy can thus be obtained as 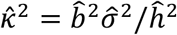, where 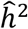 is an estimate of SNP heritability (e.g., from LD score regression^55^). Slope calibration is then achieved by rescaling the point estimate: 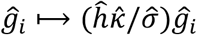.

For binary phenotypes, assume the probit liability-scale model 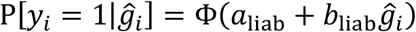, where *a*_1iab_ and *b*_1iab_ are the intercept and slope on the liability scale. By construction, 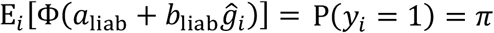. Assuming 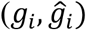 follows a bivariate normal distribution with correlation *κ*, we show in Supplementary Methods that

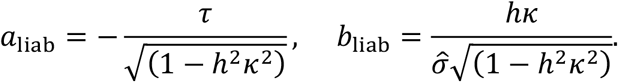

Compared with the setting for quantitative traits under the linear model, the intercept *a*_liab_ is nonzero, which ensures that the mean predicted risk matches the disease prevalence *π*. In addition, both the intercept and slope are inflated by the factor (1 − *h*^2^*κ*^2^)^−1/2^. In practice, the intercept and slope can be estimated by fitting a probit regression: 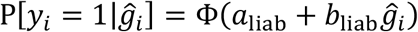. An estimate of the prediction accuracy can thus be obtained as:

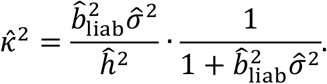

Slope calibration is then achieved by rescaling the point estimate: 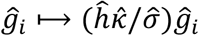.

### Variance calibration

By the law of total variance,

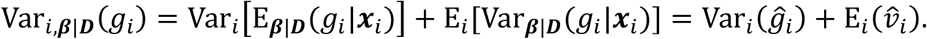

This decomposition shows that the total posterior variance of the true genetic value, Var_*i*,***β***|***D***_(*g*_*i*_), splits into two components: (i) between-individual variance of the PRS point estimate, 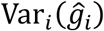, and (ii) average within-individual posterior variance, 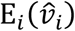. In Supplementary Methods, we show that 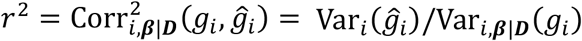, which implies that the proportion of between-individual (explained) variance relative to the total posterior variance provides a Bayesian definition of prediction accuracy. Conversely, 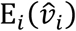 represents the remaining unexplained variance and quantifies the uncertainty in PRS estimates.

In practice, for variance calibration, we require Var_*i*,***β***|***D***_(*g*_*i*_) to match the estimated SNP heritability 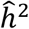, which approximates the total genetic variation captured by SNPs. After slope calibration, 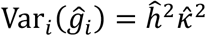, and thus variance calibration is achieved by rescaling the posterior variance 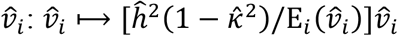. We further show in Supplementary Methods that, for slope- and variance-calibrated PRSs, the frequentist definition of prediction accuracy, 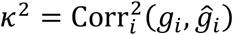, is equivalent to the Bayesian definition, 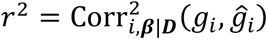.

In this work, calibration scaling factors for 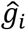 and 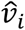 were estimated using the validation dataset and subsequently applied to individuals in the testing dataset. To evaluate PRS calibration in simulations, for each confidence level (1 − *α*), we assessed whether the empirical proportion of individuals whose confidence interval CI_1−*α*_(*g*_*i*_) covered the true genetic value *g*_*i*_ matched the nominal level (1 − *α*) in the testing dataset.

### Misclassification

Misclassification at the top-*q* percentile cutoff is defined as disagreement between the PRS-based classification and the true class determined by the latent genetic value *g*_*i*_. Let *t*_*q*_ = Φ^−1^(1 − *q*) denote the (1 − *q*)-quantile of the standard normal distribution. For calibrated PRSs, the variances of 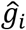 and *g*_*i*_ across individuals are *h*^2^*r*^2^ and *h*^2^, respectively. Under normal assumptions, the corresponding top-*q* thresholds are *hrt*_*q*_ for 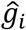 and *ht*_*q*_ for *g*_*i*_. The misclassification event can thus be represented as

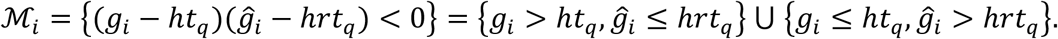

For a calibrated PRS, the conditional posterior distribution of *g*_*i*_ given ***D*** is 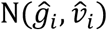. Thus, when 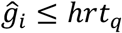,

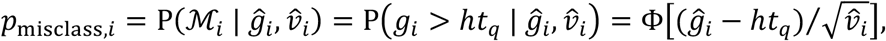

and when 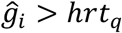,

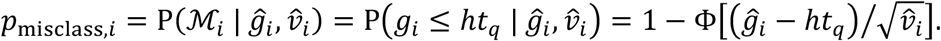

This conditional probability increases monotonically for 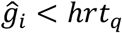 and decreases monotonically for 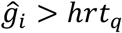, reaching its maximum at the decision boundary *hrt*_*q*_. Unless *r* = 1, the function is discontinuous at this boundary, jumping from 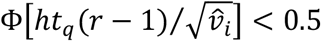 to 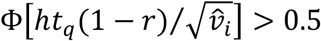. Noticing that 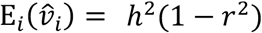 after calibration, the peak misclassification probability 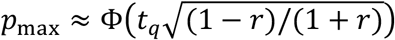.

At the population level,

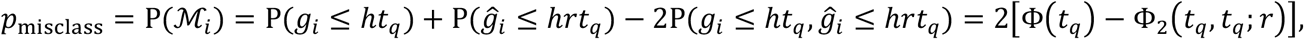

where Φ_2_(*t*_*q*_, *t*_*q*_; *r*) denotes the standard bivariate normal CDF with correlation *r*, evaluated at (*t*_*q*_, *t*_*q*_). In Supplementary Methods, we show that (i) *p*_misclass_ decreases monotonically as PRS accuracy increases, and equivalently, as PRS uncertainty decreases; and (ii) the population-level misclassification probability is well approximated by the average of the individual-level conditional posterior misclassification probabilities.

The overall PPV and NPV can be analogously estimated using conditional probabilities:

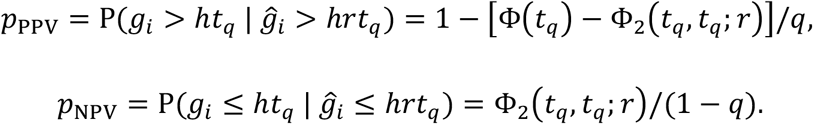

In this work, we calibrated PRSs and estimated their prediction accuracy in the validation dataset, and then predicted misclassification probabilities, both at the individual and population levels, in the testing dataset. For individual-level evaluation in simulations, the average predicted conditional misclassification probability within each PRS percentile bin was compared with the corresponding empirically observed misclassification rate. For population-level evaluation, the overall predicted misclassification probability was compared with the empirically observed misclassification rate. The top-*q* percentile cutoff in the testing dataset was determined using the PRS distribution estimated from the validation dataset.

### Classification inconsistency

Given *K* slope- and variance-calibrated PRSs for individual *i*, with point estimates and posterior variances denoted by 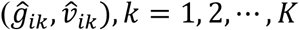, we consider two definitions of classification inconsistency.

The with-truth inconsistency, 𝒥_PRS+,*i*_, involves the true class determined by the latent genetic value *g*_*i*_ and is defined as any instance in which at least two classifications among 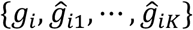 disagree. This is equivalent to requiring that at least one PRS misclassifies individual *i*: 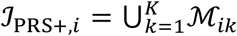, where ℳ_*ik*_ is the misclassification event for the *k*-th PRS. Assuming 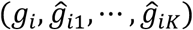 follows a MVN distribution, the probability of classification inconsistency at the top-*q* percentile cutoff is

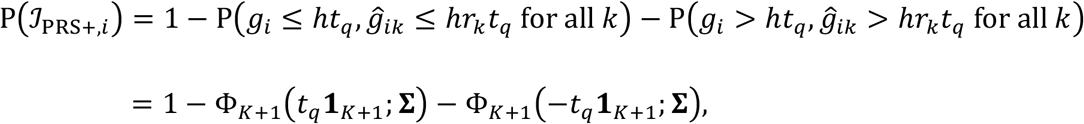

Where Φ_*K*+1_(*t*_*q*_**1**_*K*+1_; **Σ**) is the CDF of a (*K* + 1)-dimensional standard MVN distribution with correlation matrix **Σ** = [1, ***r***^*T*^; ***r, C***], evaluated at the vector *t*_*q*_**1**_*K*+1_, ***C*** is the correlation matrix among PRSs, ***r*** = (*r*_1_, *r*_2_, …, *r*_*K*_)^*T*^ is the vector of prediction accuracies, and **1**_*K*+1_ is a (*K* + 1)-dimensional vector of ones.

The PRS-only inconsistency, 𝒥_PRS,*i*_, does not incorporate the true genetic value *g*_*i*_ and evaluates agreement among the *K* PRSs alone. This definition is equivalent to requiring that at least one PRS misclassifies individual *i*, but excluding cases in which all PRS-based classifications are simultaneously incorrect: 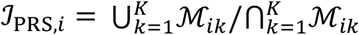. Under the MVN assumption, the probability of PRS-only classification inconsistency at the top-*q* percentile cutoff is

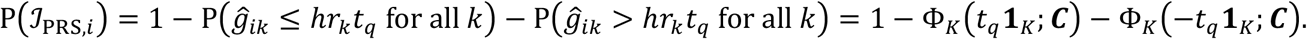

In Supplementary Methods, we show that (i) P(𝒥_PRS+,*i*_) decreases monotonically as PRS accuracy increases (equivalently, as PRS uncertainty decreases). Thus, inconsistency also declines as the misclassification probability of each PRS decreases; (ii) P(𝒥_PRS+,*i*_) decreases monotonically as correlations strengthen between any pair of PRSs; and (iii) P(𝒥_PRS,*i*_) decreases monotonically as correlations among PRSs increase.

The MVN framework enables prediction of the probability of any classification profile across PRSs. As an example, consider an extreme inconsistency event in which at least one PRS exceeds the top-*q* percentile cutoff while at least one PRS falls below the bottom-*q* percentile cutoff. Using complements and the inclusion-exclusion principle, the probability of this event can be estimated as

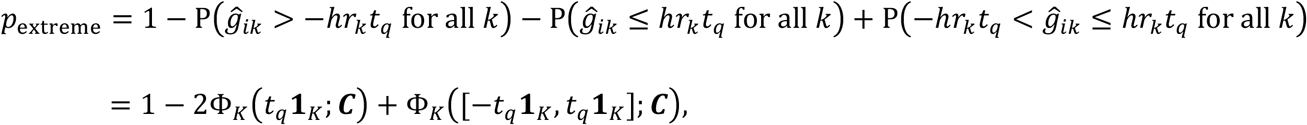

where Φ_*K*_([−*t*_*q*_**1**_*K*_, *t*_*q*_**1**_*K*_]; ***C***) denotes the MVN probability mass over the hyper-rectangle [−*t*_*q*_, *t*_*q*_]^*K*^. The derivations for probabilities of other classification profiles are provided in Supplementary Methods.

In this work, we calibrated PRSs and estimated their accuracies and correlations in the validation dataset and then used these estimates to predict classification inconsistency probabilities in the testing dataset. We compared the predicted overall inconsistency probability with the empirically observed inconsistency rate. As in previous analyses, the top-*q* percentile cutoff in the testing dataset was determined using the PRS distribution estimated from the validation dataset.

### Constraints between accuracy and correlation

For slope- and variance-calibrated PRSs, consider the correlation matrix of 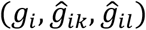:

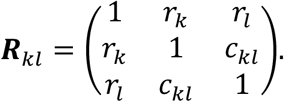

Since ***R***_*kl*_ must be positive semidefinite, det(***R***_*kl*_) ≥ 0, which yields the constraint:

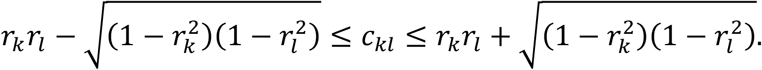

Thus, the prediction accuracy of individual PRSs and their pairwise correlations cannot vary independently; they are intrinsically constrained by one another (Supplementary Methods).

### PRS integration

Given a weight vector 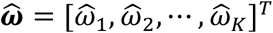 used to combine *K* PRSs, with individual-level point estimates 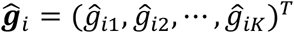, the point estimate of the integrated score is 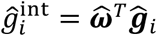. Calculation of the posterior variance of 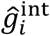 requires the individual-level posterior covariance matrix of the PRSs, which, in practice, is almost never directly available.

We show in Supplementary Methods that the posterior correlation of the PRSs, denoted as **Ω** = [Ω_*kl*_]_*K*×*K*_, can be approximated using their prediction accuracies and pairwise correlations:

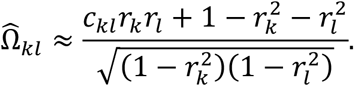

The individual-level posterior covariance matrix can then be approximated by rescaling this correlation matrix using the posterior variances of the PRSs: 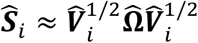, where 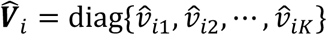 is a diagonal matrix. The posterior variance of the integrated PRS is therefore 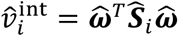. Both the integrated point estimate 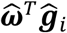 and its posterior variance 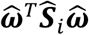 can subsequently be slope- and variance-calibrated.

In this work, we assumed that PRSs became available sequentially. Each time a new PRS was added, we fitted a non-negative regression model in the validation dataset to estimate the weights for combining all currently available PRSs^34^. These estimated weights, together with the calibration scaling factors learned in the validation dataset, were subsequently applied to individuals in the testing dataset. We assessed the calibration of the integrated PRSs and compared their misclassification and inconsistency rates with those of the individual PRSs without integration in the testing dataset.

### Uncertainty-aware classification

We evaluate an uncertainty-aware stratification approach^31^ that ranks and selects individuals based on the posterior probability that their genetic value exceeds a prespecified top-*q* percentile threshold, incorporating both the PRS point estimate 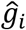 and individual-specific uncertainty 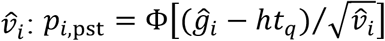. Under 0-1 loss, the Bayes-optimal decision rule classifies individual *i* as high risk if *p*_*i*,pst_ > 0.5. Since Φ(·) is monotone increasing, this decision rule is equivalent to selecting individuals with 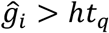. Thus, the proportion of individuals classified as high risk is 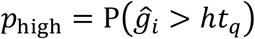. For calibrated PRSs, 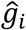 has zero mean and variance *h*^2^*r*^2^ across individuals. Under normality assumptions,

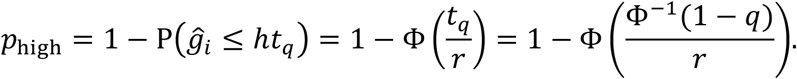

As expected, when the PRS is perfect (*r*^2^ → 1), then *p*_high_ → *q*, indicating that the PRS recovers exactly the top *q*% of individuals. Conversely, when the PRS is uninformative (*r*^2^ → 0), then *p*_high_ → 0, because the posterior probability never exceeds 0.5, and the Bayes-optimal rule classifies no one as high risk.

### Simulations

Individual-level genotypes for HapMap3 variants^39^ were simulated using HAPGEN2^40^, with the 1000 Genomes Project (1KGP)^41^ phase 3 European samples (*N* = 503) as the reference panel. In the primary simulation setting, we randomly designated 1% of HapMap3 variants as causal (polygenicity *π* = 1%) and drew their per-allele effect sizes from a normal distribution with homogeneous variance across the genome. Phenotypes were generated by adding normally distributed noise to the genetic component such that the simulated genetic effects explained 50% of phenotypic variation (SNP heritability *h*^*2*^ = 50%). This simulation procedure was repeated 10 times.

Simulated samples were divided into training, validation, and testing datasets. Training datasets, with sample sizes ranging from 50,000 to 250,000, were used to generate GWAS summary statistics, which were then provided to PRS-CS-auto^12^ to obtain posterior samples of SNP effect sizes. Validation datasets (*N* = 10,000) were used to calibrate and integrate PRSs and to estimate key quantities, including PRS accuracy and pairwise correlations, required to predict misclassification and classification inconsistency probabilities (using the top 10% as the high-risk threshold) in an independent sample. Testing datasets (*N* = 50,000) were used to evaluate PRS accuracy, calibration, misclassification, inconsistency, and integration performance. The training, validation, and testing datasets comprised non-overlapping sets of individuals.

We conducted secondary simulations varying polygenicity (*π* = 0.1% and 10%), SNP heritability (*h*^*2*^ = 20% and 80%), and percentile cutoffs (top 5% and top 2%). We additionally simulated binary phenotypes with prevalences of 10% and 20% under the liability threshold model. For binary phenotypes, GWAS summary statistics were generated using logistic regression.

### Phenotyping and data quality control in AoU

We applied the PRS calibration and uncertainty quantification framework to five representative traits and diseases in the *All of Us* Research Program (AoU): body mass index (BMI), total cholesterol (TC), coronary artery disease (CAD), type 2 diabetes (T2D), and major depressive disorder (MDD). BMI was defined using the most recent measurement of Logical Observation Identifiers Names and Codes (LOINC) code 39156-5 for each individual. TC was defined using the most recent measurement of LOINC code 2093-3, restricted to values recorded in mg/dL and within the range of 50-500. CAD cases were identified based on at least one diagnosis or procedure record of coronary artery disease or myocardial infarction (Supplementary Table 39), and controls were individuals with EHR records but no CAD diagnosis. T2D cases were defined by the presence of any diagnosis or medication prescription indicative of T2D. We excluded individuals with any type 1 diabetes diagnosis or medication records (Supplementary Table 40). Controls were defined as individuals with EHR records but no diagnosis or prescription indicative of T2D or type 1 diabetes. MDD cases were identified based on at least one diagnosis record of depressive disorder (Supplementary Table 41), with controls defined as individuals with EHR records and no MDD diagnosis. Genetic data were derived from short-read sequencing in the AoU version 8 release. For sample-level quality control, we excluded individuals flagged for quality issues or with relatedness (kinship >0.1)^56^. For variant-level quality control, we excluded flagged variants, multiallelic variants, and variants with MAF < 0.005 or call rate < 0.9. The final analytical sample included individuals with both curated phenotypes and genetic data across ancestry groups (Supplementary Table 28). Genetic principal components (PCs) were extracted from the AoU v8 release. Age was calculated based on the most recent survey date and date of birth, and sex was determined from sex chromosomes.

### Analysis in AoU

We randomly partitioned the ancestrally-diverse AoU cohort into 10 folds, iteratively using 1 fold for validation and the remaining 9 folds for testing. All results were averaged across the 10 splits. For each trait and disease, we curated recent large-scale GWASs in chronological order to construct PRSs, yielding three to six PRSs per phenotype (Supplementary Table 29). SNP-based heritability was estimated from the largest European-ancestry GWAS using LD score regression^55^. PRS-CS-auto^12^ was used to estimate the posterior mean 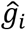 and variance 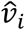 for each PRS and individual based on 500 posterior samples after burn-in and thinning. Age, sex, and the top 16 PCs were regressed out from the PRSs.

Because the true genetic value is unobserved in real data analysis, we evaluated slope and variance calibration through observable implications in the testing dataset (Supplementary Methods). For quantitative traits, calibration was assessed via empirical coverage of the (1 − *α*)-level posterior predictive interval for the observed phenotype:

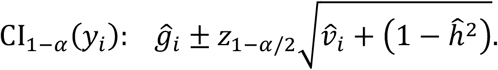

For binary phenotypes, we evaluated calibration on the observed scale. For an unseen individual *i*, the predicted disease probability is

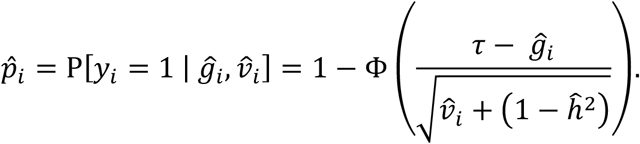

Calibration was then assessed by binning individuals according to 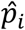 and comparing the observed case proportions against average predicted probabilities within each bin.

To evaluate misclassification without access to the true genetic value, we derived individual- and population-level misclassification probabilities for observed phenotypes (Supplementary Methods). For quantitative traits, misclassification at the top-*q* percentile cutoff was defined by disagreement between PRS-based classification and whether the observed phenotype fell within the corresponding empirical top-*q* percentile of the phenotype distribution in the testing set. For binary phenotypes, misclassification was defined as disagreement between the PRS-based classification and observed disease status. Predicted PPV and NPV for observed phenotypes were evaluated analogously.

For each phenotype, PRSs were sequentially integrated using non-negative regression following the chronological order of GWAS availability. Calibration, prediction accuracy, and PPV were evaluated at each integration step. To assess uncertainty-aware probabilistic thresholding, we further stratified high-risk individuals identified by top 10% point-estimate thresholds using the posterior probability that their genetic value exceeds the top 10% threshold. A posterior probability threshold of 0.5 corresponds to the Bayes-optimal decision rule under 0-1 loss, while less stringent cutoffs (0.2, 0.3, and 0.4) provide trade-offs between confidence and the number of individuals selected. PPV and between-PRS classification inconsistency were compared between point-estimate and probabilistic thresholding approaches.

## Acknowledgements

We gratefully acknowledge *All of Us* participants for their contributions, without whom this research would not have been possible. We also thank the National Institutes of Health’s *All of Us* Research Program for making available the participant data examined in this study. Y.Z. is supported by a Tommy Fuss Scholar in Precision Psychiatry Award. T.G. is supported by R01HG012354, R01MH130899, and U01HG011723.

## Data Availability

This study used data from the *All of Us* Research Program’s Controlled Tier Dataset v8, available to authorized users on the Research Workbench (https://workbench.researchallofus.org).

## Code Availability

PRS-CS-auto: https://github.com/getian107/PRScs

HAPGEN2: https://mathgen.stats.ox.ac.uk/genetics_software/hapgen/hapgen2.html

Analysis code: https://github.com/getian107/PRS-uncertainty

