## Supplementary Information for "Characterizing the Uncertainty, Misclassification and Inconsistency of Polygenic Prediction"

### Supplementary Methods

**Polygenic model.** For quantitative traits, we assume the standard linear model relating genotype to phenotype for individual $j$:

$$y_j = g_j + \epsilon_j = \boldsymbol{x}_j^T \boldsymbol{\beta} + \epsilon_j,$$

where $y_j$ has zero mean and unit variance; $\boldsymbol{x}_j$ is an $M \times 1$ vector of standardized genotypes; $\boldsymbol{\beta}$ is an $M \times 1$ vector of causal effect sizes; $g_j = \boldsymbol{x}_j^T \boldsymbol{\beta}$ is the true genetic value; and $\epsilon_j$ is residual noise independent of $g_j$. We assume that the trait has heritability $h^2$, and thus $\mathrm{Var}_j(g_j) = h^2$ and $\mathrm{Var}_j(\epsilon_j) = 1 - h^2$.

For binary phenotypes, we assume that the prevalence of the disease is $\pi$. Let $\tau = \Phi^{-1}(1 - \pi)$, where $\Phi$ is the cumulative distribution function (CDF) of the standard normal distribution. Under the liability threshold model,

$$l_j = g_j + \epsilon_j = \boldsymbol{x}_j^T \boldsymbol{\beta} + \epsilon_j, \qquad y_j = 1_{\{l_j > \tau\}},$$

where $l_j$ is the latent liability with zero mean and unit variance. Individual $j$ is considered a case ($y_j = 1$) if the liability exceeds the threshold $\tau$, and a control ($y_j = 0$) otherwise. We assume that the heritability on the liability scale is $h^2$, and thus $\mathrm{Var}_j(g_j) = h^2$ and $\mathrm{Var}_j(\epsilon_j) = 1 - h^2$.

**Bayesian PRS construction and posterior inference.** Under a Bayesian polygenic prediction framework, a prior distribution is specified for the effect size vector $\boldsymbol{\beta}$, and its posterior distribution is often approximated using Markov chain Monte Carlo (MCMC) methods, given GWAS summary statistics and an LD reference panel, collectively denoted by $\boldsymbol{D}$. Let $\widehat{\boldsymbol{\beta}}^{(1)}, \widehat{\boldsymbol{\beta}}^{(2)}, \cdots, \widehat{\boldsymbol{\beta}}^{(L)}$ denote posterior samples of SNP effect sizes. For an unseen individual $i$, the corresponding posterior samples of the PRS can be calculated as

$$\hat{g}_i^{(l)} = \boldsymbol{x}_i^T \widehat{\boldsymbol{\beta}}^{(l)}, \quad l = 1, 2, \cdots, L.$$

The posterior mean and variance of the PRS,

$$\hat{g}_i = \mathrm{E}_{\boldsymbol{\beta}|\boldsymbol{D}}[g_i | \boldsymbol{x}_i], \quad \hat{v}_i = \mathrm{Var}_{\boldsymbol{\beta}|\boldsymbol{D}}[g_i | \boldsymbol{x}_i],$$

are then estimated by the empirical mean and variance of $\{\hat{g}_i^{(l)}\}_{l=1}^{L}$. The posterior mean $\hat{g}_i$ serves as the point estimate of the PRS, while the posterior variance $\hat{v}_i$ quantifies individual-specific uncertainty in $\hat{g}_i$.

In practice, a threshold $t$ is often applied to $\hat{g}_i$ to identify individuals at elevated genetic risk. If the posterior distribution of $g_i$ is approximately normal, an individual-specific $(1 - \alpha)$-level confidence interval (CI) for $g_i$ can be constructed as

$$\mathrm{CI}_{1-\alpha}(g_i): \quad \hat{g}_i \pm z_{1-\alpha/2} \hat{v}_i^{1/2},$$

where $z_{1-\alpha/2}$ is the $(1 - \alpha/2)$-quantile of the standard normal distribution.

**Slope calibration.** Slope calibration evaluates whether the regression slope of the true genetic value $g_i$ on the predicted risk score $\hat{g}_i$ equals 1. A slope-calibrated PRS preserves relative differences in genetic risk. When the slope is less than 1, the PRS overstates risk differences (the distribution is overly dispersed) and should be shrunken toward the mean. When the slope is greater than 1, the PRS underestimates risk differences (the distribution is too compressed) and should be stretched to match the magnitude of the true genetic signal.

For quantitative traits, assume the conditional mean model

$$\mathrm{E}_i[y_i \mid \hat{g}_i] = \mathrm{E}_i[g_i \mid \hat{g}_i] = a + b\hat{g}_i,$$

where $a$ is the intercept and $b$ is the slope. When both $y_i$ and $\hat{g}_i$ are mean-centered, $a = 0$. Let $\kappa = \mathrm{Corr}_i(g_i, \hat{g}_i)$ be the correlation between $g_i$ and $\hat{g}_i$ across individuals, and $\hat{\sigma} = \mathrm{Var}_i^{1/2}(\hat{g}_i)$ be the standard deviation of $\hat{g}_i$ across individuals. The slope is then

$$b = \frac{\mathrm{Cov}_i(g_i, \hat{g}_i)}{\mathrm{Var}_i(\hat{g}_i)} = \frac{\kappa h \hat{\sigma}}{\hat{\sigma}^2} = \frac{\kappa h}{\hat{\sigma}}.$$

For binary phenotypes, assume the conditional mean model

$$\mathrm{P}[y_i = 1 \mid \hat{g}_i] = \Phi(a_{\mathrm{liab}} + b_{\mathrm{liab}}\hat{g}_i),$$

where $a_{\mathrm{liab}}$ and $b_{\mathrm{liab}}$ denote the intercept and slope on the liability scale. By construction,

$$\mathrm{E}_i[\Phi(a_{\mathrm{liab}} + b_{\mathrm{liab}}\hat{g}_i)] = \mathrm{P}(y_i = 1) = \pi.$$

Assuming $(g_i, \hat{g}_i)$ follows a bivariate normal distribution with correlation $\kappa$, the conditional distribution of $g_i$ given $\hat{g}_i$ is also normal:

$$g_i \mid \hat{g}_i \sim \mathrm{N}\left(\frac{h\kappa}{\hat{\sigma}}\hat{g}_i, h^2(1 - \kappa^2)\right).$$

Since $\epsilon_i$ is independent of $g_i$ with mean zero and variance $1 - h^2$, the conditional distribution of the liability is:

$$l_i \mid \hat{g}_i \sim \mathrm{N}\left(\frac{h\kappa}{\hat{\sigma}}\hat{g}_i, 1 - h^2\kappa^2\right).$$

Therefore,

$$\mathrm{P}[y_i = 1 \mid \hat{g}_i] = \mathrm{P}[l_i > \tau \mid \hat{g}_i] = \mathrm{P}\left[z_i > \frac{\tau}{\sqrt{(1 - h^2\kappa^2)}} - \frac{h\kappa}{\hat{\sigma}\sqrt{(1 - h^2\kappa^2)}}\hat{g}_i\right]$$

$$= \Phi\left(-\frac{\tau}{\sqrt{(1 - h^2\kappa^2)}} + \frac{h\kappa}{\hat{\sigma}\sqrt{(1 - h^2\kappa^2)}}\hat{g}_i\right),$$

where $z_i$ follows the standard normal distribution. This suggests that

$$a_{\text{liab}} = -\frac{\tau}{\sqrt{(1-h^2\kappa^2)}}, \quad b_{\text{liab}} = \frac{h\kappa}{\hat{\sigma}\sqrt{(1-h^2\kappa^2)}}.$$

We note that compared with calibration for quantitative traits under the linear model, the intercept $a_{\text{liab}}$ is nonzero, which ensures that the mean predicted risk matches the population prevalence $\pi$. In addition, both the intercept and slope are inflated by the factor $(1-h^2\kappa^2)^{-1/2}$, because the conditional variance of the liability $l_i$ given $\hat{g}_i$ is

$$\text{Var}_i(l_i \mid \hat{g}_i) = 1 - h^2\kappa^2,$$

which shrinks as prediction accuracy $\kappa^2$ increases. Under the nonlinear probit link, the intercept and slope must therefore be rescaled to maintain the overall prevalence at $\pi$.

**Variance calibration.** Variance calibration assesses whether the uncertainty around the point estimate $\hat{g}_i$ is accurately quantified. A variance-calibrated PRS produces confidence intervals with accurate frequentist coverage, neither systematically too wide (over-coverage, leading to uninformative estimates) nor too narrow (under-coverage, giving false confidence).

By the law of total variance,

$$\text{Var}_{i,\boldsymbol{\beta}|\boldsymbol{D}}(g_i) = \text{Var}_i\left[\text{E}_{\boldsymbol{\beta}|\boldsymbol{D}}(g_i|\boldsymbol{x}_i)\right] + \text{E}_i[\text{Var}_{\boldsymbol{\beta}|\boldsymbol{D}}(g_i|\boldsymbol{x}_i)] = \text{Var}_i(\hat{g}_i) + \text{E}_i(\hat{v}_i).$$

This decomposition shows that the total posterior variance of the true genetic value, $\text{Var}_{i,\boldsymbol{\beta}|\boldsymbol{D}}(g_i)$, splits into two components: (i) between-individual variance of the PRS point estimate, $\text{Var}_i(\hat{g}_i)$; and (ii) within-individual posterior uncertainty, averaged across individuals, $\text{E}_i(\hat{v}_i)$. For variance calibration, we require that $\text{Var}_{i,\boldsymbol{\beta}|\boldsymbol{D}}(g_i) = \text{Var}_i(g_i) = h^2$, that is, the total posterior variance of $g_i$ matches the true genetic variance across individuals.

**Bayesian prediction accuracy.** Bayesian PRS prediction accuracy is defined as

$$r^2 = \text{Corr}^2_{i,\boldsymbol{\beta}|\boldsymbol{D}}(g_i, \hat{g}_i),$$

the posterior squared correlation between $g_i$ and $\hat{g}_i$ across individuals. We first compute the posterior covariance. Using the law of total expectation and noticing that $\hat{g}_i$ is deterministic given $\boldsymbol{D}$,

$$\begin{aligned}\text{Cov}_{i,\boldsymbol{\beta}|\boldsymbol{D}}(g_i, \hat{g}_i) &= \text{E}_{i,\boldsymbol{\beta}|\boldsymbol{D}}(g_i\hat{g}_i) - \text{E}_{i,\boldsymbol{\beta}|\boldsymbol{D}}(g_i)\text{E}_{i,\boldsymbol{\beta}|\boldsymbol{D}}(\hat{g}_i) \\ &= \text{E}_i\left[\text{E}_{\boldsymbol{\beta}|\boldsymbol{D}}(g_i\hat{g}_i|\boldsymbol{x}_i)\right] - \text{E}_i\left[\text{E}_{\boldsymbol{\beta}|\boldsymbol{D}}(g_i|\boldsymbol{x}_i)\right]\text{E}_i\left[\text{E}_{\boldsymbol{\beta}|\boldsymbol{D}}(\hat{g}_i|\boldsymbol{x}_i)\right] \\ &= \text{E}_i(\hat{g}_i^2) - \text{E}_i^2(\hat{g}_i) = \text{Var}_i(\hat{g}_i).\end{aligned}$$

Therefore,

$$r^2 = \mathrm{Corr}^2_{i,\boldsymbol{\beta}|\boldsymbol{D}}(g_i, \hat{g}_i) = \frac{\mathrm{Cov}^2_{i,\boldsymbol{\beta}|\boldsymbol{D}}(g_i, \hat{g}_i)}{\mathrm{Var}_{i,\boldsymbol{\beta}|\boldsymbol{D}}(g_i)\mathrm{Var}_{i,\boldsymbol{\beta}|\boldsymbol{D}}(\hat{g}_i)} = \frac{\mathrm{Var}_i(\hat{g}_i)}{\mathrm{Var}_{i,\boldsymbol{\beta}|\boldsymbol{D}}(g_i)} = 1 - \frac{\mathrm{E}_i(\hat{v}_i)}{\mathrm{Var}_{i,\boldsymbol{\beta}|\boldsymbol{D}}(g_i)}.$$

The last equity follows from the variance decomposition in the previous section. This expression shows that Bayesian prediction accuracy is the proportion of the total posterior variance of the true genetic value explained by between-individual variation in the PRS point estimate. The term $\mathrm{E}_i(\hat{v}_i)$ represents the remaining within-individual unexplained variance and quantifies posterior uncertainty in the PRS estimates. Thus, accuracy and uncertainty are complementary components of the same variance decomposition: as PRS performance improves, posterior uncertainty decreases and a larger fraction of the genetic variance is captured across individuals.

**Calibration in practice.** For quantitative traits, we fit a linear regression, $y_i \sim a + b\hat{g}_i$, to obtain an estimate of the slope $\hat{b}$. An empirical estimate of prediction accuracy is then

$$\hat{\kappa}^2 = \frac{\hat{b}^2\hat{\sigma}^2}{\hat{h}^2},$$

where $\hat{h}^2$ is an estimate of SNP heritability (e.g., from LD score regression).

For binary phenotypes, we fit a probit regression, $\mathrm{P}[y_i = 1|\hat{g}_i] = \Phi(a_{\mathrm{liab}} + b_{\mathrm{liab}}\hat{g}_i)$, to obtain estimates of the intercept and slope on the liability scale, $\hat{a}_{\mathrm{liab}}$ and $\hat{b}_{\mathrm{liab}}$. The corresponding estimate of prediction accuracy is

$$\hat{\kappa}^2 = \frac{\hat{b}^2_{\mathrm{liab}}\hat{\sigma}^2}{\hat{h}^2} \cdot \frac{1}{1 + \hat{b}^2_{\mathrm{liab}}\hat{\sigma}^2}.$$

For both quantitative traits and binary phenotypes, slope and variance calibration are achieved by rescaling the PRS point estimate $\hat{g}_i$ and its posterior variance $\hat{v}_i$:

$$(\hat{g}_i, \hat{v}_i) \longmapsto \left(\frac{\hat{h}\hat{\kappa}}{\hat{\sigma}}\hat{g}_i, \frac{\hat{h}^2(1-\hat{\kappa}^2)}{\mathrm{E}_i(\hat{v}_i)}\hat{v}_i\right).$$

After calibration, it follows that $\mathrm{Var}_i(\hat{g}_i) = \hat{h}^2\hat{\kappa}^2$, $\mathrm{E}_i(\hat{v}_i) = \hat{h}^2(1-\hat{\kappa}^2)$, and $\mathrm{Var}_{i,\boldsymbol{\beta}|\boldsymbol{D}}(g_i) = \hat{h}^2$, confirming that the calibrated PRS satisfies both slope and variance calibration conditions.

**Implications of slope calibration.** We first note that since slope calibration scales $\hat{g}_i$ by a global multiplicative factor, it does not affect population-level prediction accuracy and discrimination metrics such as the phenotypic variance explained for quantitative traits and area under the receiver operating characteristic curve (AUROC) for binary phenotypes.

Define $g_i^* = \mathrm{E}(g_i|\boldsymbol{x}_i)$, where the expectation is taken under the true data-generating distribution of $(g_i, \boldsymbol{x}_i)$, independent of any PRS model or dataset. Since both $y_i$ and $\hat{g}_i$ are mean-centered, slope calibration requires

$$\mathrm{Cov}_i(g_i, \hat{g}_i) = \mathrm{Var}_i(\hat{g}_i).$$

Meanwhile,

$$\mathrm{Cov}_i(g_i, \hat{g}_i) = \mathrm{E}_i(g_i\hat{g}_i) - \mathrm{E}_i(g_i)\mathrm{E}_i(\hat{g}_i) = \mathrm{E}_i[\mathrm{E}(g_i\hat{g}_i|\boldsymbol{x}_i)] - \mathrm{E}_i[\mathrm{E}(g_i|\boldsymbol{x}_i)]\mathrm{E}_i(\hat{g}_i)$$

$$= \mathrm{E}_i(g_i^*\hat{g}_i) - \mathrm{E}_i(g_i^*)\mathrm{E}_i(\hat{g}_i) = \mathrm{Cov}_i(g_i^*, \hat{g}_i) = \mathrm{Cov}_i(\hat{g}_i - b_i^*, \hat{g}_i) = \mathrm{Var}_i(\hat{g}_i) - \mathrm{Cov}_i(\hat{g}_i, b_i^*),$$

where $b_i^* = \hat{g}_i - g_i^*$ denotes the bias of the point estimate. Therefore, slope calibration implies $\mathrm{Cov}_i(\hat{g}_i, b_i^*) = 0$, i.e., the bias term is uncorrelated with the predicted score.

If the PRS is a shrunken version of the true genetic signal, $\hat{g}_i = sg_i^*, 0 < s < 1$, as is typical for Bayesian PRSs, then $b_i^* = \hat{g}_i - g_i^* = (s-1)g_i^*$, and

$$\mathrm{Cov}_i(\hat{g}_i, b_i^*) = \mathrm{Cov}_i(sg_i^*, (s-1)g_i^*) = s(s-1)\mathrm{Var}_i(g_i^*) < 0.$$

Thus, shrinkage induces negative correlation between the point estimate and its bias, implying a regression slope greater than 1.

If the PRS is unbiased but noisy, $\hat{g}_i = g_i^* + \varrho_i$, where $\varrho_i$ is a noise term independent of $g_i^*$, then $b_i^* = \varrho_i$, and

$$\mathrm{Cov}_i(\hat{g}_i, b_i^*) = \mathrm{Cov}_i(g_i^* + \varrho_i, \varrho_i) = \mathrm{Var}_i(\varrho_i) > 0.$$

Thus, noise induces positive correlation between the point estimate and its bias, implying a regression slope less than 1.

In practice, both shrinkage and noise may be present. If $\hat{g}_i = sg_i^* + \varrho_i$, then $b_i^* = (s-1)g_i^* + \varrho_i$, and

$$\mathrm{Cov}_i(\hat{g}_i, b_i^*) = \mathrm{Cov}_i(sg_i^* + \varrho_i, (s-1)g_i^* + \varrho_i) = s(s-1)\mathrm{Var}_i(g_i^*) + \mathrm{Var}_i(\varrho_i).$$

The covariance can be either negative or positive, depending on whether shrinkage or noise dominates. It equals zero when $\mathrm{Var}_i(\varrho_i) = s(1-s)\mathrm{Var}_i(g_i^*)$.

**Equivalence of frequentist and Bayesian accuracy under calibration.** For slope-calibrated PRSs, the frequentist prediction accuracy is

$$\kappa^2 = \mathrm{Corr}_i^2(g_i, \hat{g}_i) = \frac{\mathrm{Cov}_i^2(g_i, \hat{g}_i)}{\mathrm{Var}_i(g_i)\mathrm{Var}_i(\hat{g}_{ik})} = \frac{[\mathrm{Var}_i(\hat{g}_i) - \mathrm{Cov}_i(\hat{g}_i, b_i^*)]^2}{\mathrm{Var}_i(g_i)\mathrm{Var}_i(\hat{g}_i)} = \frac{\mathrm{Var}_i(\hat{g}_i)}{h^2}.$$

For variance-calibrated PRSs, the Bayesian prediction accuracy is

$$r^2 = \mathrm{Corr}_{i,\boldsymbol{\beta}|\boldsymbol{D}}^2(g_i, \hat{g}_i) = \frac{\mathrm{Cov}_{i,\boldsymbol{\beta}|\boldsymbol{D}}^2(g_i, \hat{g}_i)}{\mathrm{Var}_{i,\boldsymbol{\beta}|\boldsymbol{D}}(g_i)\mathrm{Var}_{i,\boldsymbol{\beta}|\boldsymbol{D}}(\hat{g}_{ik})} = \frac{\mathrm{Var}_i(\hat{g}_i)}{\mathrm{Var}_{i,\boldsymbol{\beta}|\boldsymbol{D}}(g_i)} = \frac{\mathrm{Var}_i(\hat{g}_i)}{h^2}.$$

Therefore, for PRSs that satisfy both slope and variance calibration, the frequentist and Bayesian definitions of prediction accuracy coincide:

$$r^2 = \kappa^2 = \frac{\mathrm{Var}_i(\hat{g}_i)}{h^2}.$$

**Individual-level misclassification.** Misclassification at the top-$q$ percentile cutoff is defined as disagreement between the PRS-based classification and the true class determined by the latent genetic value $g_i$. Let $t_q = \Phi^{-1}(1-q)$ be the $(1-q)$-quantile of the standard normal distribution. For calibrated PRSs, the variances of $\hat{g}_i$ and $g_i$ across individuals are $h^2 r^2$ and $h^2$, respectively. Under normality assumptions, the corresponding top-$q$ percentile thresholds are $hrt_q$ for $\hat{g}_i$ and $ht_q$ for $g_i$. Thus, the misclassification event for individual $i$ can be represented as

$$\mathcal{M}_i = \{(g_i - ht_q)(\hat{g}_i - hrt_q) < 0\} = \{g_i > ht_q, \hat{g}_i \le hrt_q\} \cup \{g_i \le ht_q, \hat{g}_i > hrt_q\}.$$

The conditional posterior distribution of $g_i$ given $\hat{g}_i$ and $\hat{v}_i$ is $\mathrm{N}(\hat{g}_i, \hat{v}_i)$. Thus, when $\hat{g}_i \le hrt_q$,

$$p_{\text{misclass},i} = \mathrm{P}(\mathcal{M}_i \mid \hat{g}_i, \hat{v}_i) = \mathrm{P}(g_i > ht_q \mid \hat{g}_i, \hat{v}_i) = \Phi\left[(\hat{g}_i - ht_q)/\sqrt{\hat{v}_i}\right],$$

and when $\hat{g}_i > hrt_q$,

$$p_{\text{misclass},i} = \mathrm{P}(\mathcal{M}_i \mid \hat{g}_i, \hat{v}_i) = \mathrm{P}(g_i \le ht_q \mid \hat{g}_i, \hat{v}_i) = 1 - \Phi\left[(\hat{g}_i - ht_q)/\sqrt{\hat{v}_i}\right].$$

This conditional misclassification probability depends on the normalized distance between the point estimate $\hat{g}_i$ and the quantile cutoff $ht_q$. It increases monotonically for $\hat{g}_i < hrt_q$ and decreases monotonically for $\hat{g}_i > hrt_q$, reaching its maximum at the decision threshold $hrt_q$. Unless $r = 1$, the function is discontinuous at this threshold because $\Phi\left[ht_q(r-1)/\sqrt{\hat{v}_i}\right] < 0.5$ and $\Phi\left[ht_q(1-r)/\sqrt{\hat{v}_i}\right] > 0.5$. Noticing that $\mathrm{E}_i(\hat{v}_i) = h^2(1-r^2)$ after calibration, the peak misclassification probability satisfies

$$p_{\max} = \Phi\left[ht_q(1-r)/\sqrt{\hat{v}_i}\right] \approx \Phi\left(t_q\sqrt{(1-r)/(1+r)}\right).$$

As expected, $p_{\max} \to 0.5$ as $r^2 \to 1$ (perfect PRS) and $p_{\max} \to 1-q$ as $r^2 \to 0$ (uninformative PRS).

**Population-level misclassification.** At the population level, assume $(g_i, \hat{g}_i)$ follows a bivariate normal distribution with correlation $r$. The overall misclassification probability at the top-$q$ percentile cutoff can be estimated as:

$$p_{\text{misclass}} = \mathrm{P}(\mathcal{M}_i) = \mathrm{P}(g_i > ht_q, \hat{g}_i \le hrt_q) + \mathrm{P}(g_i \le ht_q, \hat{g}_i > hrt_q)$$

$$= \mathrm{P}(g_i \le ht_q) + \mathrm{P}(\hat{g}_i \le hrt_q) - 2\mathrm{P}(g_i \le ht_q, \hat{g}_i \le hrt_q) = 2\left[\Phi(t_q) - \Phi_2(t_q, t_q; r)\right],$$

where $\Phi(t_q)$ is the CDF of the standard normal distribution evaluated at $t_q$, and $\Phi_2(t_q, t_q; r)$ is the bivariate standard normal CDF with correlation $r$, evaluated at $(t_q, t_q)$. Thus, the population-level misclassification probability is a deterministic function of the PRS prediction accuracy $r$.

The overall positive predictive value (PPV) and negative predictive value (NPV) at the top-*q* percentile cutoff can be analogously estimated as:

$$p_{\text{PPV}} = \mathrm{P}\big(g_i > ht_q \mid \hat{g}_i > hrt_q\big) = \mathrm{P}\big(g_i > ht_q, \hat{g}_i > hrt_q\big)/\mathrm{P}\big(\hat{g}_i > hrt_q\big)$$

$$= 1 - \big[\Phi(t_q) - \Phi_2(t_q, t_q; r)\big]/[1 - \Phi(t_q)] = 1 - \big[\Phi(t_q) - \Phi_2(t_q, t_q; r)\big]/q,$$

$$p_{\text{NPV}} = \mathrm{P}\big(g_i \le ht_q \mid \hat{g}_i \le hrt_q\big) = \mathrm{P}\big(g_i \le ht_q, \hat{g}_i \le hrt_q\big)/\mathrm{P}\big(\hat{g}_i \le hrt_q\big)$$

$$= \Phi_2(t_q, t_q; r)/\Phi(t_q) = \Phi_2(t_q, t_q; r)/(1 - q).$$

Let $\varphi_K(\boldsymbol{u}; \boldsymbol{\Gamma})$ denote the probability density function (PDF) of a $K$-dimensional standard MVN distribution with correlation matrix $\boldsymbol{\Gamma} = [\rho_{kl}]_{K\times K}$. By Plackett's identity (see R.L. Plackett, A reduction formula for normal multivariate integrals, *Biometrika*, 41:351-360, 1954; A. Genz and F. Bretz, Computation of multivariate normal and *t* probabilities, *Springer*, 2009),

$$\frac{\partial}{\partial \rho_{kl}} \varphi_K(\boldsymbol{u}; \boldsymbol{\Gamma}) = \frac{\partial^2}{\partial u_k \partial u_l} \varphi_K(\boldsymbol{u}; \boldsymbol{\Gamma}).$$

For the bivariate case,

$$\frac{p_{\text{misclass}}}{\partial r} = -2\frac{\partial}{\partial r}\Phi_2(t_q, t_q; r) = -2\frac{\partial}{\partial r}\int_{-\infty}^{t_q}\int_{-\infty}^{t_q} \varphi_2(u_1, u_2; r)\,\mathrm{d}u_1\mathrm{d}u_2$$

$$= -2\int_{-\infty}^{t_q}\int_{-\infty}^{t_q} \frac{\partial}{\partial r}\varphi_2(u_1, u_2; r)\,\mathrm{d}u_1\mathrm{d}u_2 = -2\int_{-\infty}^{t_q}\int_{-\infty}^{t_q} \frac{\partial^2}{\partial u_1 \partial u_2}\varphi_2(u_1, u_2; r)\,du_1\mathrm{d}u_2$$

$$= -2\varphi_2(t_q, t_q; r) < 0,$$

Therefore, population-level misclassification probability $p_{\text{misclass}}$ decreases monotonically as PRS accuracy increases.

For calibrated PRSs, $r = \sqrt{1 - \mathrm{E}(\hat{v}_i)/h^2}$, thus

$$\frac{\partial r}{\partial \mathrm{E}(\hat{v}_i)} = -\frac{1}{2h^2 r} < 0.$$

By the chain rule,

$$\frac{p_{\text{misclass}}}{\partial \text{E}(\hat{v}_i)} = \frac{\partial p_{\text{misclass}}}{\partial r}\frac{\partial r}{\partial \text{E}(\hat{v}_i)} = \frac{\varphi_2(q, q; r)}{h^2 r} > 0.$$

Therefore, population-level misclassification probability $p_{\text{misclass}}$ decreases monotonically as posterior uncertainty in the PRS declines.

By the law of total expectation,

$$p_{\text{misclass}} = \text{P}(\mathcal{M}_i) = \text{E}(1_{\mathcal{M}_i}) = \text{E}_i[\text{E}(1_{\mathcal{M}_i} \mid \hat{g}_i, \hat{v}_i)] = \text{E}_i[\text{P}(\mathcal{M}_i \mid \hat{g}_i, \hat{v}_i)] = \text{E}_i(p_{\text{misclass},i}).$$

Therefore, when the joint distribution of $g_i$ and $\hat{g}_i$ is well approximated under normality assumptions, the population-level misclassification probability is closely approximately by the average of the individual-level conditional misclassification probabilities.

**Misclassification of the phenotype.** Individual-level and population-level misclassification probabilities can be similarly calculated for the observed phenotype.

At the individual-level, for quantitative traits, misclassification at the top-$q$ percentile cutoff is defined by comparing the PRS-based classification with whether the observed phenotype $y_i$ exceeds its top-$q$ threshold $t_q$. Under calibrated PRSs and normality assumptions, $y_i \mid \hat{g}_i, \hat{v}_i \sim \text{N}(\hat{g}_i, \hat{v}_i + 1 - \hat{h}^2)$.

Therefore, when $\hat{g}_i \leq hrt_q$,

$$p^y_{\text{misclass},i} = \text{P}(y_i > t_q \mid \hat{g}_i, \hat{v}_i) = \Phi[(\hat{g}_i - t_q)/(\hat{v}_i + 1 - \hat{h}^2)^{1/2}],$$

and when $\hat{g}_i > hrt_q$,

$$p^y_{\text{misclass},i} = \text{P}(y_i \leq t_q \mid \hat{g}_i, \hat{v}_i) = 1 - \Phi[(\hat{g}_i - t_q)/(\hat{v}_i + 1 - \hat{h}^2)^{1/2}].$$

For binary phenotypes, misclassification is defined as disagreement between the PRS-based classification and the observed label $y_i$. Under the liability threshold model, $l_i \mid \hat{g}_i, \hat{v}_i \sim \text{N}(\hat{g}_i, \hat{v}_i + 1 - \hat{h}^2)$.

Therefore, when $\hat{g}_i \leq hrt_q$,

$$p^y_{\text{misclass},i} = \text{P}(y_i = 1 \mid \hat{g}_i, \hat{v}_i) = \text{P}(l_i > \tau \mid \hat{g}_i, \hat{v}_i) = \Phi[(\hat{g}_i - \tau)/(\hat{v}_i + 1 - \hat{h}^2)^{1/2}],$$

and when $\hat{g}_i > hrt_q$,

$$p^y_{\text{misclass},i} = \text{P}(y_i = 0 \mid \hat{g}_i, \hat{v}_i) = \text{P}(l_i \leq \tau \mid \hat{g}_i, \hat{v}_i) = 1 - \Phi[(\hat{g}_i - \tau)/(\hat{v}_i + 1 - \hat{h}^2)^{1/2}].$$

At the population-level, for quantitative traits, assume $(y_i, \hat{g}_i)$ follows a bivariate normal distribution with correlation $hr$. The overall misclassification probability at the top-$q$ percentile cutoff can be estimated as:

$$p^y_{\text{misclass}} = \mathrm{P}(y_i > t_q, \hat{g}_i \le hrt_q) + \mathrm{P}(y_i \le t_q, \hat{g}_i > hrt_q)$$

$$= \mathrm{P}(y_i \le t_q) + \mathrm{P}(\hat{g}_i \le hrt_q) - 2\mathrm{P}(y_i \le t_q, \hat{g}_i \le hrt_q) = 2[\Phi(t_q) - \Phi_2(t_q, t_q; hr)].$$

For binary phenotypes, under the liability threshold model, assume $(l_i, \hat{g}_i)$ follows a bivariate normal distribution with correlation $hr$. The overall misclassification probability can be estimated as:

$$p^y_{\text{misclass}} = \mathrm{P}(y_i = 1, \hat{g}_i \le hrt_q) + \mathrm{P}(y_i = 0, \hat{g}_i > hrt_q)$$

$$= \mathrm{P}(l_i > \tau, \hat{g}_i \le hrt_q) + \mathrm{P}(l_i \le \tau, \hat{g}_i > hrt_q)$$

$$= \mathrm{P}(l_i \le \tau) + \mathrm{P}(\hat{g}_i \le hrt_q) - 2\mathrm{P}(l_i \le \tau, \hat{g}_i \le hrt_q)$$

$$= \Phi(\tau) + \Phi(t_q) - 2\Phi_2(\tau, t_q; hr).$$

**With-truth classification inconsistency.** Consider $K$ slope- and variance-calibrated PRSs, with point estimates and posterior variances for individual $i$ denoted by $(\hat{g}_{ik}, \hat{v}_{ik}), k = 1, 2, \cdots, K$, and corresponding prediction accuracies $\boldsymbol{r} = (r_1, r_2, \cdots, r_K)^T$. Let the joint correlation matrix between the true genetic value $g_i$ and the $K$ PRSs be

$$\boldsymbol{\Sigma} = \begin{bmatrix} 1 & \boldsymbol{r}^T \\ \boldsymbol{r} & \boldsymbol{C} \end{bmatrix},$$

where $\boldsymbol{C} = [c_{kl}]_{K \times K}$ is the correlation matrix among the PRSs.

The with-truth inconsistency, $\mathcal{I}_{\text{PRS+},i}$, compares each PRS-based classification with the true class determined by $g_i$ and is defined as any instance in which at least two classifications among $\{g_i, \hat{g}_{i1}, \cdots, \hat{g}_{iK}\}$ disagree at the top-*q* percentile cutoff. This is equivalent to requiring that at least one PRS misclassifies individual $i$:

$$\mathcal{I}_{\text{PRS+},i} = \cup_{k=1}^{K} \mathcal{M}_{ik},$$

where $\mathcal{M}_{ik}$ is the misclassification event for the $k$-th PRS. Thus, the probability of with-truth inconsistency increases as the misclassification probabilities of individual PRSs increase.

Assuming that $(g_i, \hat{g}_{i1}, \cdots, \hat{g}_{iK})$ follows a MVN distribution with correlation matrix $\boldsymbol{\Sigma}$, the overall probability of with-truth inconsistency at the top-*q* percentile cutoff is

$$\mathrm{P}(\mathcal{I}_{\text{PRS+},i}) = 1 - \mathrm{P}(g_i \le ht_q, \hat{g}_{ik} \le hr_k t_q \text{ for all } k) - \mathrm{P}(g_i > ht_q, \hat{g}_{ik} > hr_k t_q \text{ for all } k)$$

$$= 1 - \Phi_{K+1}(t_q \mathbf{1}_{K+1}; \boldsymbol{\Sigma}) - \Phi_{K+1}(-t_q \mathbf{1}_{K+1}; \boldsymbol{\Sigma}),$$

Where $\Phi_{K+1}(t_q \mathbf{1}_{K+1}; \boldsymbol{\Sigma})$ is the CDF of a $(K+1)$-dimensional standard MVN distribution with correlation matrix $\boldsymbol{\Sigma}$, evaluated at the vector $t_q \mathbf{1}_{K+1}$, and $\mathbf{1}_{K+1}$ is a $(K+1)$-dimensional vector of ones. Thus, the probability of with-truth inconsistency is jointly determined by the accuracies of the PRSs and their pairwise correlations.

By Plackett's identity,

$$\frac{\partial}{\partial c_{kl}}\Phi_{K+1}\left(t_q\mathbf{1}_{K+1};\mathbf{\Sigma}\right) = \frac{\partial}{\partial c_{kl}}\int_{-\infty}^{t_q}\int_{-\infty}^{t_q}\cdots\int_{-\infty}^{t_q}\varphi_{K+1}(\boldsymbol{u};\mathbf{\Sigma})\mathrm{d}\boldsymbol{u} = \int_{-\infty}^{t_q}\int_{-\infty}^{t_q}\cdots\int_{-\infty}^{t_q}\frac{\partial}{\partial c_{kl}}\varphi_{K+1}(\boldsymbol{u};\mathbf{\Sigma})\mathrm{d}\boldsymbol{u}$$

$$= \int_{-\infty}^{t_q}\int_{-\infty}^{t_q}\cdots\int_{-\infty}^{t_q}\frac{\partial^2}{\partial u_k \partial u_l}\varphi_{K+1}(\boldsymbol{u};\mathbf{\Sigma})\mathrm{d}\boldsymbol{u}$$

$$= \int_{-\infty}^{t_q}\int_{-\infty}^{t_q}\cdots\int_{-\infty}^{t_q}\varphi_{K+1}\left(u_k = t_q, u_l = t_q, \boldsymbol{u}_{-kl};\mathbf{\Sigma}\right)\mathrm{d}\boldsymbol{u}_{-kl} > 0,$$

where $\boldsymbol{u} = (u_0, u_1, \cdots, u_K)^T$ is a $(K+1)$-dimension vector, and $\boldsymbol{u}_{-kl}$ denotes the remaining $K-1$ components excluding $u_k$ and $u_l$. The same argument shows that the derivative of $\Phi_{K+1}\left(-t_q\mathbf{1}_{K+1};\mathbf{\Sigma}\right)$ with respect to $c_{kl}$ is positive. Consequently,

$$\frac{\partial}{\partial c_{kl}}\mathrm{P}\left(\mathcal{I}_{\mathrm{PRS}+,i}\right) = -\frac{\partial}{\partial c_{kl}}\Phi_{K+1}\left(t_q\mathbf{1}_{K+1};\mathbf{\Sigma}\right) - \frac{\partial}{\partial c_{kl}}\Phi_{K+1}\left(-t_q\mathbf{1}_{K+1};\mathbf{\Sigma}\right) < 0.$$

Similarly, applying Plackett's identity to derivatives with respect to any accuracy parameter $r_k$ yields

$$\frac{\partial}{\partial r_k}\mathrm{P}\left(\mathcal{I}_{\mathrm{PRS}+,i}\right) = -\frac{\partial}{\partial r_k}\Phi_{K+1}\left(t_q\mathbf{1}_{K+1};\mathbf{\Sigma}\right) - \frac{\partial}{\partial r_k}\Phi_{K+1}\left(-t_q\mathbf{1}_{K+1};\mathbf{\Sigma}\right) < 0.$$

Therefore, the probability of with-truth classification inconsistency $\mathrm{P}\left(\mathcal{I}_{\mathrm{PRS}+,i}\right)$ decreases monotonically as (i) the accuracy of any PRS increases (and equivalently, as any PRS uncertainty decreases), and (ii) the correlation between any pair of PRSs increases.

**PRS-only classification inconsistency.** The PRS-only inconsistency, $\mathcal{I}_{\mathrm{PRS},i}$, evaluates agreement among the $K$ PRSs alone, without reference to the true genetic value $g_i$. It is defined as any instance in which at least two PRSs disagree at the top-$q$ percentile cutoff. This is equivalent to requiring that at least one PRS misclassifies individual $i$, excluding scenarios in which all PRS-based classifications are simultaneously incorrect:

$$\mathcal{I}_{\mathrm{PRS},i} = \cup_{k=1}^{K}\mathcal{M}_{ik} / \cap_{k=1}^{K}\mathcal{M}_{ik}.$$

Under the MVN assumption, the probability of PRS-only inconsistency at the top-$q$ percentile cutoff is

$$\mathrm{P}\left(\mathcal{I}_{\mathrm{PRS},i}\right) = 1 - \mathrm{P}\left(\hat{g}_{ik} \le hr_k t_q \text{ for all } k\right) - \mathrm{P}\left(\hat{g}_{ik} > hr_k t_q \text{ for all } k\right)$$

$$= 1 - \Phi_K\left(t_q\mathbf{1}_K;\boldsymbol{C}\right) - \Phi_K\left(-t_q\mathbf{1}_K;\boldsymbol{C}\right).$$

Thus, the PRS-only inconsistency probability depends only on the correlation structure $\boldsymbol{C}$ among the PRSs.

A similar argument using Plackett's identity shows that

$$\frac{\partial}{\partial c_{kl}} \mathrm{P}(\mathcal{I}_{\mathrm{PRS},i}) = -\frac{\partial}{\partial c_{kl}} \Phi_K(t_q \mathbf{1}_K; \boldsymbol{C}) - \frac{\partial}{\partial c_{kl}} \Phi_K(-t_q \mathbf{1}_K; \boldsymbol{C}) < 0.$$

Therefore, the probability of PRS-only inconsistency, $\mathrm{P}(\mathcal{I}_{\mathrm{PRS},i})$, decreases monotonically as the correlation between any pair of PRSs increases.

By definition,

$$\mathrm{P}(\mathcal{I}_{\mathrm{PRS}+,i}) - \mathrm{P}(\mathcal{I}_{\mathrm{PRS},i}) = \mathrm{P}(\cap_{k=1}^{K} \mathcal{M}_{ik}),$$

which implies that $\mathrm{P}(\mathcal{I}_{\mathrm{PRS},i})$ is always smaller than $\mathrm{P}(\mathcal{I}_{\mathrm{PRS}+,i})$. The difference arises from scenarios in which all PRS-based classifications are simultaneously incorrect – events that are treated as consistent under the PRS-only definition but inconsistent when the true latent class is taken into account. As PRS accuracies and their correlations increase, the probability that all classifications are simultaneously incorrect decreases, and the two inconsistency measures tend to converge.

**Probability of classification profiles.** The MVN-based framework enables computation of the probability of any joint classification profile across the PRSs. Throughout, we assume that the $K$ PRSs follow a MVN distribution with zero mean and correlation matrix $\boldsymbol{C}$.

The probability that all PRSs exceed the top-$q$ percentile cutoff can be estimated as

$$p_{\mathrm{all-high}} = \mathrm{P}(\hat{g}_{ik} > hr_k t_q \text{ for all } k) = \Phi_K(-t_q \mathbf{1}_K; \boldsymbol{C}).$$

The probability that all PRSs fall below the top-$q$ percentile cutoff can be estimated as

$$p_{\mathrm{all-low}} = \mathrm{P}(\hat{g}_{ik} \le hr_k t_q \text{ for all } k) = \Phi_K(t_q \mathbf{1}_K; \boldsymbol{C}).$$

The probability that at least one PRS exceeds the top-$q$ percentile cutoff can be estimated as

$$p_{\mathrm{any-high}} = 1 - \mathrm{P}(\hat{g}_{ik} \le hr_k t_q \text{ for all } k) = 1 - \Phi_K(t_q \mathbf{1}_K; \boldsymbol{C}).$$

Finally, the probability of an extreme inconsistency event where at least one PRS exceeds the top-$q$ percentile cutoff and at least one PRS falls below the bottom-$q$ percentile cutoff can be estimated as

$$\begin{aligned} p_{\mathrm{extreme}} &= 1 - \mathrm{P}(\hat{g}_{ik} > -hr_k t_q \text{ for all } k) - \mathrm{P}(\hat{g}_{ik} \le hr_k t_q \text{ for all } k) + \mathrm{P}(-hr_k t_q < \hat{g}_{ik} \le hr_k t_q \text{ for all } k) \\ &= 1 - 2\Phi_K(t_q \mathbf{1}_K; \boldsymbol{C}) + \Phi_K([-t_q \mathbf{1}_K, t_q \mathbf{1}_K]; \boldsymbol{C}), \end{aligned}$$

where $\Phi_K([-t_q \mathbf{1}_K, t_q \mathbf{1}_K]; \boldsymbol{C})$ denotes the integral of the $K$-dimensional standard MVN distribution with correlation matrix $\boldsymbol{C}$ over the hyper-rectangle $[-t_q, t_q]^K$.

**Constraints between accuracy and correlation.** For slope- and variance-calibrated PRSs, consider the correlation matrix of $(g_i, \hat{g}_{ik}, \hat{g}_{il})$:

$$\boldsymbol{R}_{kl} = \begin{pmatrix} 1 & r_k & r_l \\ r_k & 1 & c_{kl} \\ r_l & c_{kl} & 1 \end{pmatrix}.$$

Since $\boldsymbol{R}_{kl}$ must be positive semidefinite, its determinant must satisfy $\det(\boldsymbol{R}_{kl}) \geq 0$. Evaluating the determinant yields the constraint:

$$r_k r_l - \sqrt{(1 - r_k^2)(1 - r_l^2)} \leq c_{kl} \leq r_k r_l + \sqrt{(1 - r_k^2)(1 - r_l^2)}.$$

If $c_{kl} = 0$, the inequality reduces to $r_k^2 + r_l^2 \leq 1$, implying that two highly accurate PRSs cannot be mutually uncorrelated. Conversely, if both PRSs are near-perfect ($r_k \approx r_l \approx 1$), then $c_{kl} \approx 1$, indicating that strong PRSs must be highly correlated. More generally, as either $r_k$ or $r_l$ increase, the feasible range of $c_{kl}$ tightens, constraining the degree of independence between accurate PRSs. Thus, the prediction accuracy of individual PRSs and their pairwise correlations cannot vary independently; they are intrinsically linked. Improving accuracy necessarily reduces the allowed range of correlations, and highly accurate PRSs must exhibit strong mutual dependence.

**Approximation of posterior covariance matrix.** Given any weight vector $\widehat{\boldsymbol{\omega}} = [\widehat{\omega}_1, \widehat{\omega}_2, \cdots, \widehat{\omega}_K]^T$ used to combine $K$ PRSs, with individual-level point estimates $\widehat{\boldsymbol{g}}_i = (\hat{g}_{i1}, \hat{g}_{i2}, \cdots, \hat{g}_{iK})^T$, the integrated score for individual $i$ is $\widehat{\boldsymbol{\omega}}^T \widehat{\boldsymbol{g}}_i$. Estimating the posterior variance of this integrated score requires the individual-level posterior covariance matrix of the PRSs, $\widehat{\boldsymbol{S}}_i = [\hat{S}_{i,kl}]_{K \times K}$, which is rarely available in practice as PRSs are typically trained separately. To approximate $\widehat{\boldsymbol{S}}_i$, consider the individual-level error model:

$$g_i = \hat{g}_{ik} + \varepsilon_{ik}, \quad k = 1, 2, \cdots, K,$$

where $\varepsilon_{ik}$ is an error term with posterior mean zero and posterior variance $\hat{v}_{ik}$. By definition,

$$\hat{S}_{i,kl} = \mathrm{Cov}_{\boldsymbol{\beta}|\boldsymbol{D}}[\varepsilon_{ik}, \varepsilon_{il} | \boldsymbol{x}_i].$$

By the law of total covariance and the fact that $\mathrm{E}_{\boldsymbol{\beta}|\boldsymbol{D}}(\varepsilon_{ik} | \boldsymbol{x}_i) = \mathrm{E}_{\boldsymbol{\beta}|\boldsymbol{D}}(\varepsilon_{il} | \boldsymbol{x}_i) = 0$,

$$\mathrm{Cov}_{i,\boldsymbol{\beta}|\boldsymbol{D}}(\varepsilon_{ik}, \varepsilon_{il}) = \mathrm{E}_i[\mathrm{Cov}_{\boldsymbol{\beta}|\boldsymbol{D}}(\varepsilon_{ik}, \varepsilon_{il} | \boldsymbol{x}_i)] + \mathrm{Cov}_i\left[\mathrm{E}_{\boldsymbol{\beta}|\boldsymbol{D}}(\varepsilon_{ik} | \boldsymbol{x}_i), \mathrm{E}_{\boldsymbol{\beta}|\boldsymbol{D}}(\varepsilon_{il} | \boldsymbol{x}_i)\right] = \mathrm{E}_i\left(\hat{S}_{i,kl}\right).$$

One the other hand, for slope- and variance-calibrated PRSs,

$$\begin{aligned} \mathrm{Cov}_{i,\boldsymbol{\beta}|\boldsymbol{D}}(\varepsilon_{ik}, \varepsilon_{il}) &= \mathrm{Cov}_{i,\boldsymbol{\beta}|\boldsymbol{D}}(g_i - \hat{g}_{ik}, g_i - \hat{g}_{il}) \\ &= \mathrm{Var}_{i,\boldsymbol{\beta}|\boldsymbol{D}}(g_i) - \mathrm{Cov}_{i,\boldsymbol{\beta}|\boldsymbol{D}}(g_i, \hat{g}_{ik}) - \mathrm{Cov}_{i,\boldsymbol{\beta}|\boldsymbol{D}}(g_i, \hat{g}_{il}) + \mathrm{Cov}_{i,\boldsymbol{\beta}|\boldsymbol{D}}(\hat{g}_{ik}, \hat{g}_{il}) \\ &= \mathrm{Var}_{i,\boldsymbol{\beta}|\boldsymbol{D}}(g_i) - \mathrm{Var}_i(\hat{g}_{ik}) - \mathrm{Var}_i(\hat{g}_{il}) + \mathrm{Cov}_i(\hat{g}_{ik}, \hat{g}_{il}) = h^2(1 - r_k^2 - r_l^2 + c_{kl} r_k r_l). \end{aligned}$$

Similarly,

$$\mathrm{E}_i\left(\hat{S}_{i,kk}\right) = \mathrm{Var}_{i,\boldsymbol{\beta}|\boldsymbol{D}}(\varepsilon_{ik}) = \mathrm{E}_i(\hat{v}_{ik}) = h^2(1 - r_k^2).$$

Therefore, the posterior correlation matrix of the errors, denoted as $\boldsymbol{\Omega} = [\Omega_{kl}]_{K\times K}$, can be approximated using PRS accuracies and their pairwise correlations:

$$\widehat{\Omega}_{kl} \approx \frac{\mathrm{E}_i(\hat{S}_{i,kl})}{\sqrt{\mathrm{E}_i(\hat{S}_{i,kk})\mathrm{E}_i(\hat{S}_{i,ll})}} = \frac{1 - r_k^2 - r_l^2 + c_{kl} r_k r_l}{\sqrt{(1 - r_k^2)(1 - r_l^2)}}.$$

The individual-level posterior covariance matrix of the $K$ PRSs can then be approximated by rescaling this correlation matrix using the posterior variances:

$$\widehat{\boldsymbol{S}}_i \approx \widehat{\boldsymbol{V}}_i^{1/2} \widehat{\boldsymbol{\Omega}} \widehat{\boldsymbol{V}}_i^{1/2},$$

where $\widehat{\boldsymbol{V}}_i = \mathrm{diag}\{\hat{v}_{i1}, \hat{v}_{i2}, \cdots, \hat{v}_{iK}\}$ is a diagonal matrix.

**PRS integration.** Given the approximate posterior covariance matrix of the PRSs, the integrated PRS for individual $i$ is defined by the weighted linear combination $\hat{g}_i^{\mathrm{int}} = \widehat{\boldsymbol{\omega}}^T \widehat{\boldsymbol{g}}_i$, with posterior variance $\hat{v}_i^{\mathrm{int}} = \widehat{\boldsymbol{\omega}}^T \widehat{\boldsymbol{S}}_i \widehat{\boldsymbol{\omega}}$. Assuming the posterior distribution of $g_i$ is approximately normal, and that $(\hat{g}_i^{\mathrm{int}}, \hat{v}_i^{\mathrm{int}})$ has been slope- and variance-calibrated, an individual-specific $(1 - \alpha)$-level confidence interval (CI) for $g_i$ can be constructed as:

$$\mathrm{CI}_{1-\alpha}(g_i): \qquad \hat{g}_i^{\mathrm{int}} \pm z_{1-\alpha/2} (\hat{v}_i^{\mathrm{int}})^{1/2}.$$

**Uncertainty-aware classification.** The probabilistic uncertainty-aware stratification method ranks and selects individuals based on the posterior probability that their genetic value exceeds a prespecified top-$q$ percentile threshold, which incorporates both the PRS point estimate $\hat{g}_i$ and its individual-specific posterior uncertainty $\hat{v}_i$:

$$p_{i,\mathrm{pst}} = \Phi\left[(\hat{g}_i - ht_q)/\sqrt{\hat{v}_i}\right].$$

Under 0-1 loss, the Bayes-optimal decision rule classifies individual $i$ as high risk if $p_{i,\mathrm{pst}} > 0.5$ and as non-high risk otherwise. Since $\Phi(\cdot)$ is monotone increasing, this rule is equivalent to selecting individuals satisfying $\hat{g}_i > ht_q$. Thus, the proportion of individuals classified as high risk is

$$p_{\mathrm{high}} = \mathrm{P}\left(\hat{g}_i > ht_q\right).$$

For slope- and variance-calibrated PRSs, $\hat{g}_i$ has zero mean and variance $h^2 r^2$ across individuals. Under the normality assumption,

$$p_{\mathrm{high}} = 1 - \mathrm{P}\left(\hat{g}_i \le ht_q\right) = 1 - \Phi\left(\frac{t_q}{r}\right) = 1 - \Phi\left(\frac{\Phi^{-1}(1 - q)}{r}\right).$$

As expected, if the PRS is perfect ($r^2 \to 1$), then $p_{\text{high}} \to q$, indicating that the PRS recovers exactly the top $q\%$ of individuals. If the PRS is uninformative ($r^2 \to 0$), then $p_{\text{high}} \to 0$, because the posterior probability never exceeds 0.5, and the Bayes-optimal rule classifies no one as high risk.

More generally, if a posterior probability threshold $\tau$ is used to classify individuals, the decision rule is

$$\delta(\hat{g}_i, \hat{v}_i) = 1\{\mathrm{P}(g_i > ht_q \mid \boldsymbol{D}) > \tau\},$$

where $\tau = 0.5$ yields the Bayes-optimal classifier under 0-1 loss, $\tau < 0.5$ corresponds to a more liberal classification rule, and $\tau > 0.5$ corresponds to a more conservative classification rule. Recall that for calibrated PRSs, under the normality assumption, $g_i \mid \boldsymbol{D} \sim \mathrm{N}(\hat{g}_i, \hat{v}_i)$. Therefore,

$$\delta(\hat{g}_i, \hat{v}_i) = 1\{\mathrm{P}(g_i > ht_q \mid \boldsymbol{D}) > \tau\} = 1\left\{\Phi\left(\frac{\hat{g}_i - ht_q}{\sqrt{\hat{v}_i}}\right) > \tau\right\}$$

$$= 1\left\{\frac{\hat{g}_i - ht_q}{\sqrt{\hat{v}_i}} > z_\tau\right\} = 1\{\hat{g}_i > ht_q + z_\tau\sqrt{\hat{v}_i}\},$$

where $z_\tau$ is the $\tau$-quantile of the standard normal distribution. Thus, the individual-specific uncertainty-aware decision threshold is $ht_q + z_\tau\sqrt{\hat{v}_i}$, which increases with $\hat{v}_i$. This indicates that individuals with greater posterior uncertainty require a larger point estimate $\hat{g}_i$ to exceed the posterior probability threshold $\tau$ and be classified as high risk.

### Supplementary Figures

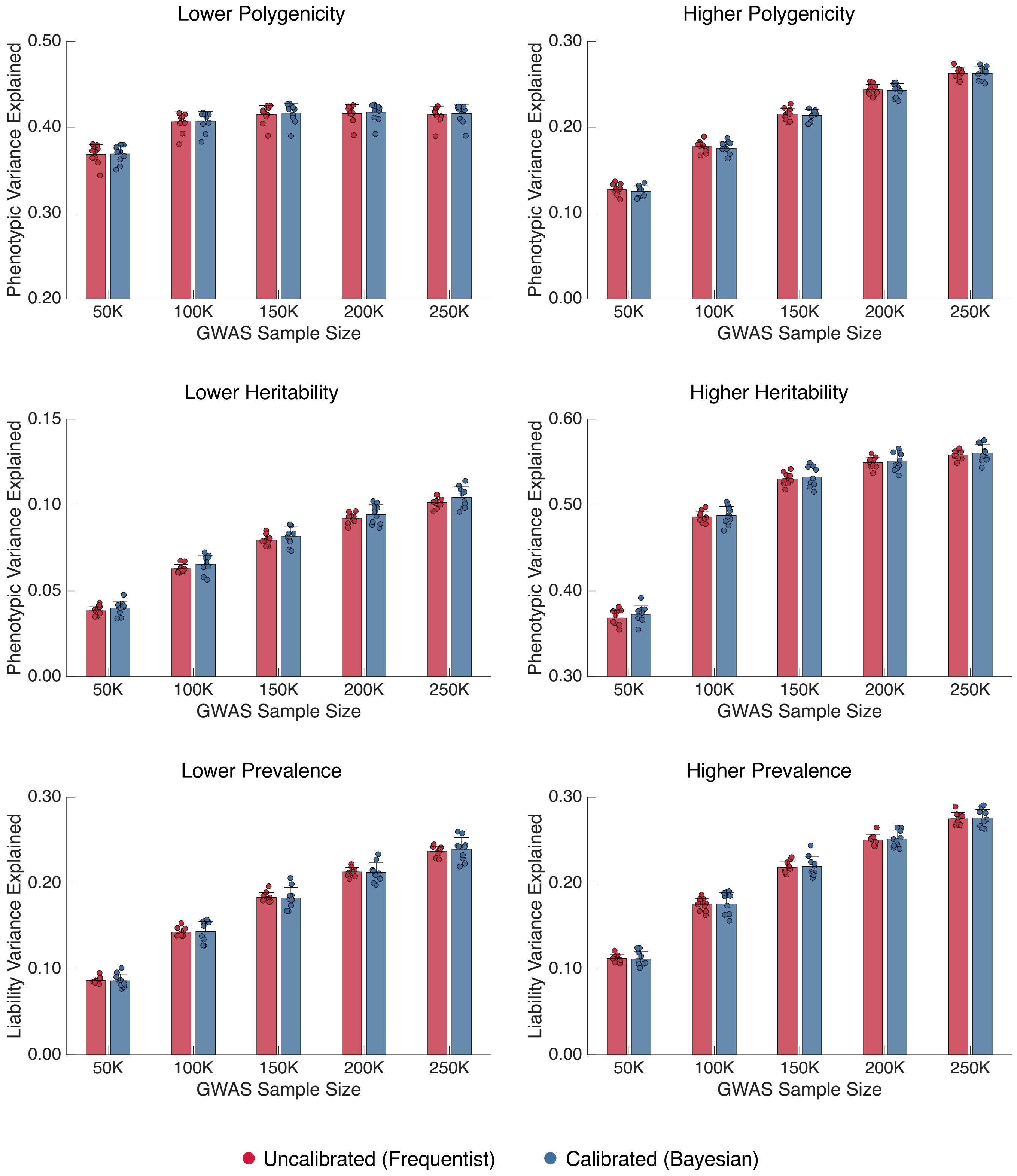


**Supplementary Figure 1:** The prediction accuracy for uncalibrated PRSs (frequentist definition) and calibrated PRSs (Bayesian definition) across simulation settings and GWAS training sample sizes. Bars and error bars indicate the mean and standard deviation across 10 simulation replicates, with individual estimates overlaid.

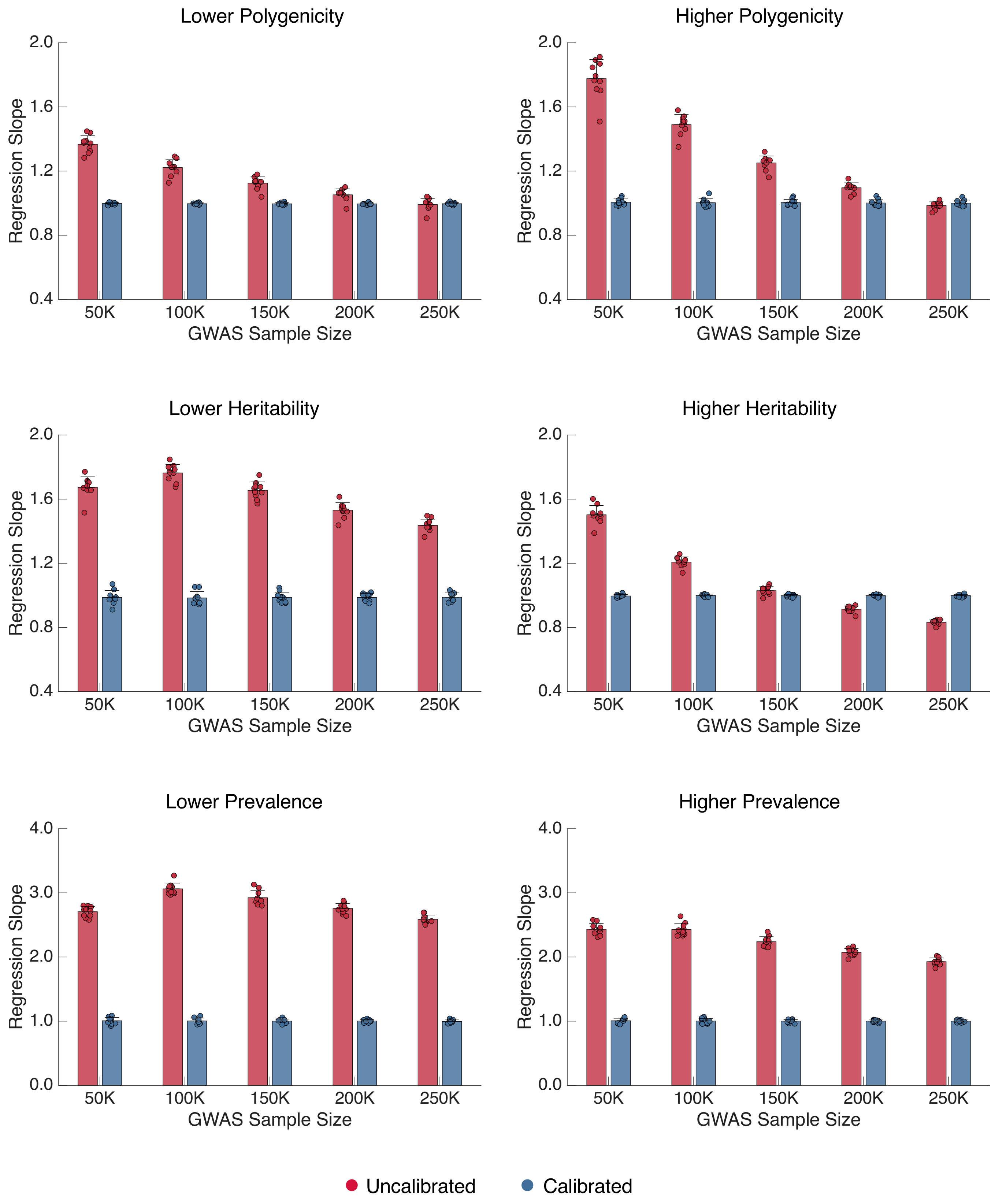


**Supplementary Figure 2:** The regression slope of the true genetic value $g_i$ on the calibrated and uncalibrated posterior point estimate $\hat{g}_i$ in the testing dataset across simulation settings and GWAS training sample sizes. Bars and error bars indicate the mean and standard deviation across 10 simulation replicates, with individual estimates overlaid.

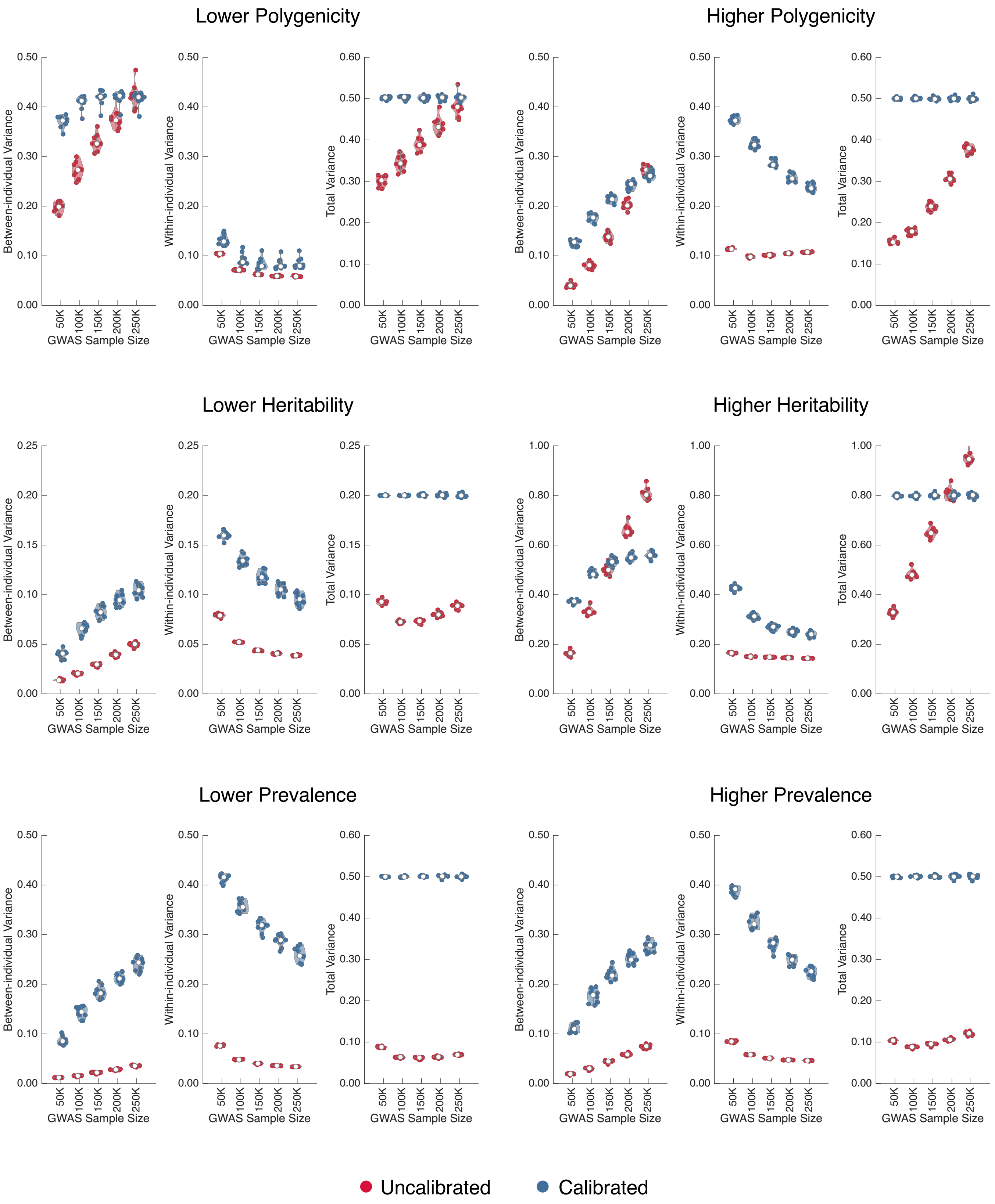


**Supplementary Figure 3:** Between-individual variance of the PRS point estimate $\mathrm{Var}_i(\hat{g}_i)$ (left panel), average within-individual posterior variance $\mathrm{E}_i(\hat{v}_i)$ (middle panel), and total posterior predictive variance (right panel) for calibrated and uncalibrated PRSs in the testing dataset across simulation settings and GWAS training sample sizes. Each violin plot shows the median across 10 simulation replicates, with individual estimates overlaid.

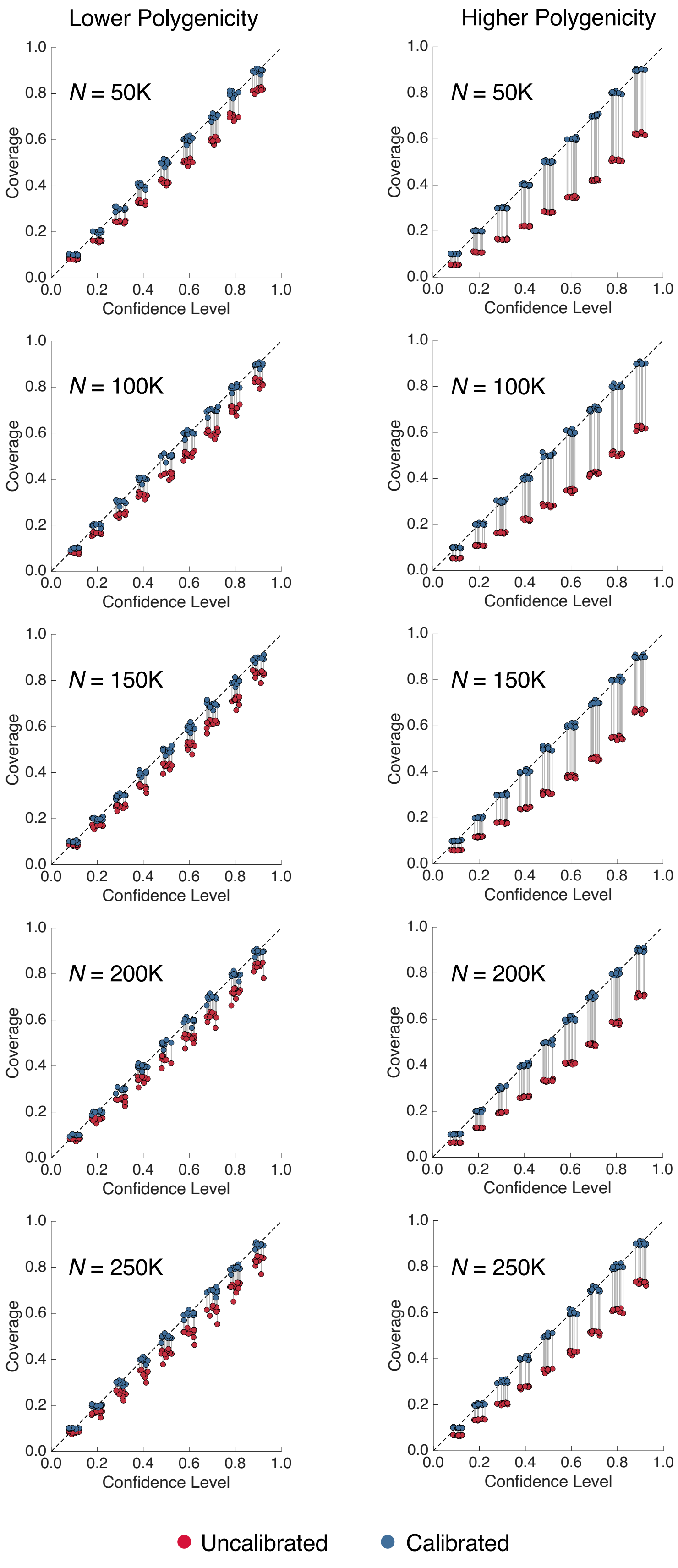


**Supplementary Figure 4:** The coverage (the proportion of confidence intervals that contain the true genetic value) of calibrated and uncalibrated PRSs in the testing dataset across confidence levels and GWAS training sample sizes, under different polygenicity of the genetic architecture. Each dot represents a simulation replicate, with slight horizontal jitter for visualization.

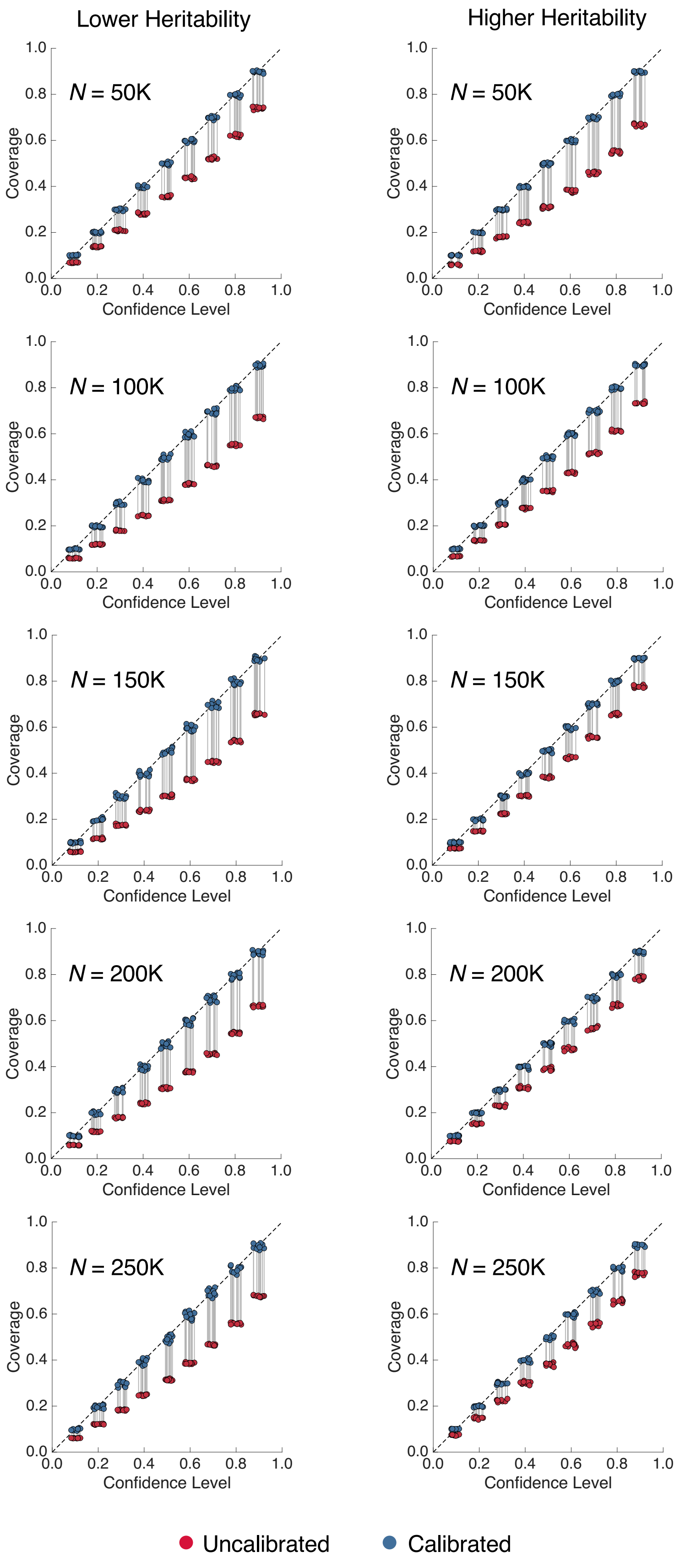


**Supplementary Figure 5:** The coverage (the proportion of confidence intervals that contain the true genetic value) of calibrated and uncalibrated PRSs in the testing dataset across confidence levels and GWAS training sample sizes, under different SNP heritability. Each dot represents a simulation replicate, with slight horizontal jitter for visualization.

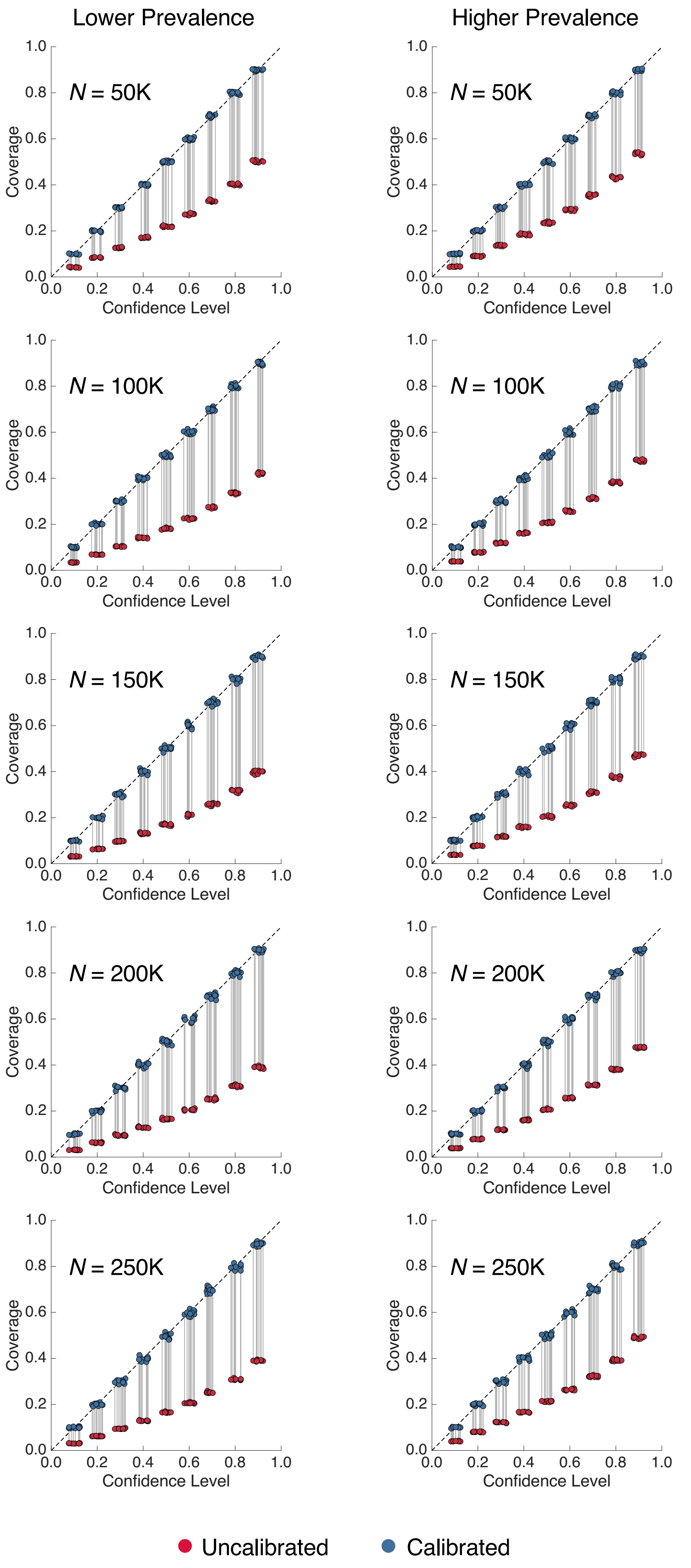


**Supplementary Figure 6:** The coverage (the proportion of confidence intervals that contain the true genetic value) of calibrated and uncalibrated PRSs in the testing dataset across confidence levels and GWAS training sample sizes, under different prevalences of the binary phenotype. Each dot represents a simulation replicate, with slight horizontal jitter for visualization.

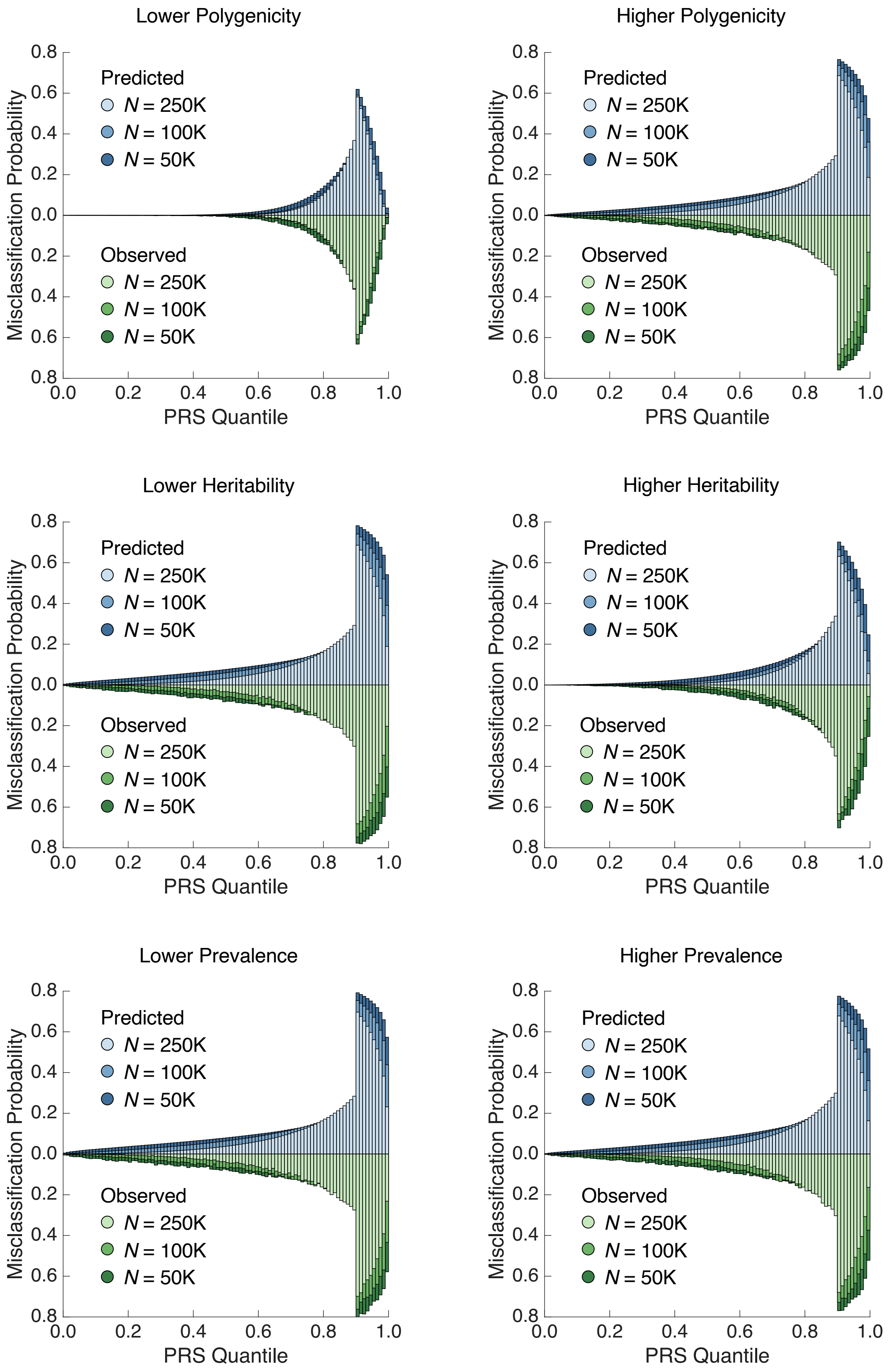


**Supplementary Figure 7:** The distribution of predicted (upper panel) and empirically observed (lower panel) conditional (individual-level) misclassification probabilities across PRS percentiles in the testing dataset for three GWAS training sample sizes (50K, 100K and 250K) across simulation settings, using the top 10% as the risk threshold.

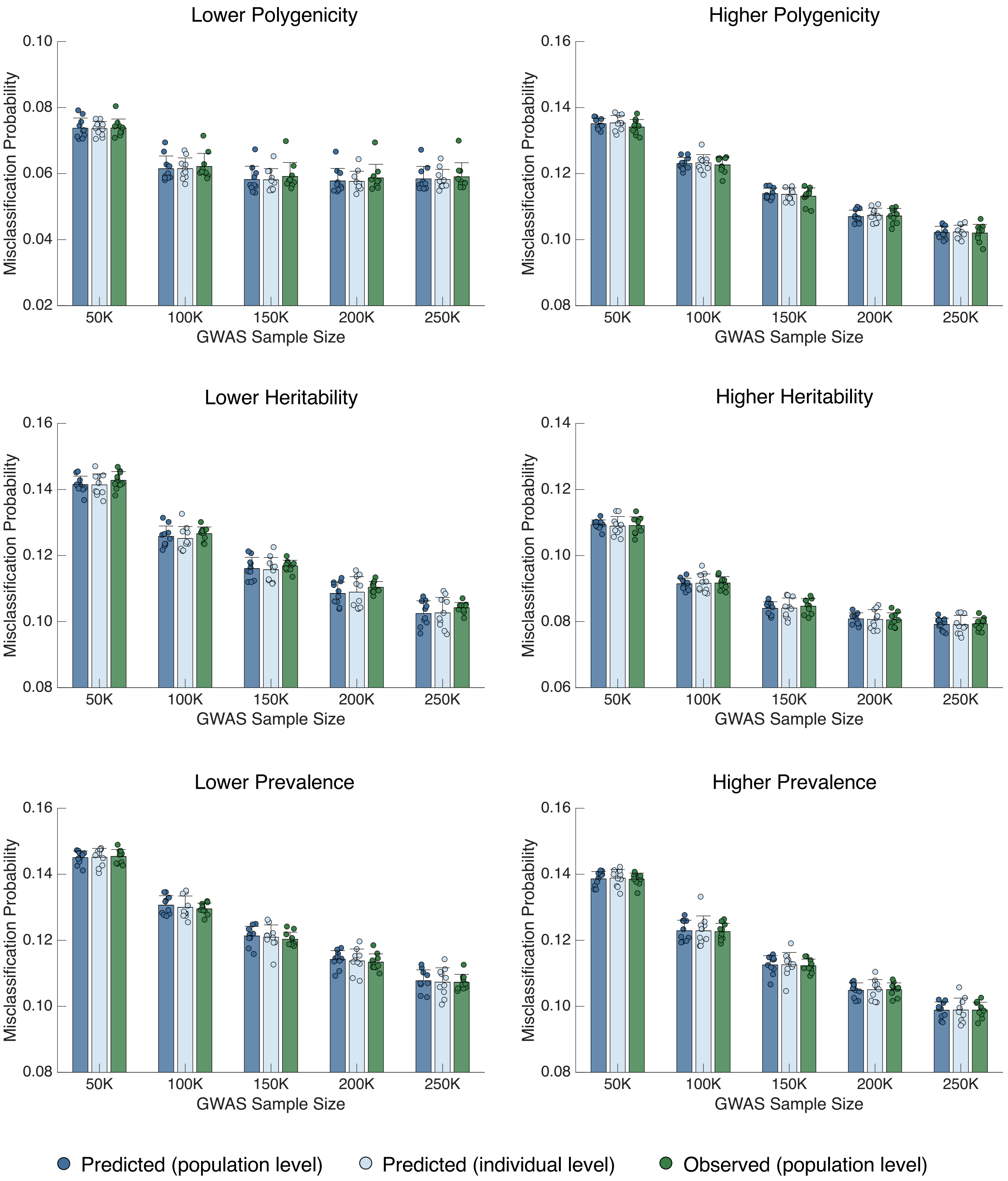


**Supplementary Figure 8:** Comparison of (i) the predicted population-level misclassification probability, (ii) the average predicted individual-level conditional misclassification probability, and (iii) the empirically observed overall misclassification rate in the testing dataset across simulation settings and GWAS training sample sizes, using the top 10% as the risk threshold. Bars and error bars indicate the mean and standard deviation across 10 simulation replicates, with individual estimates overlaid.

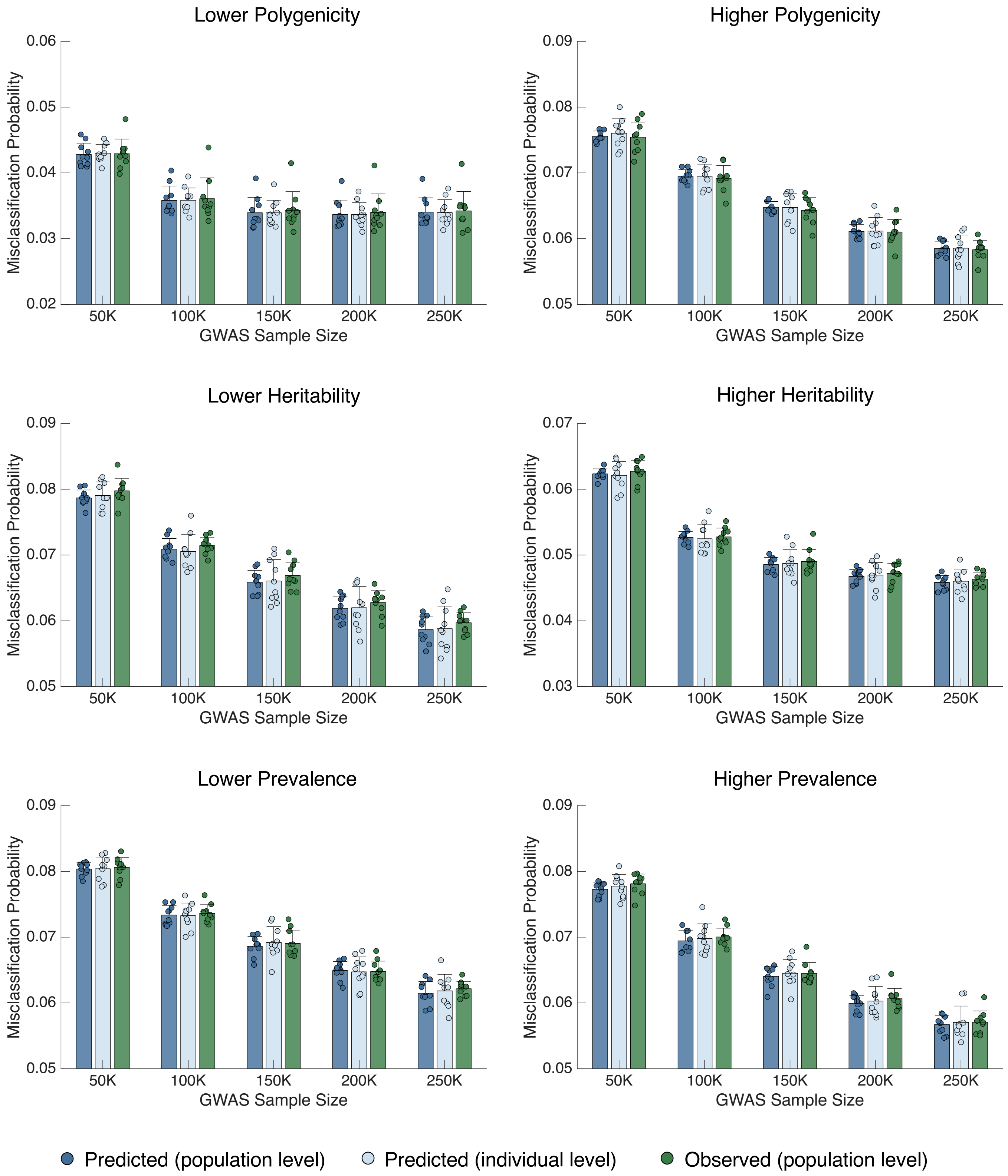


**Supplementary Figure 9:** Comparison of (i) the predicted population-level misclassification probability, (ii) the average predicted individual-level conditional misclassification probability, and (iii) the empirically observed overall misclassification rate in the testing dataset across simulation settings and GWAS training sample sizes, using the top 5% as the risk threshold. Bars and error bars indicate the mean and standard deviation across 10 simulation replicates, with individual estimates overlaid.

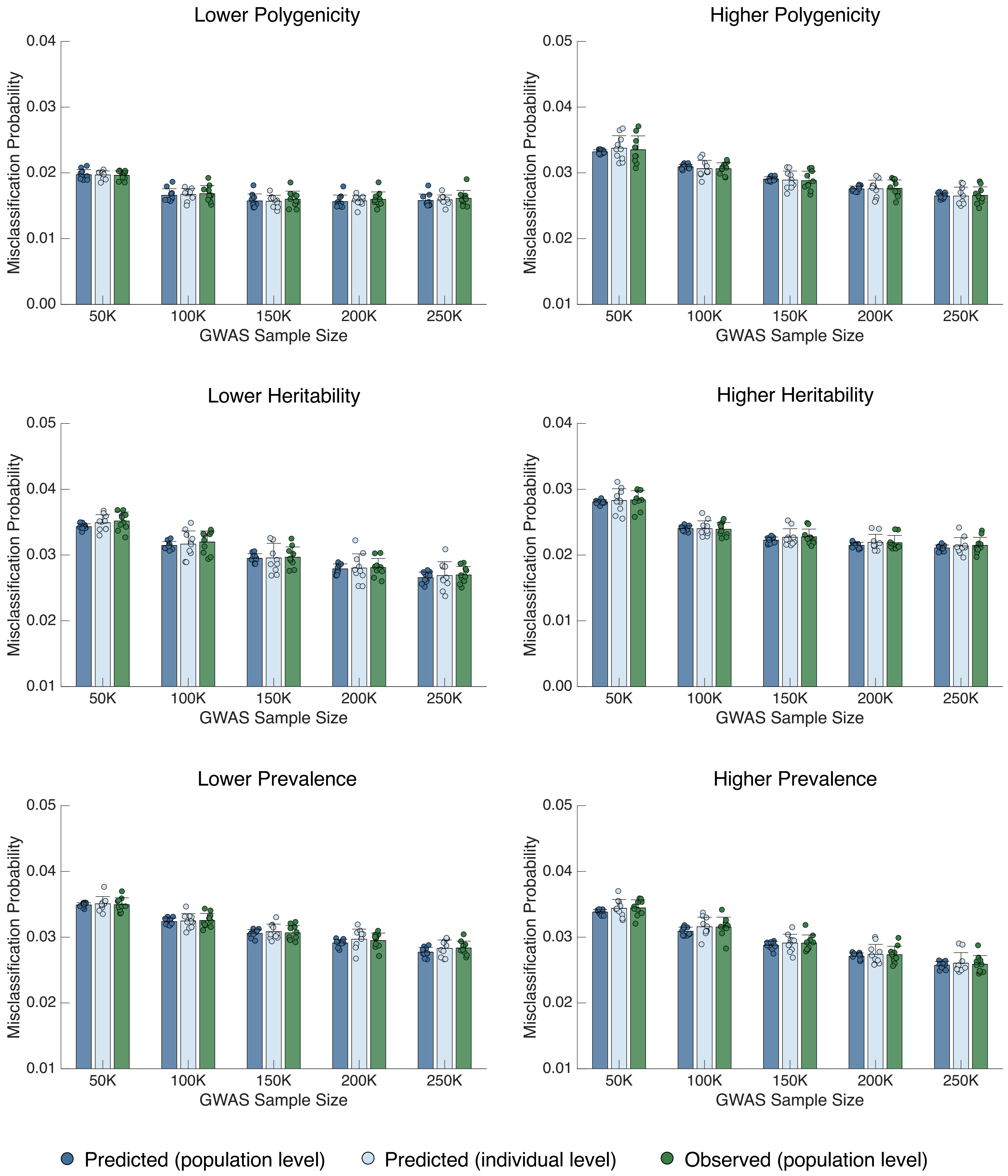


**Supplementary Figure 10:** Comparison of (i) the predicted population-level misclassification probability, (ii) the average predicted individual-level conditional misclassification probability, and (iii) the empirically observed overall misclassification rate in the testing dataset across simulation settings and GWAS training sample sizes, using the top 2% as the risk threshold. Bars and error bars indicate the mean and standard deviation across 10 simulation replicates, with individual estimates overlaid.

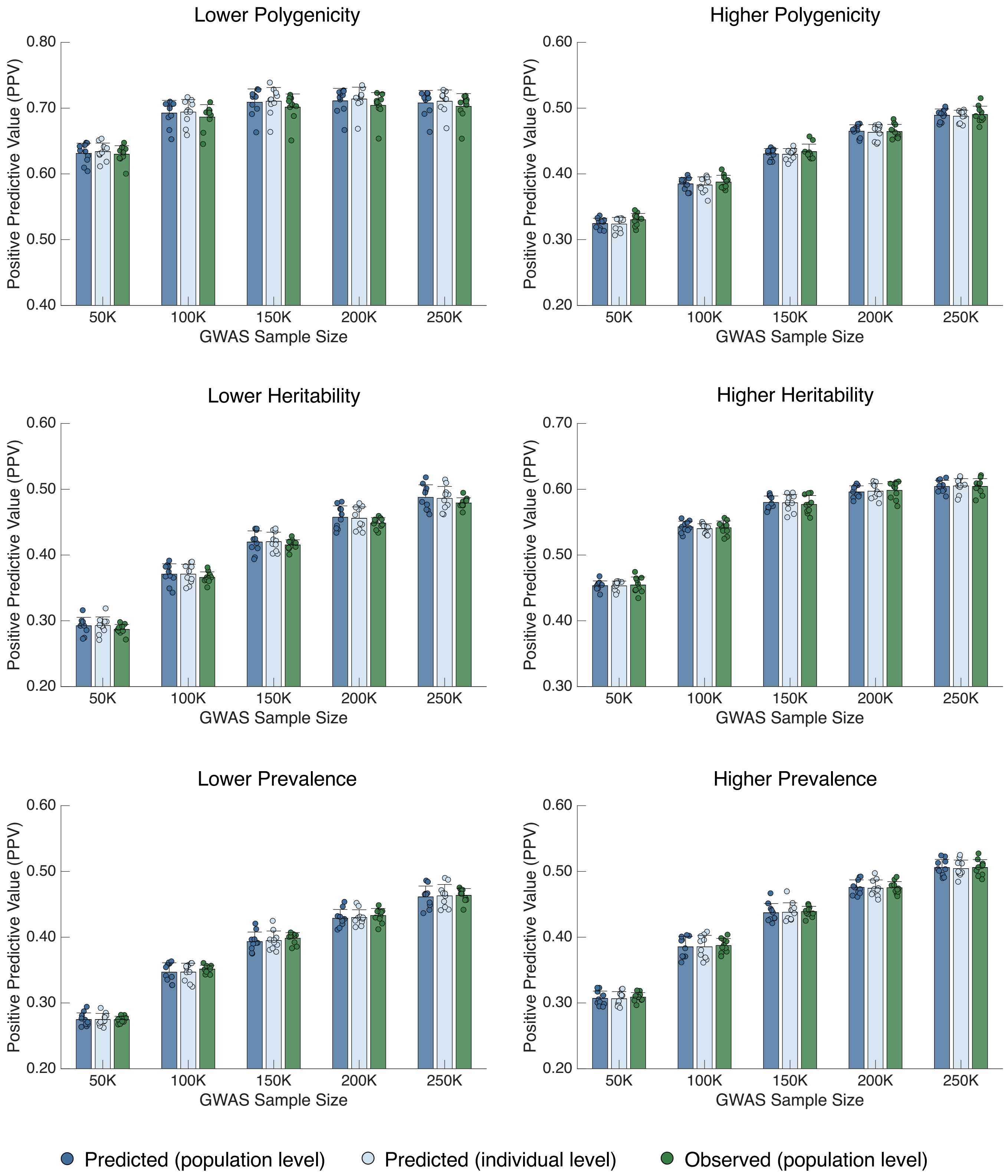


**Supplementary Figure 11:** Comparison of predicted and empirically observed positive predictive values (PPV) in the testing dataset across simulation settings and GWAS training sample sizes, using the top 10% as the risk threshold. Bars and error bars indicate the mean and standard deviation across 10 simulation replicates, with individual estimates overlaid.

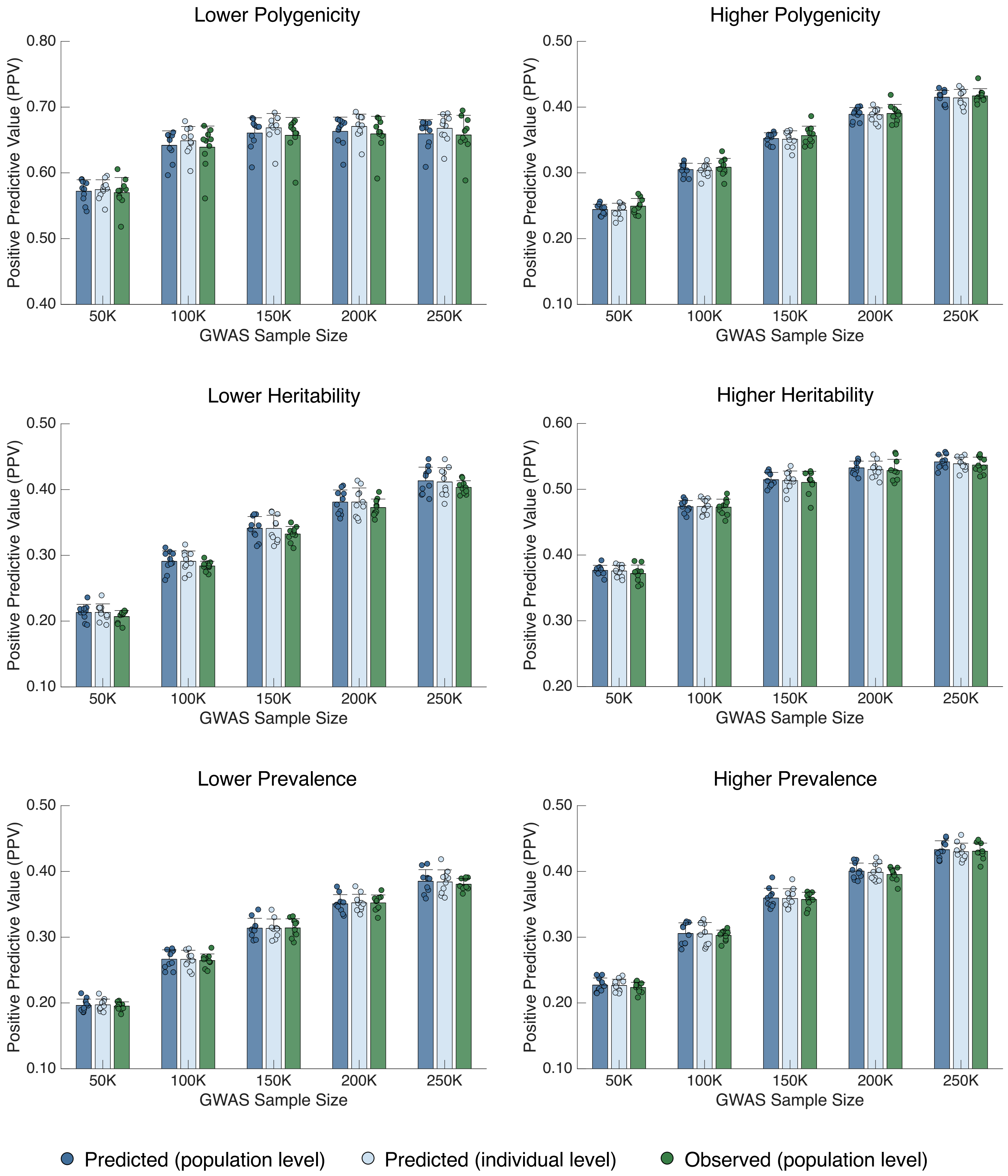


**Supplementary Figure 12:** Comparison of predicted and empirically observed positive predictive values (PPV) in the testing dataset across simulation settings and GWAS training sample sizes, using the top 5% as the risk threshold. Bars and error bars indicate the mean and standard deviation across 10 simulation replicates, with individual estimates overlaid.

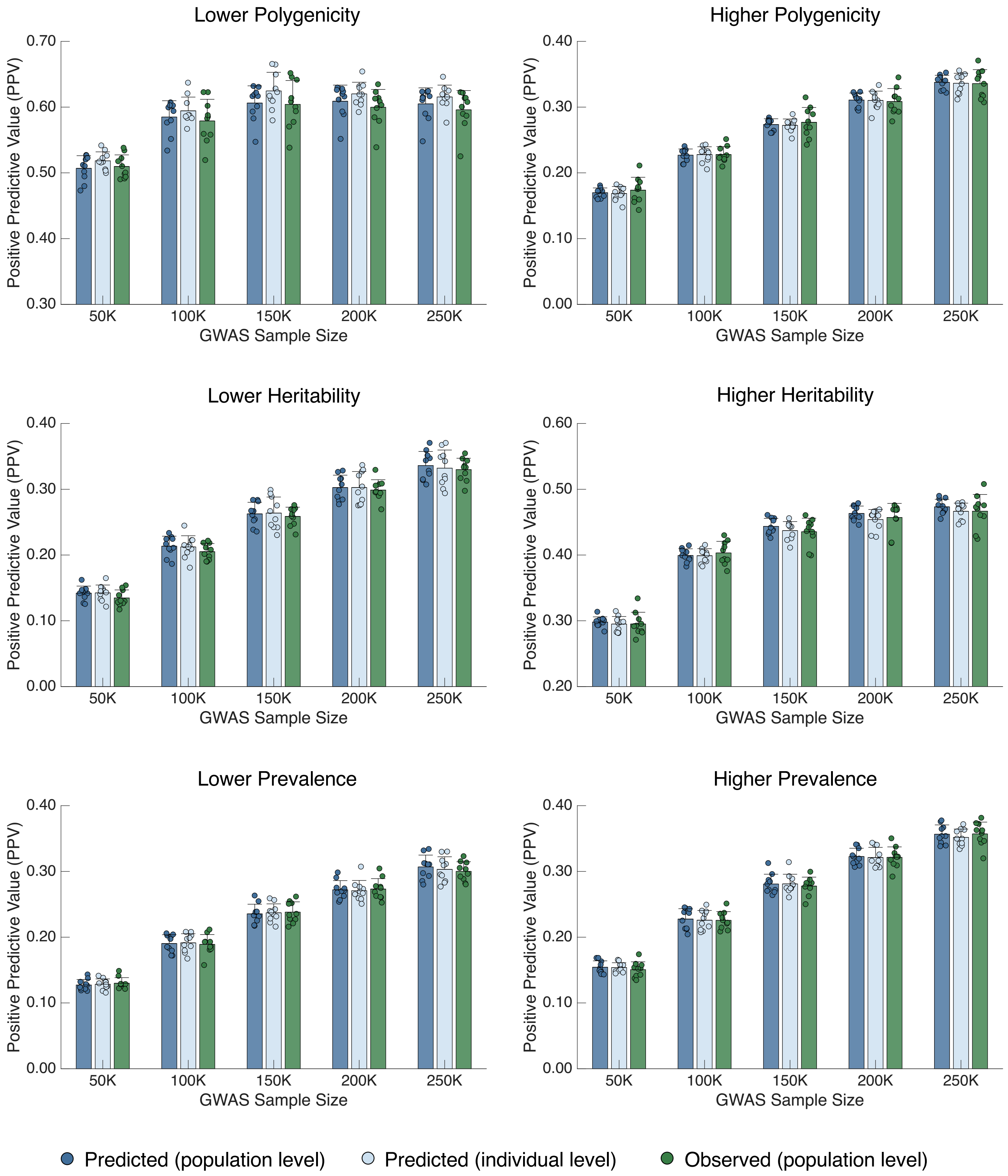


**Supplementary Figure 13:** Comparison of predicted and empirically observed positive predictive values (PPV) in the testing dataset across simulation settings and GWAS training sample sizes, using the top 2% as the risk threshold. Bars and error bars indicate the mean and standard deviation across 10 simulation replicates, with individual estimates overlaid.

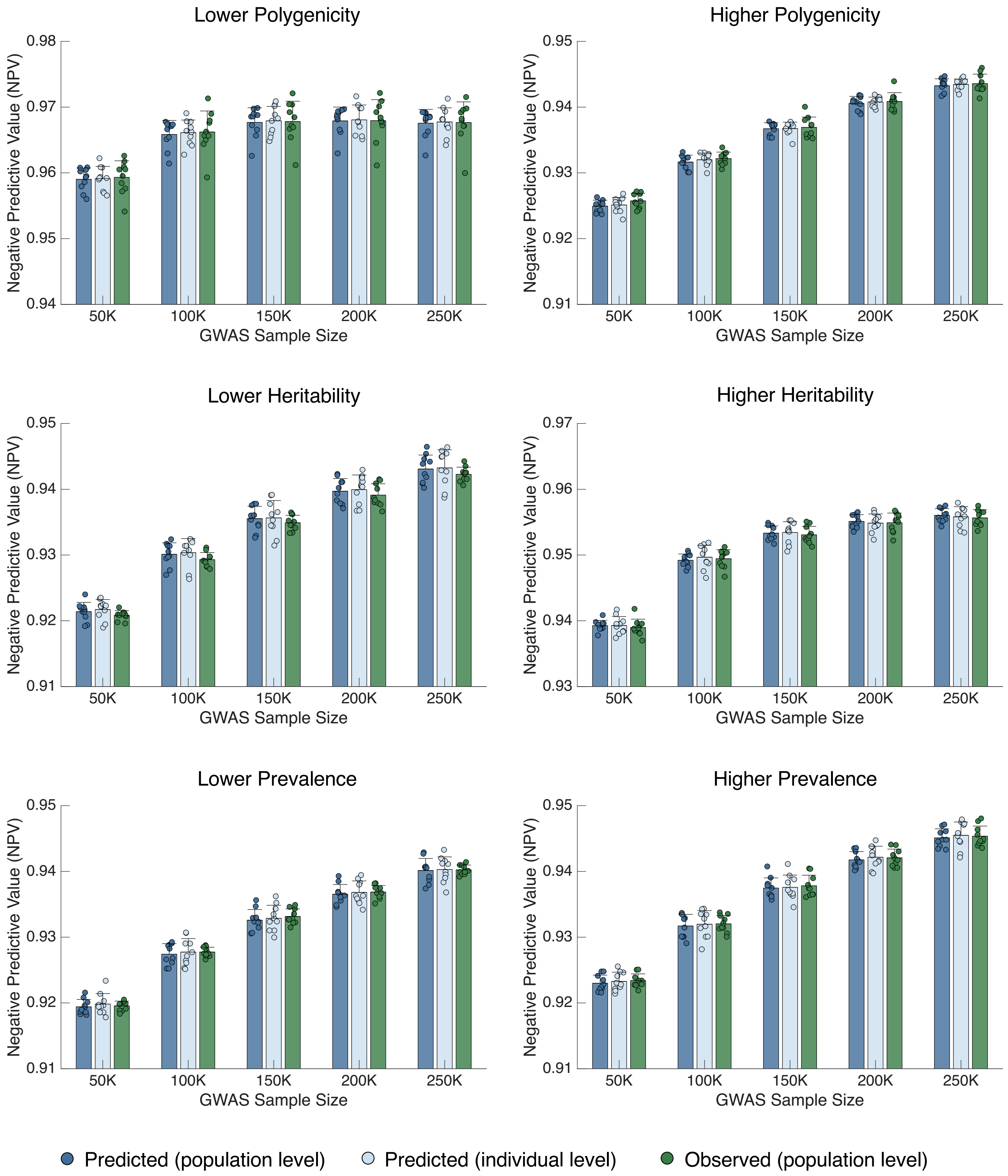


**Supplementary Figure 14:** Comparison of predicted and empirically observed negative predictive values (NPV) in the testing dataset across simulation settings and GWAS training sample sizes, using the top 10% as the risk threshold. Bars and error bars indicate the mean and standard deviation across 10 simulation replicates, with individual estimates overlaid.

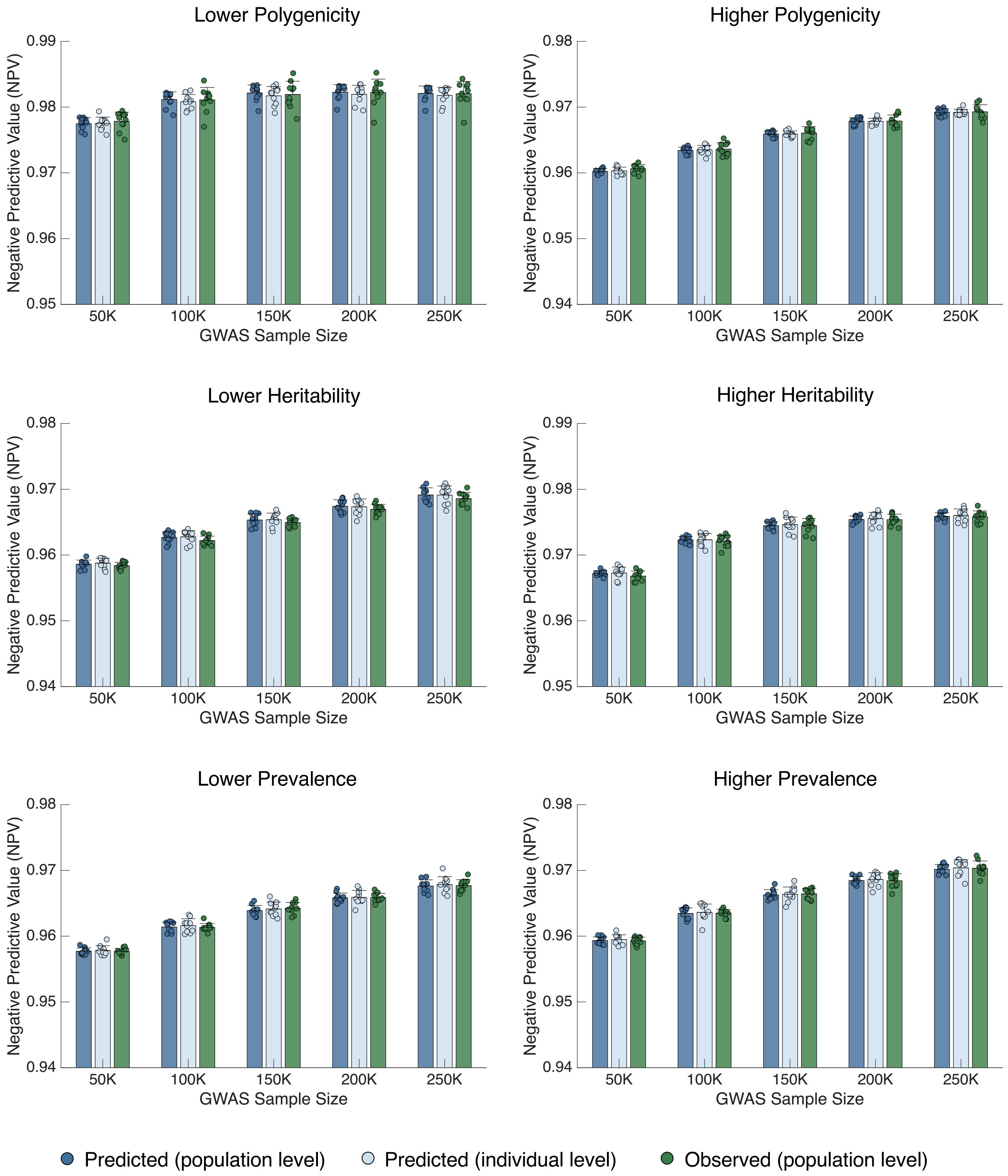


**Supplementary Figure 15:** Comparison of predicted and empirically observed negative predictive values (NPV) in the testing dataset across simulation settings and GWAS training sample sizes, using the top 5% as the risk threshold. Bars and error bars indicate the mean and standard deviation across 10 simulation replicates, with individual estimates overlaid.

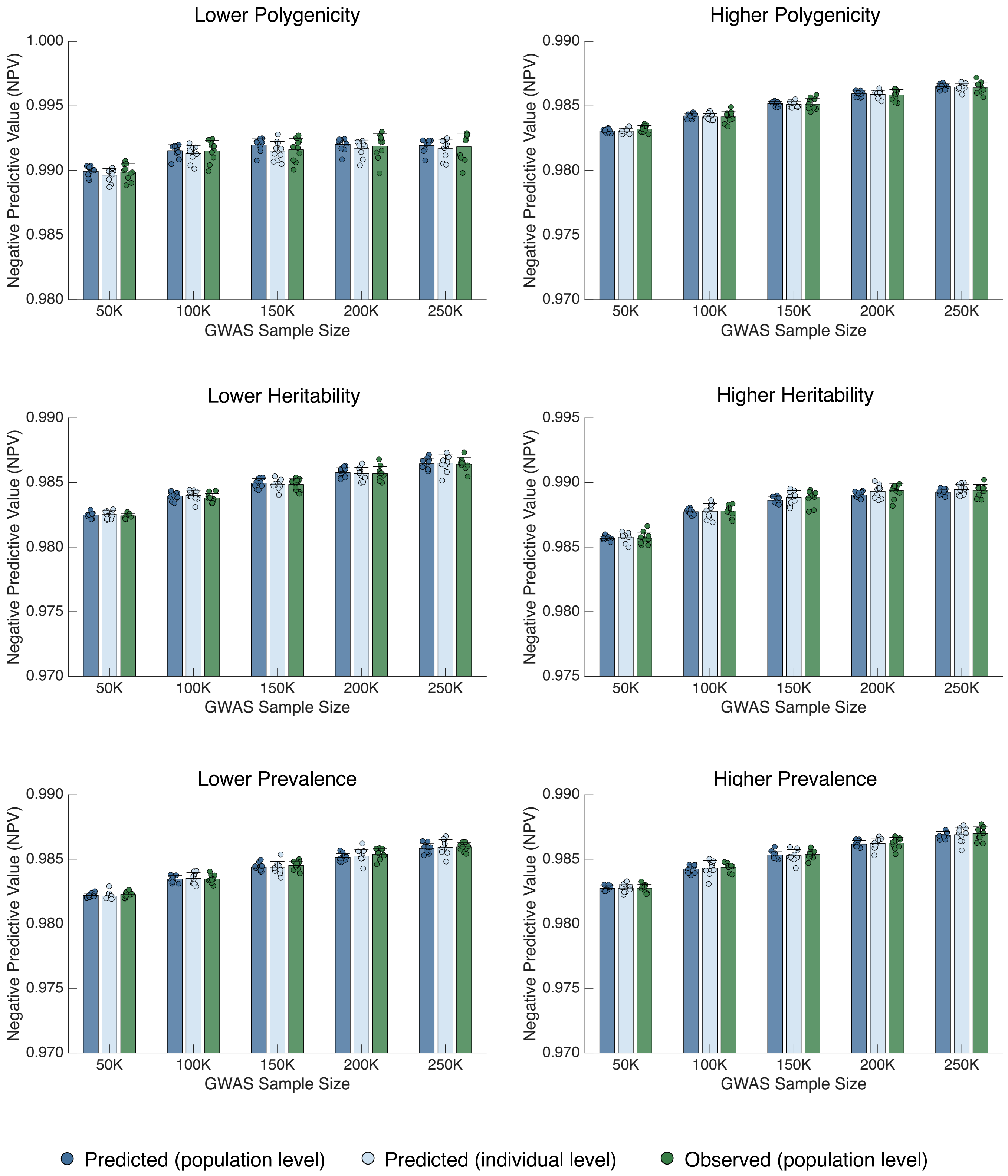


**Supplementary Figure 16:** Comparison of predicted and empirically observed negative predictive values (NPV) in the testing dataset across simulation settings and GWAS training sample sizes, using the top 2% as the risk threshold. Bars and error bars indicate the mean and standard deviation across 10 simulation replicates, with individual estimates overlaid.

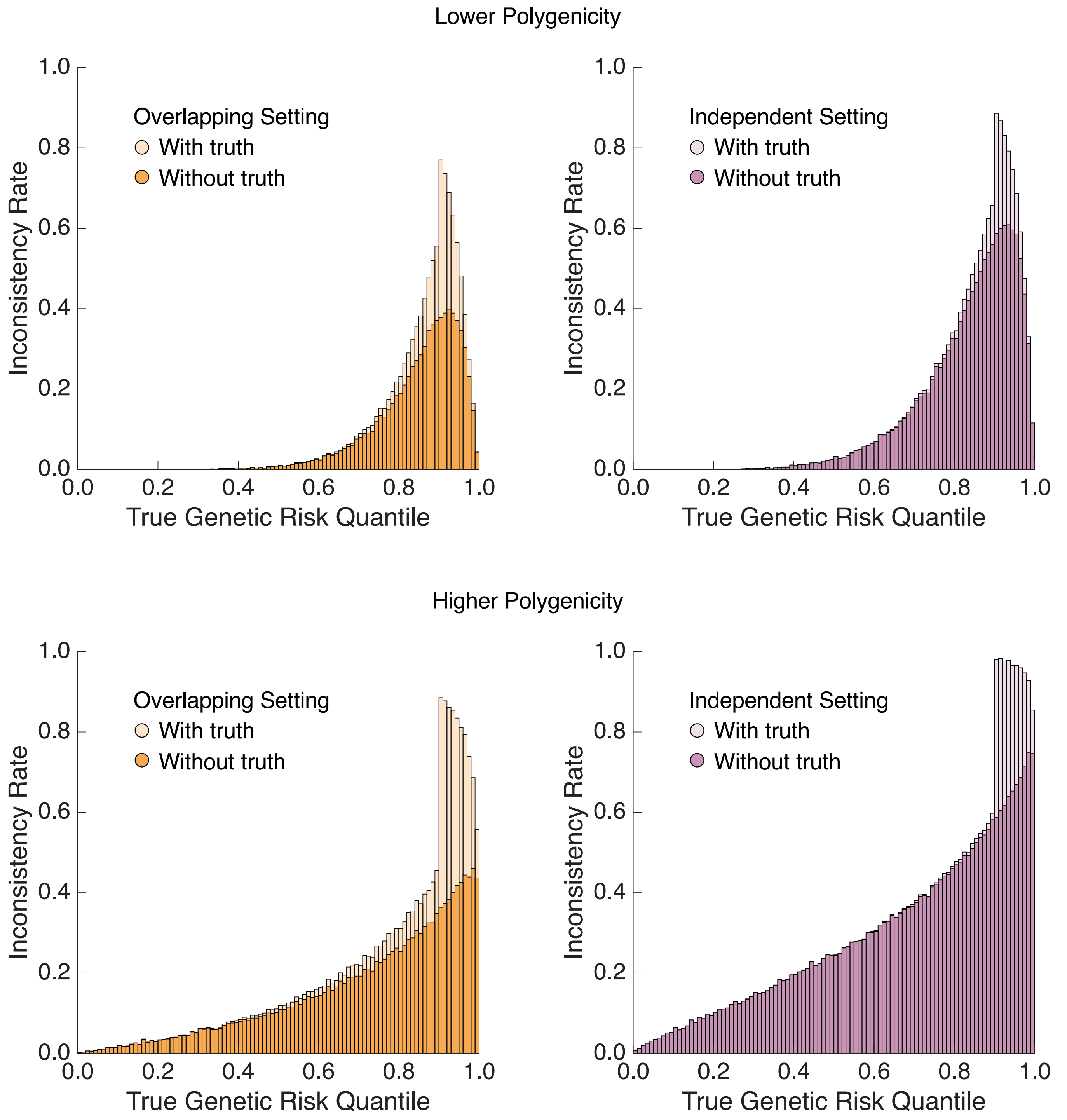


**Supplementary Figure 17:** The distribution of empirically observed with-truth (lighter color) and PRS-only (darker color) inconsistency rates across genetic risk percentiles in the testing dataset, under the overlapping (left panel) and independent (right panel) simulation settings and different polygenicity of the genetic architecture.

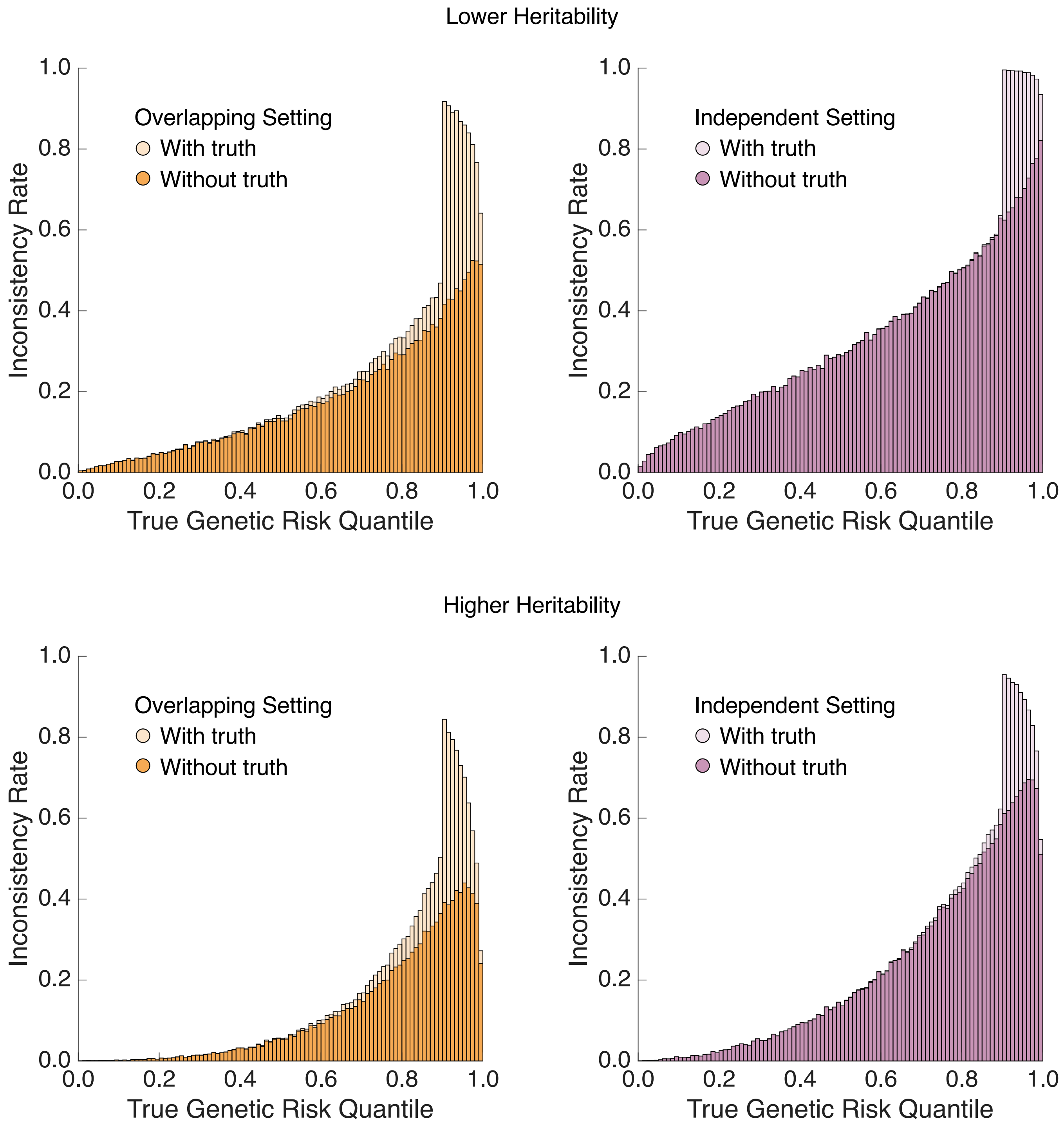


**Supplementary Figure 18:** The distribution of empirically observed with-truth (lighter color) and PRS-only (darker color) inconsistency rates across genetic risk percentiles in the testing dataset, under the overlapping (left panel) and independent (right panel) simulation settings and different SNP heritability.

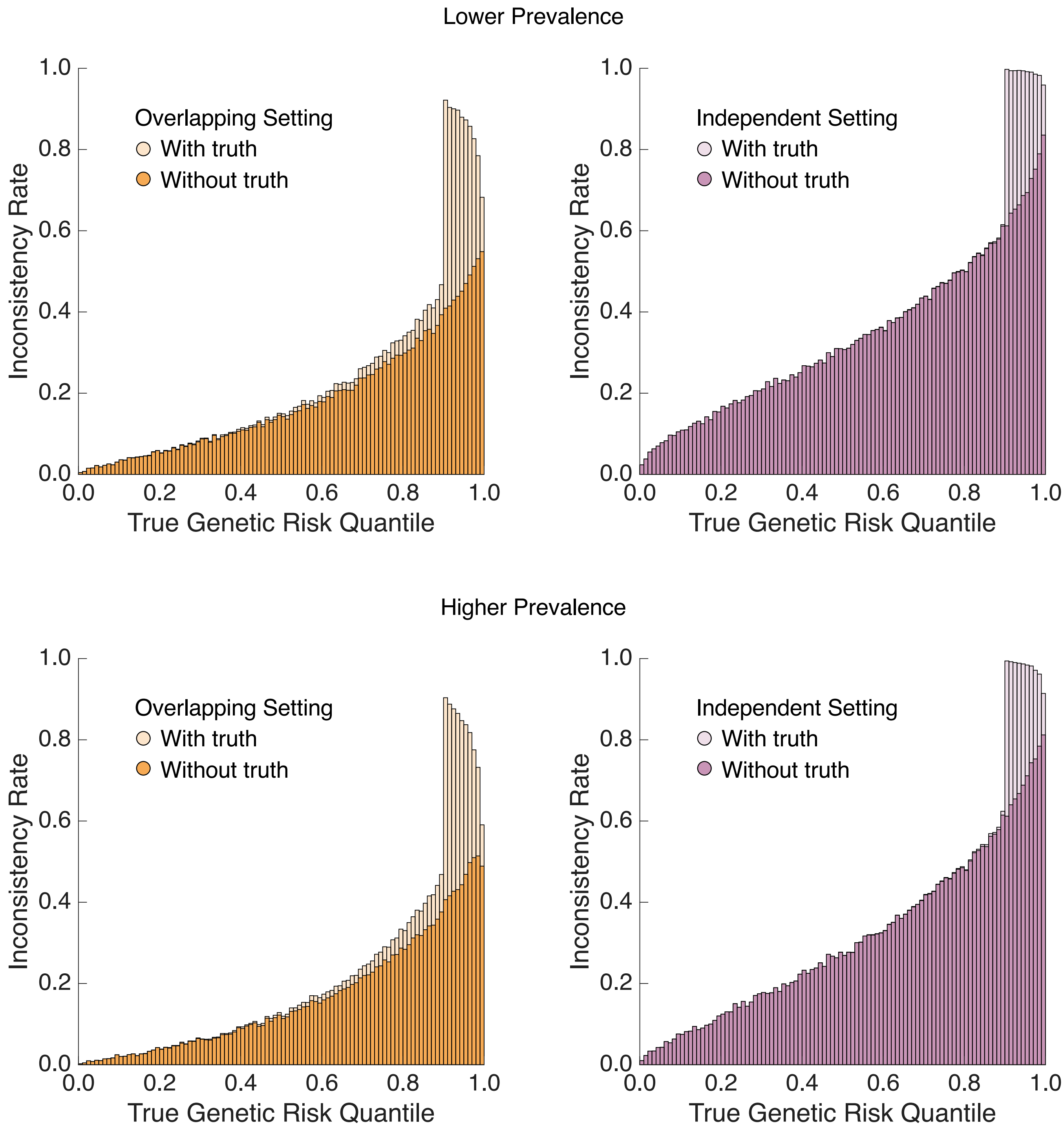


**Supplementary Figure 19:** The distribution of empirically observed with-truth (lighter color) and PRS-only (darker color) inconsistency rates across genetic risk percentiles in the testing dataset, under the overlapping (left panel) and independent (right panel) simulation settings and different prevalences of the binary phenotype.

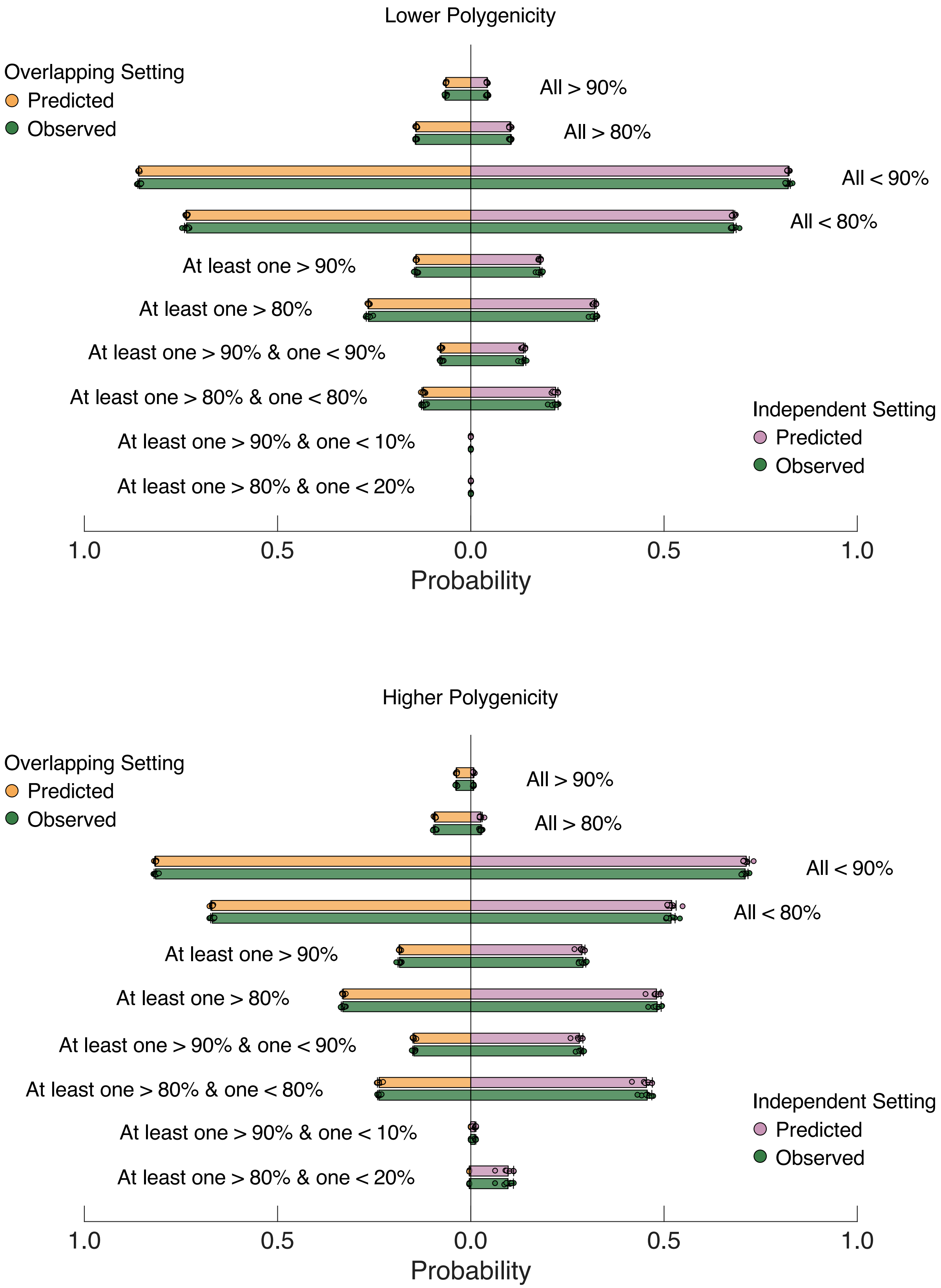


**Supplementary Figure 20:** Comparison of predicted and empirically observed probabilities of PRS classification profiles in the testing dataset under the overlapping (left panel) and independent (right panel) simulation settings and different polygenicity of the genetic architecture (upper and lower panels). Bars and error bars indicate the mean and standard deviation across 10 simulation replicates, with individual estimates overlaid.

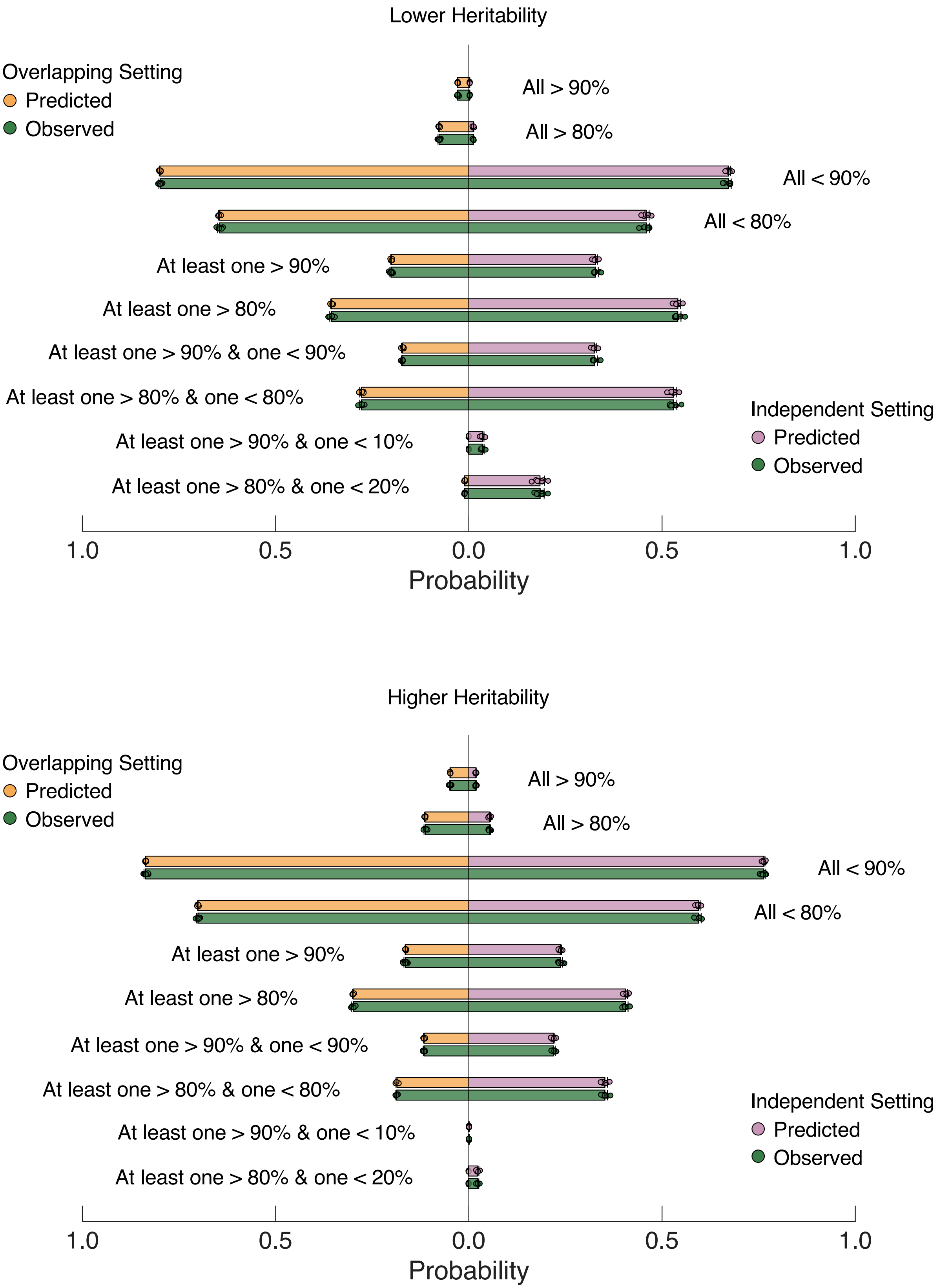


**Supplementary Figure 21:** Comparison of predicted and empirically observed probabilities of PRS classification profiles in the testing dataset under the overlapping (left panel) and independent (right panel) simulation settings and different SNP heritability (upper and lower panels). Bars and error bars indicate the mean and standard deviation across 10 simulation replicates, with individual estimates overlaid.

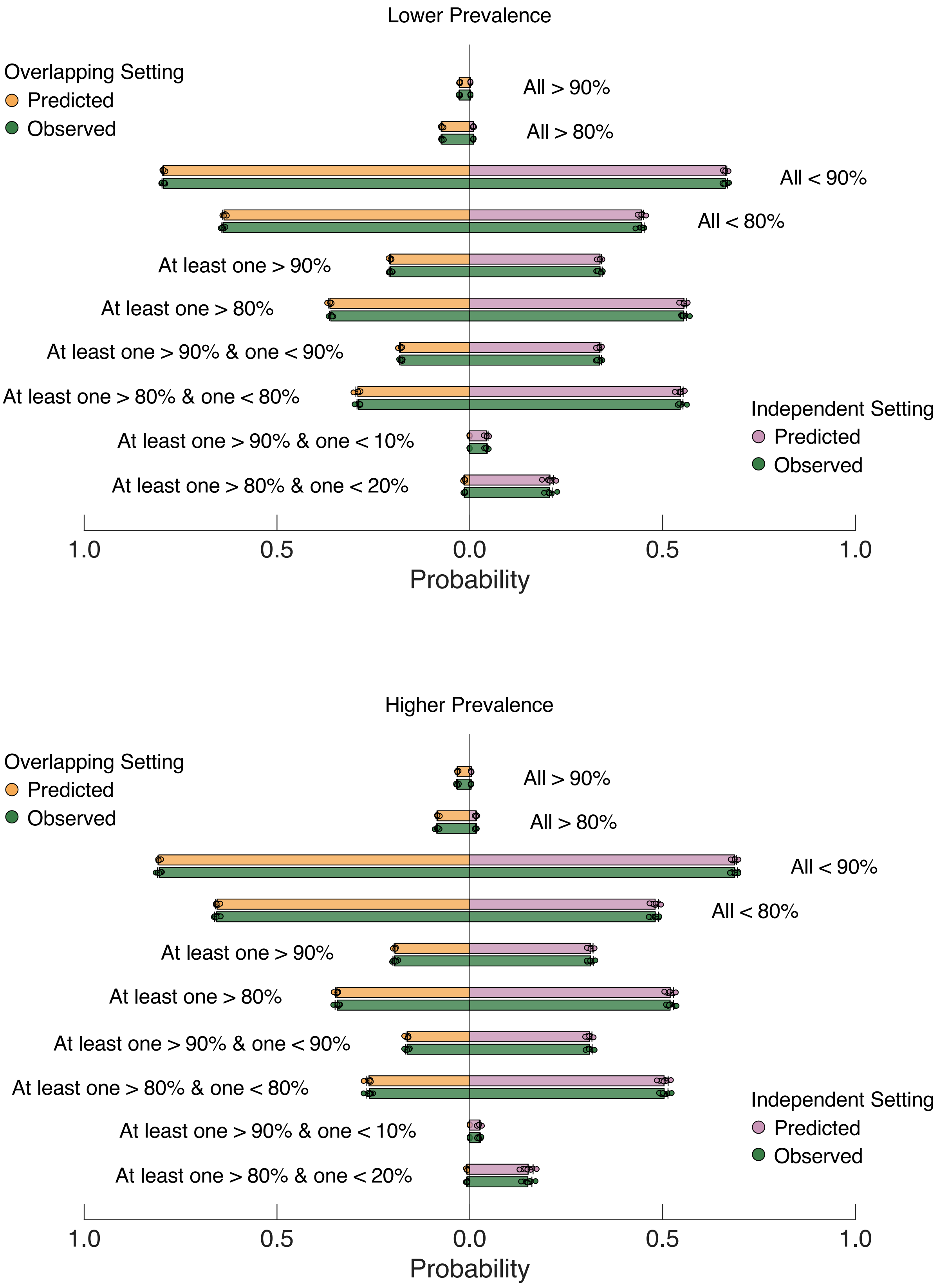


**Supplementary Figure 22:** Comparison of predicted and empirically observed probabilities of PRS classification profiles in the testing dataset under the overlapping (left panel) and independent (right panel) simulation settings and different prevalences of the binary phenotype (upper and lower panels). Bars and error bars indicate the mean and standard deviation across 10 simulation replicates, with individual estimates overlaid.

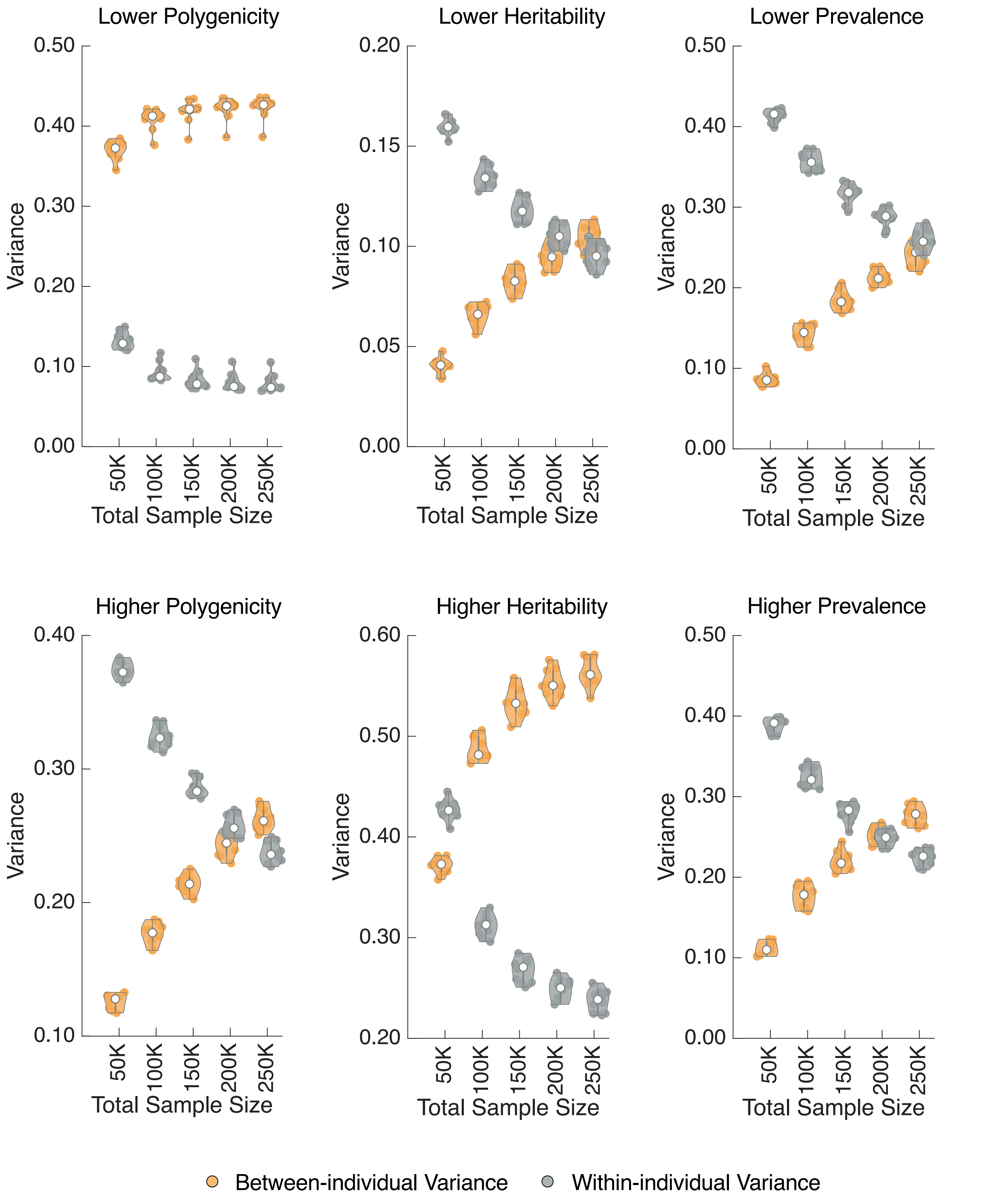


**Supplementary Figure 23:** Between-individual variance of the PRS point estimate and average within-individual posterior variance for sequentially integrated PRSs in the testing dataset under the overlapping design across simulation settings. Each violin plot shows the median across 10 simulation replicates, with individual estimates overlaid.

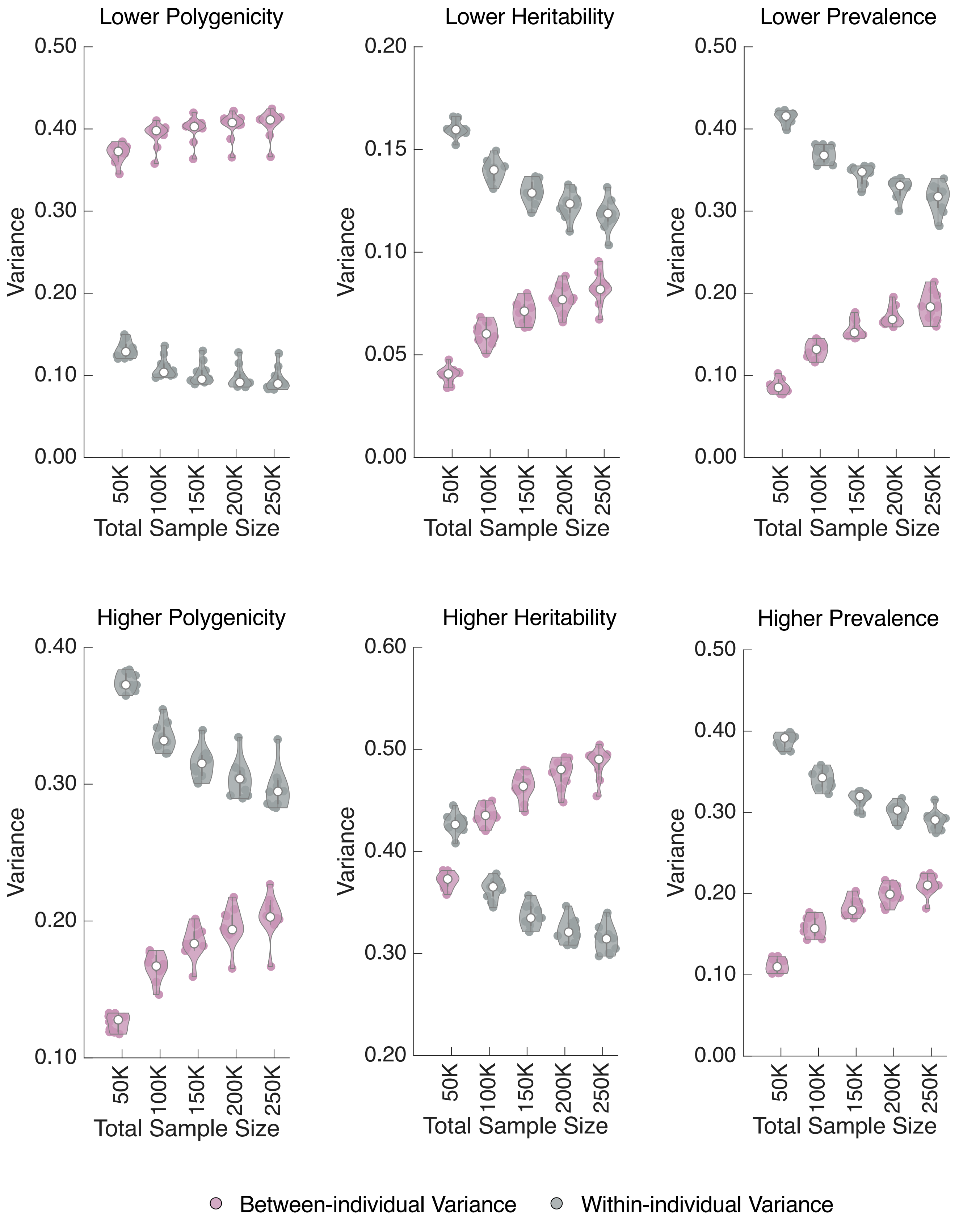


**Supplementary Figure 24:** Between-individual variance of the PRS point estimate and average within-individual posterior variance for sequentially integrated PRSs in the testing dataset under the independent design across simulation settings. Each violin plot shows the median across 10 simulation replicates, with individual estimates overlaid.

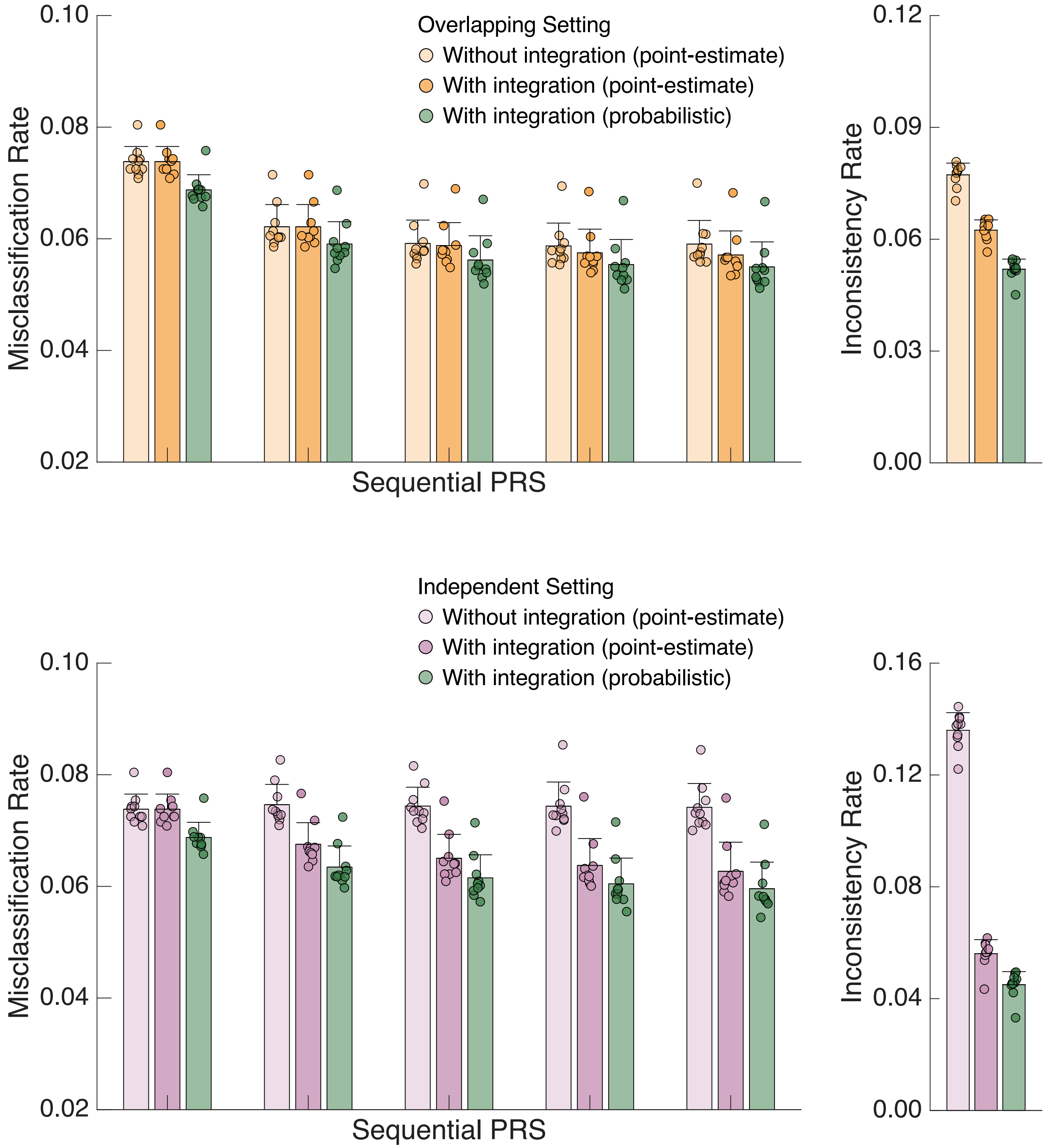


**Supplementary Figure 25:** *Left*: Comparison of (i) the empirically observed misclassification rate without PRS integration using point-estimate thresholding, (ii) the empirically observed misclassification rate for sequentially integrated PRSs using point-estimate thresholding, and (iii) the empirically observed misclassification rate for sequentially integrated PRSs using uncertainty-aware probabilistic thresholding in the testing dataset under the overlapping (upper panel) and independent (lower panel) designs and the lower polygenicity simulation setting. *Right*: Comparison of empirically observed classification inconsistency rates with and without PRS integration, using either point-estimate thresholding or uncertainty-aware probabilistic thresholding, in the testing dataset under the overlapping (upper panel) and independent (lower panel) designs and the lower polygenicity simulation setting. Bars and error bars indicate the mean and standard deviation across 10 simulation replicates, with individual estimates overlaid.

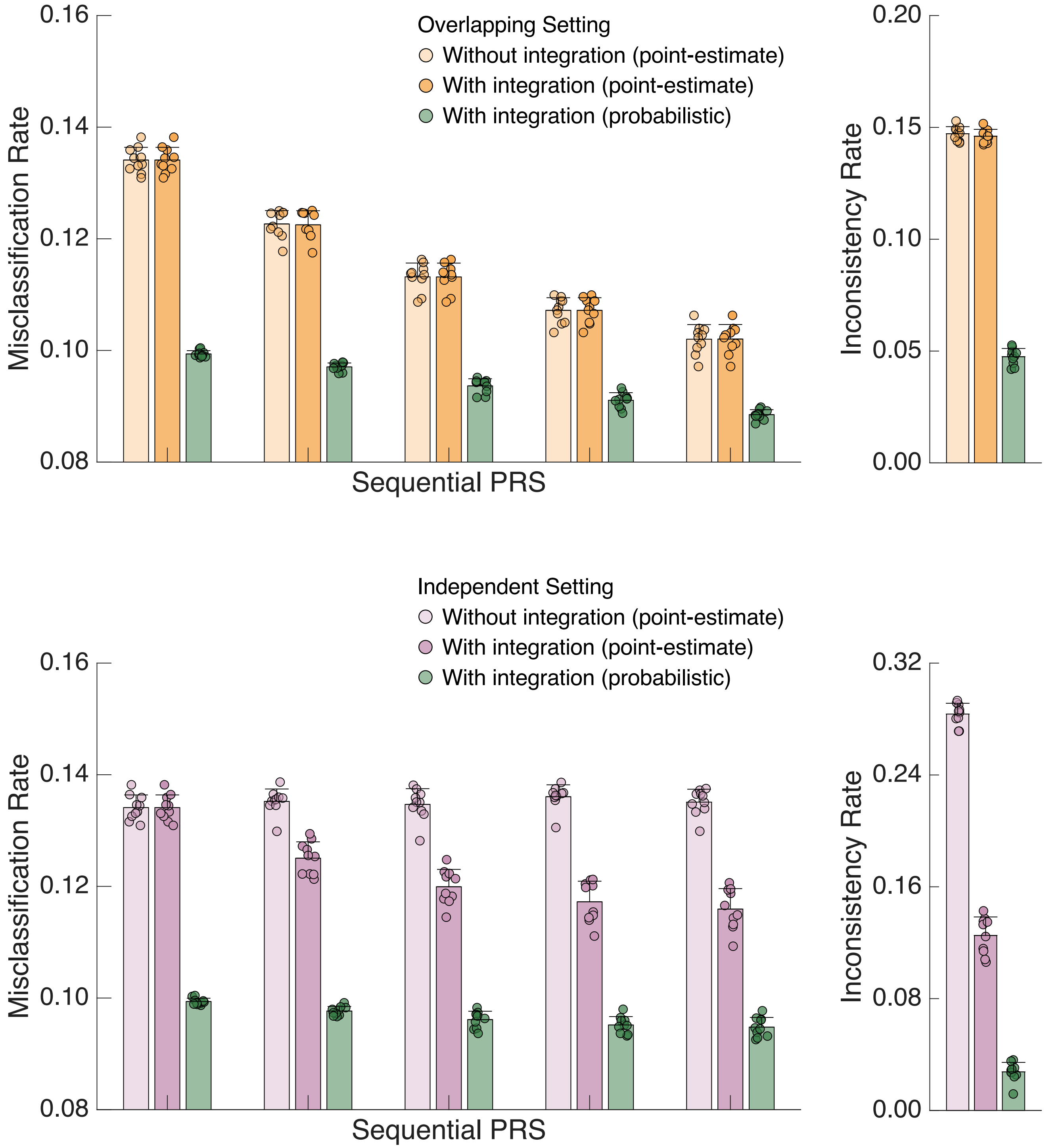


**Supplementary Figure 26:** *Left*: Comparison of (i) the empirically observed misclassification rate without PRS integration using point-estimate thresholding, (ii) the empirically observed misclassification rate for sequentially integrated PRSs using point-estimate thresholding, and (iii) the empirically observed misclassification rate for sequentially integrated PRSs using uncertainty-aware probabilistic thresholding in the testing dataset under the overlapping (upper panel) and independent (lower panel) designs and the higher polygenicity simulation setting. *Right*: Comparison of empirically observed classification inconsistency rates with and without PRS integration, using either point-estimate thresholding or uncertainty-aware probabilistic thresholding, in the testing dataset under the overlapping (upper panel) and independent (lower panel) designs and the higher polygenicity simulation setting. Bars and error bars indicate the mean and standard deviation across 10 simulation replicates, with individual estimates overlaid.

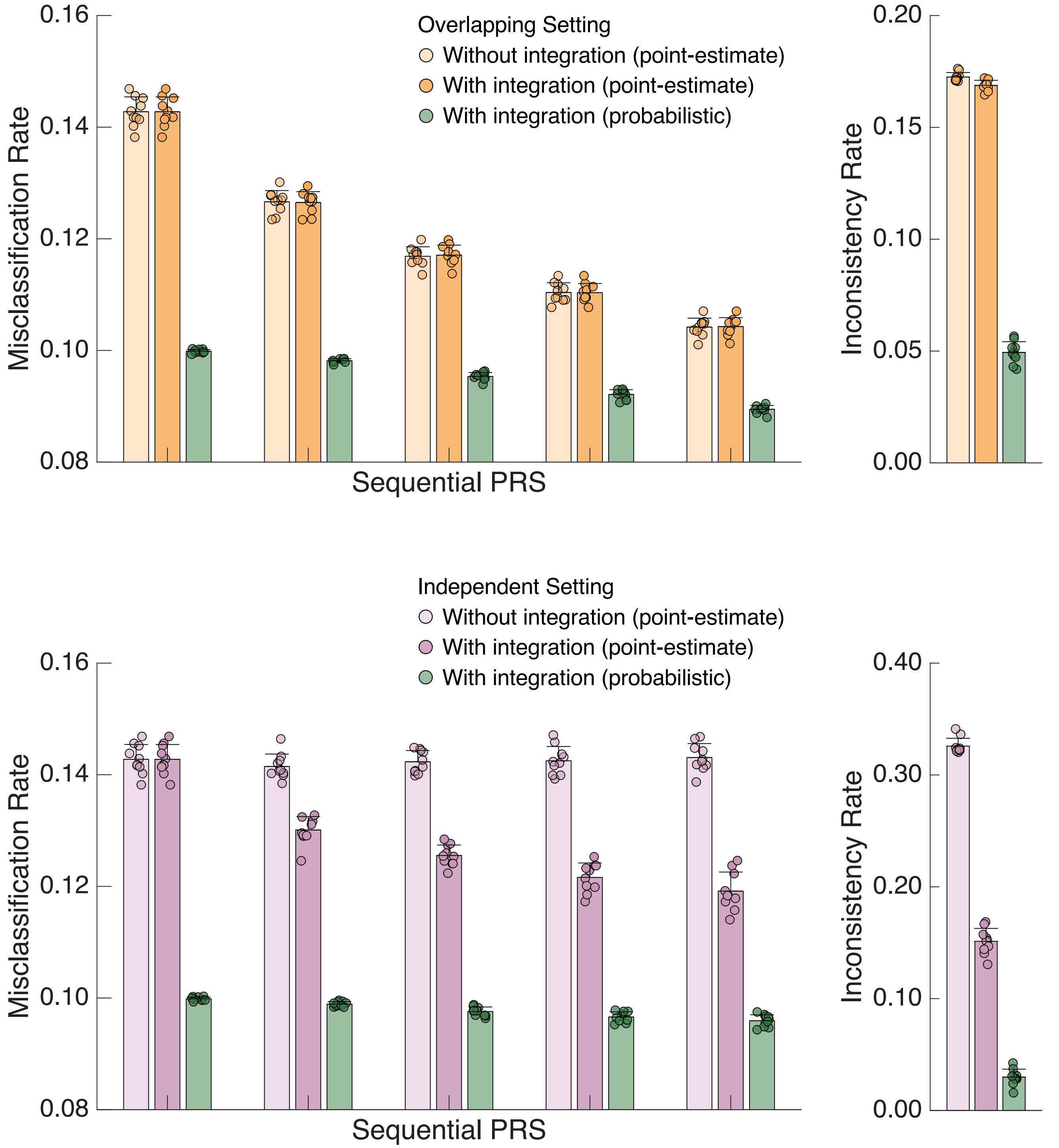


**Supplementary Figure 27:** *Left*: Comparison of (i) the empirically observed misclassification rate without PRS integration using point-estimate thresholding, (ii) the empirically observed misclassification rate for sequentially integrated PRSs using point-estimate thresholding, and (iii) the empirically observed misclassification rate for sequentially integrated PRSs using uncertainty-aware probabilistic thresholding in the testing dataset under the overlapping (upper panel) and independent (lower panel) designs and the lower heritability simulation setting. *Right*: Comparison of empirically observed classification inconsistency rates with and without PRS integration, using either point-estimate thresholding or uncertainty-aware probabilistic thresholding, in the testing dataset under the overlapping (upper panel) and independent (lower panel) designs and the lower heritability simulation setting. Bars and error bars indicate the mean and standard deviation across 10 simulation replicates, with individual estimates overlaid.

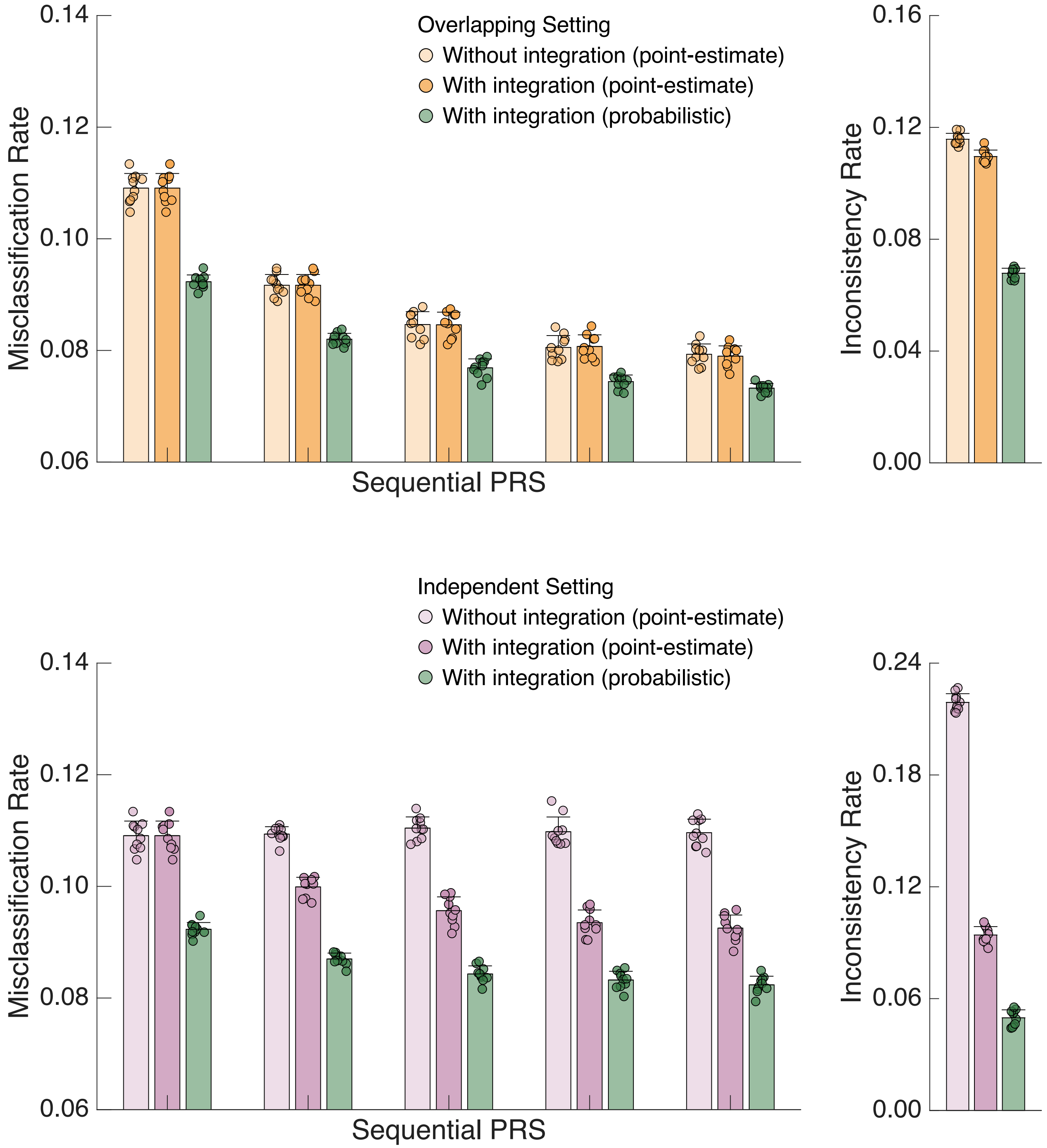


**Supplementary Figure 28:** *Left*: Comparison of (i) the empirically observed misclassification rate without PRS integration using point-estimate thresholding, (ii) the empirically observed misclassification rate for sequentially integrated PRSs using point-estimate thresholding, and (iii) the empirically observed misclassification rate for sequentially integrated PRSs using uncertainty-aware probabilistic thresholding in the testing dataset under the overlapping (upper panel) and independent (lower panel) designs and the higher heritability simulation setting. *Right*: Comparison of empirically observed classification inconsistency rates with and without PRS integration, using either point-estimate thresholding or uncertainty-aware probabilistic thresholding, in the testing dataset under the overlapping (upper panel) and independent (lower panel) designs and the higher heritability simulation setting. Bars and error bars indicate the mean and standard deviation across 10 simulation replicates, with individual estimates overlaid.

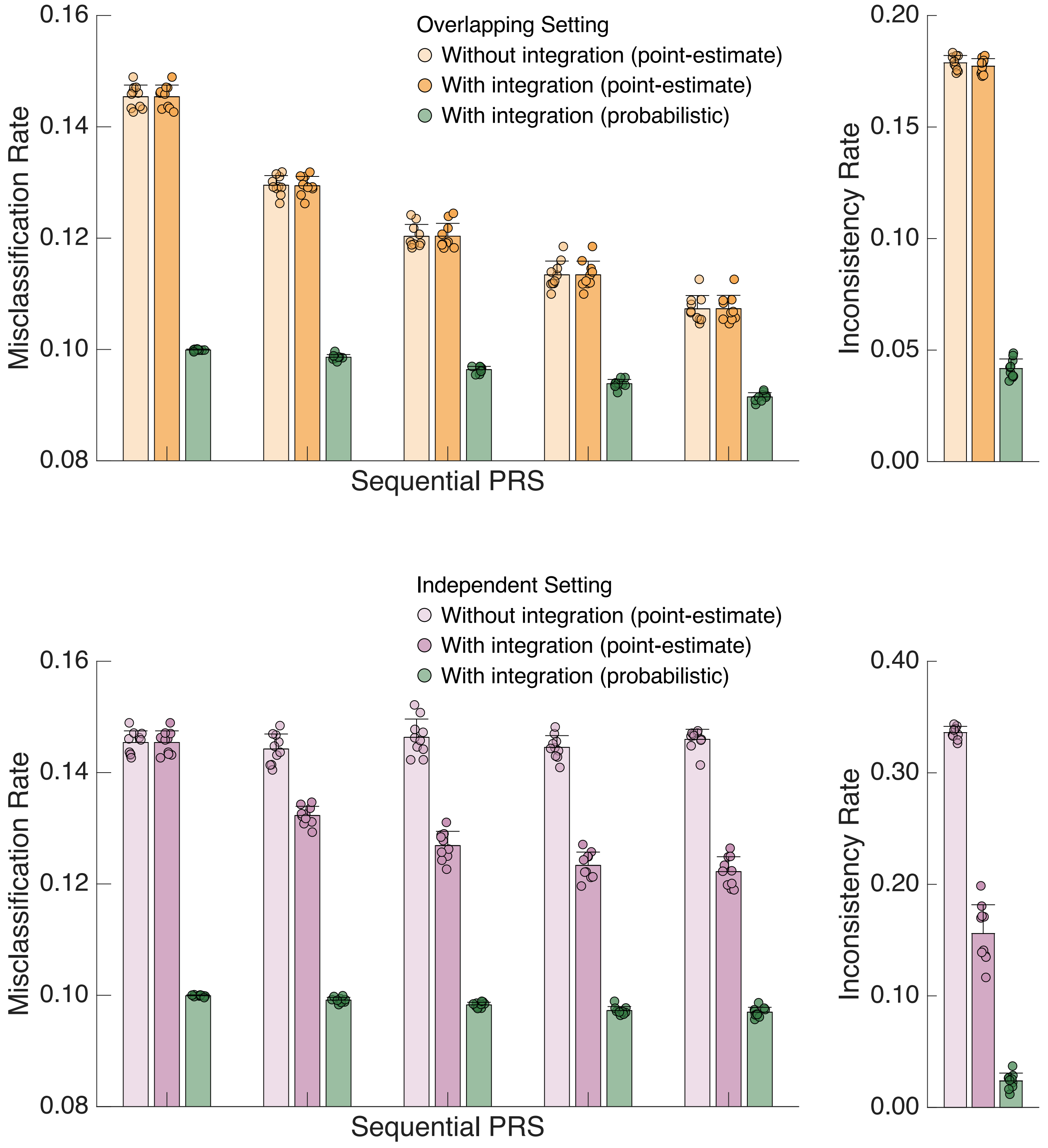


**Supplementary Figure 29:** *Left*: Comparison of (i) the empirically observed misclassification rate without PRS integration using point-estimate thresholding, (ii) the empirically observed misclassification rate for sequentially integrated PRSs using point-estimate thresholding, and (iii) the empirically observed misclassification rate for sequentially integrated PRSs using uncertainty-aware probabilistic thresholding in the testing dataset under the overlapping (upper panel) and independent (lower panel) designs and the lower prevalence simulation setting for binary phenotypes. *Right*: Comparison of empirically observed classification inconsistency rates with and without PRS integration, using either point-estimate thresholding or uncertainty-aware probabilistic thresholding, in the testing dataset under the overlapping (upper panel) and independent (lower panel) designs and the lower prevalence simulation setting for binary phenotypes. Bars and error bars indicate the mean and standard deviation across 10 simulation replicates, with individual estimates overlaid.

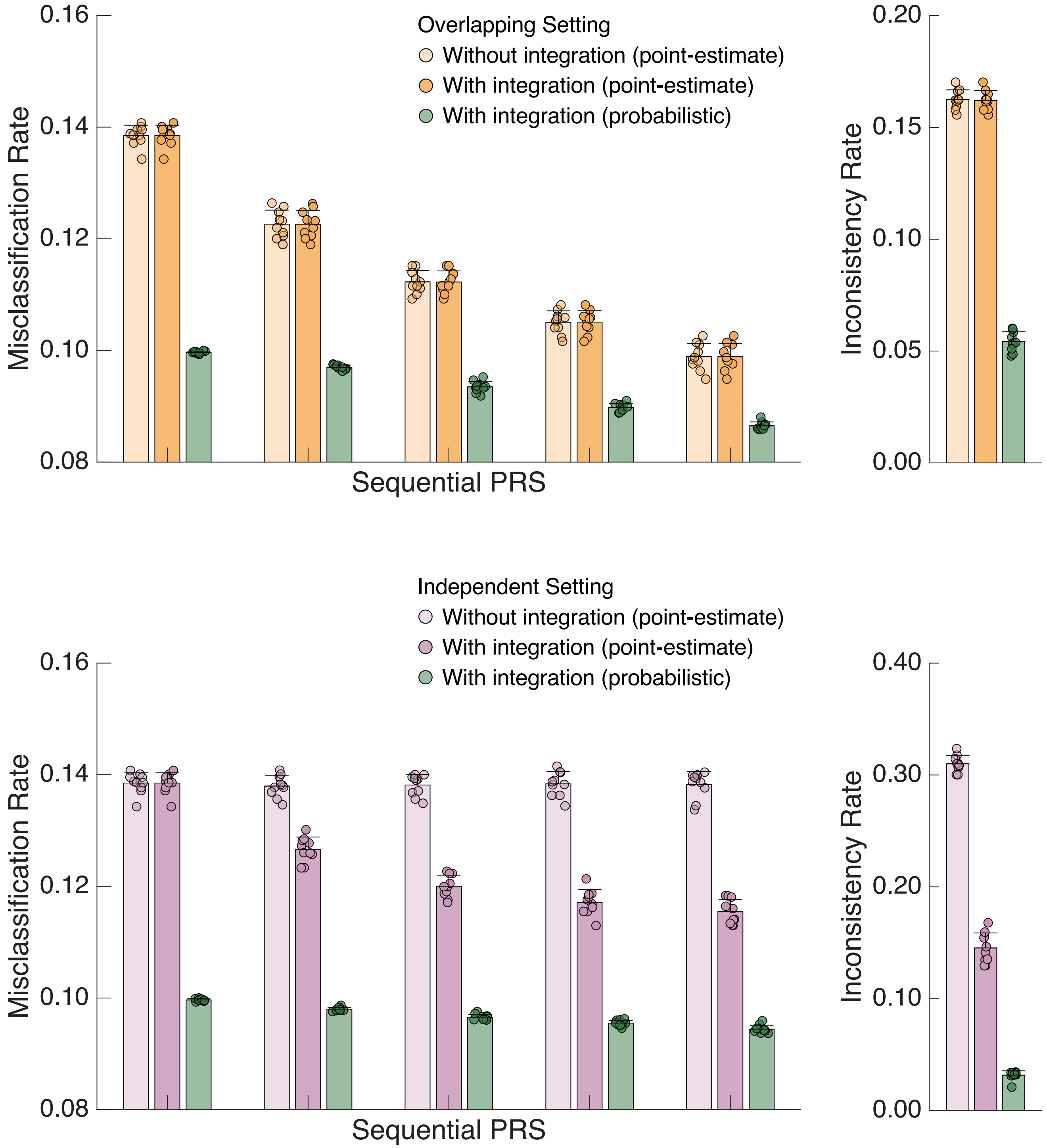


**Supplementary Figure 30:** *Left*: Comparison of (i) the empirically observed misclassification rate without PRS integration using point-estimate thresholding, (ii) the empirically observed misclassification rate for sequentially integrated PRSs using point-estimate thresholding, and (iii) the empirically observed misclassification rate for sequentially integrated PRSs using uncertainty-aware probabilistic thresholding in the testing dataset under the overlapping (upper panel) and independent (lower panel) designs and the higher prevalence simulation setting for binary phenotypes. *Right*: Comparison of empirically observed classification inconsistency rates with and without PRS integration, using either point-estimate thresholding or uncertainty-aware probabilistic thresholding, in the testing dataset under the overlapping (upper panel) and independent (lower panel) designs and the higher prevalence simulation setting for binary phenotypes. Bars and error bars indicate the mean and standard deviation across 10 simulation replicates, with individual estimates overlaid.

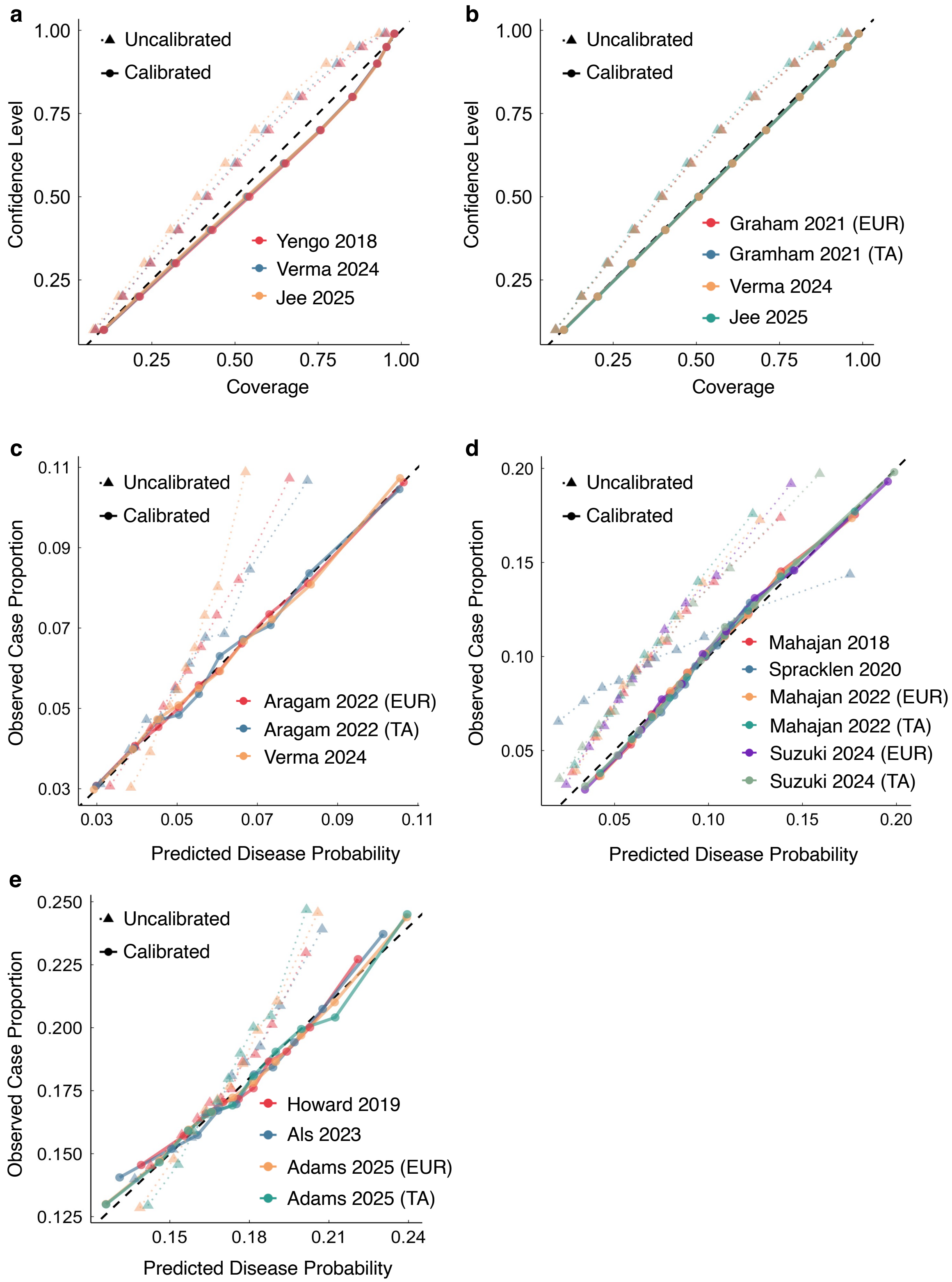


**Supplementary Figure 31: a-b,** Calibration of BMI and TC PRSs in the AoU testing set, comparing confidence levels against empirical coverage (the proportion of confidence intervals that contain the observed phenotype value). **c-e,** Calibration of CAD, T2D, and MDD PRSs in the AoU testing set, comparing observed case proportions against predicted probabilities.

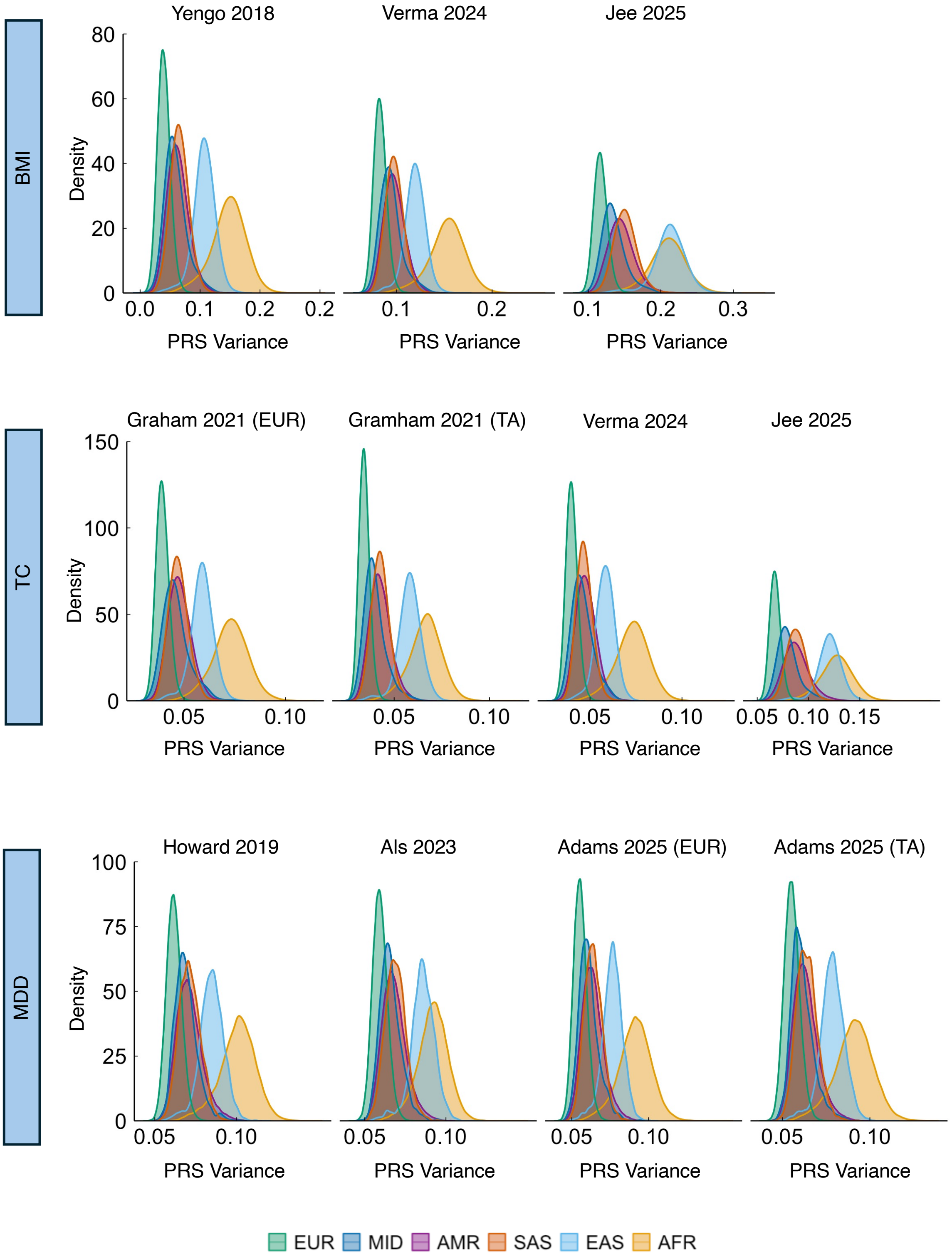


**Supplementary Figure 32:** Distribution of within-individual posterior variance (uncertainty), stratified by ancestry, for each BMI, TC, and MDD PRS after calibration.

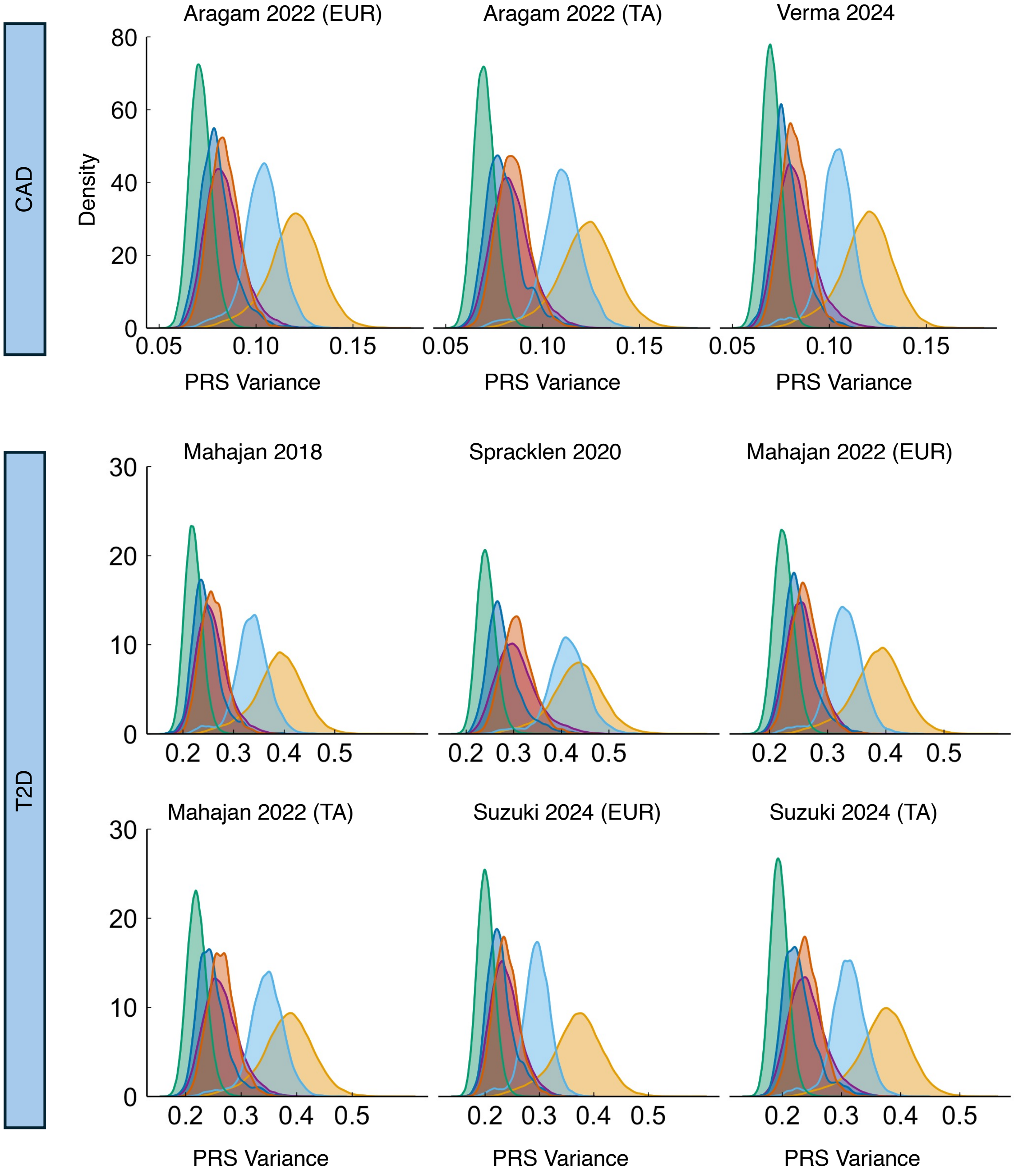


**Supplementary Figure 33:** Distribution of within-individual posterior variance (uncertainty), stratified by ancestry, for each CAD and T2D PRS after calibration.

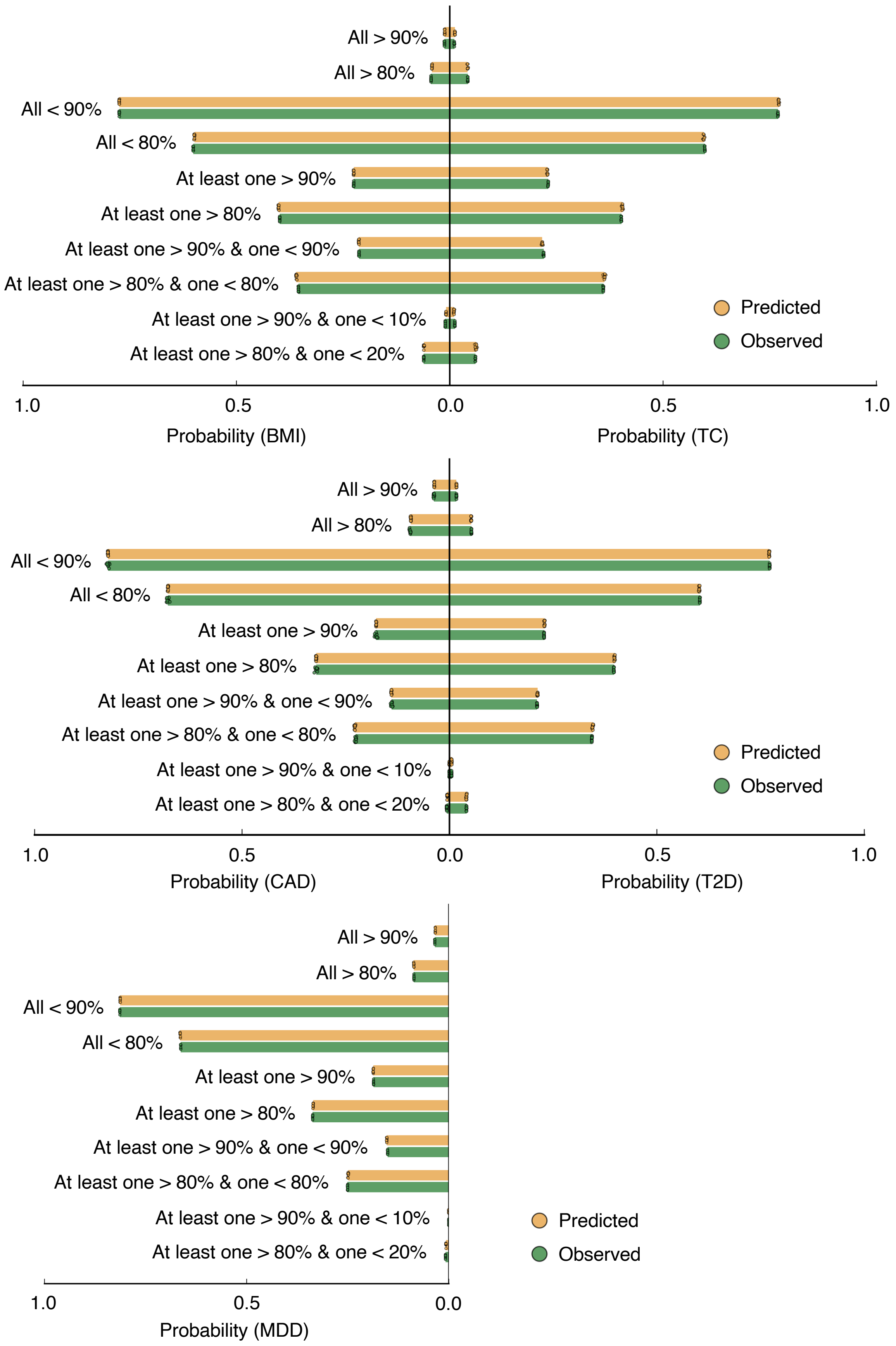


**Supplementary Figure 34:** Comparison of predicted and empirically observed probabilities of PRS classification profiles for five representative complex traits and diseases. Bars indicate the mean across 10 data splits, with individual estimates overlaid.

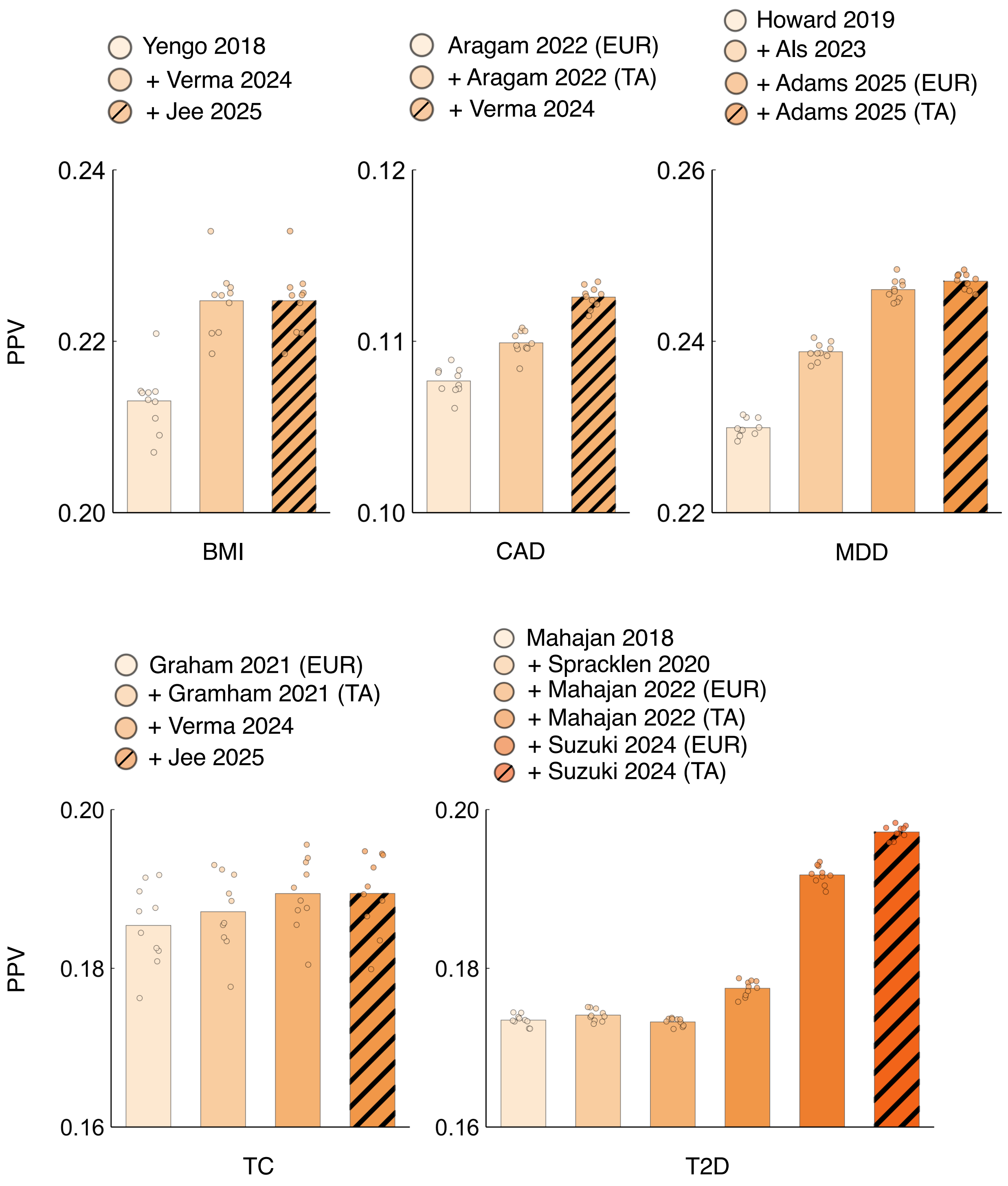


**Supplementary Figure 35:** Positive predictive value (PPV) of sequentially integrated PRSs for five representative complex traits and diseases, evaluated using a top 10% point-estimate threshold.

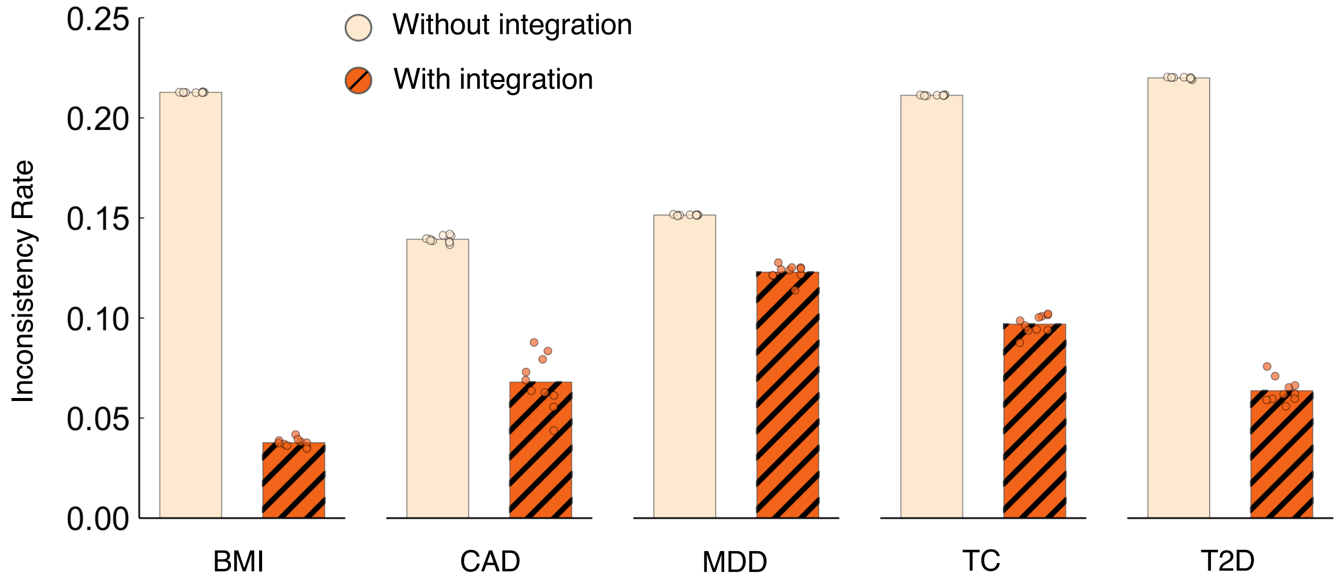


**Supplementary Figure 36:** Comparison of empirically observed classification inconsistency rates with and without PRS integration for five representative complex traits and diseases, using a top 10% point-estimate threshold.

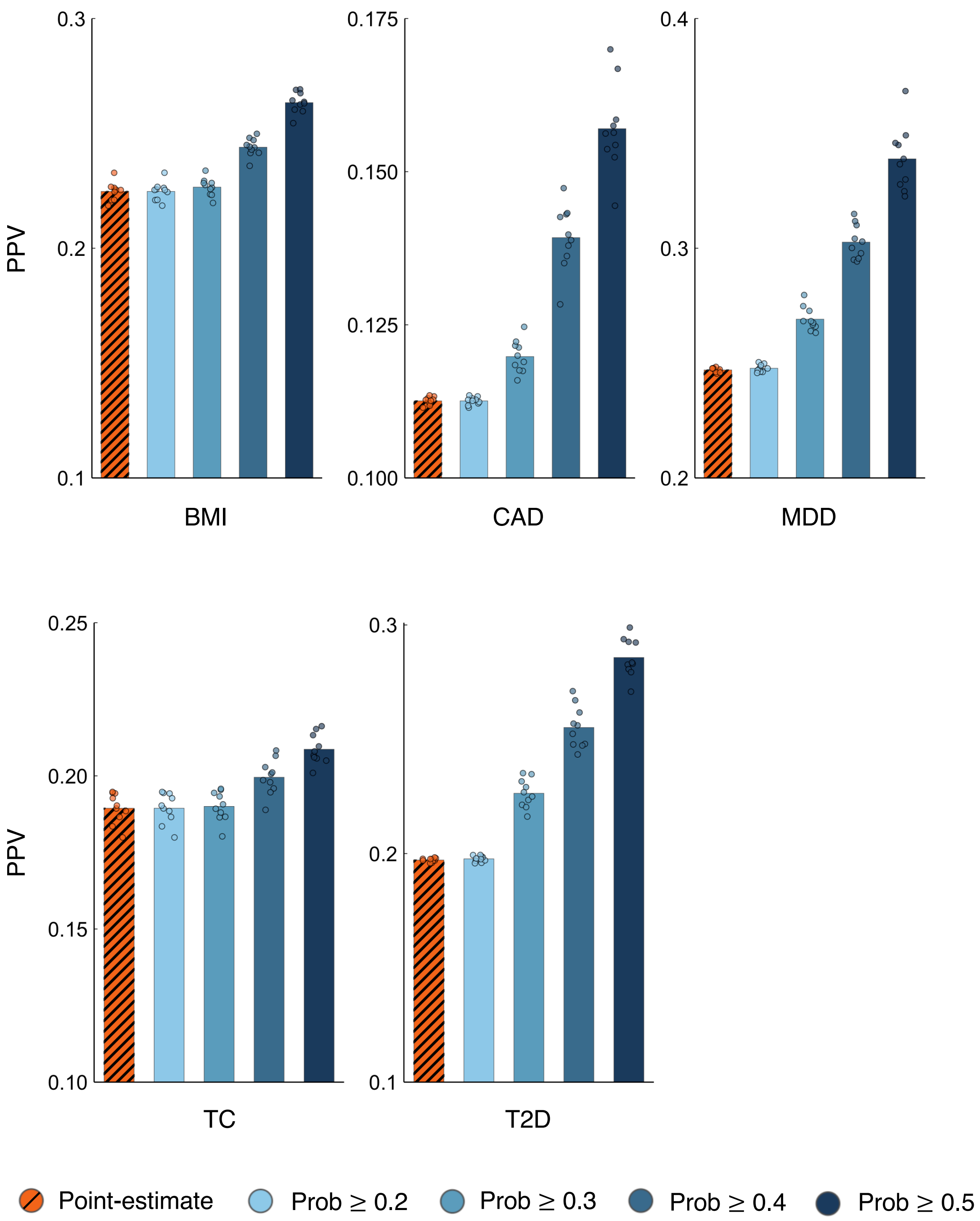


**Supplementary Figure 37:** Comparison of positive predictive value (PPV) for fully integrated PRSs of five representative complex traits and diseases under point-estimate thresholding versus uncertainty-aware probabilistic thresholding across a range of posterior probability cutoffs.

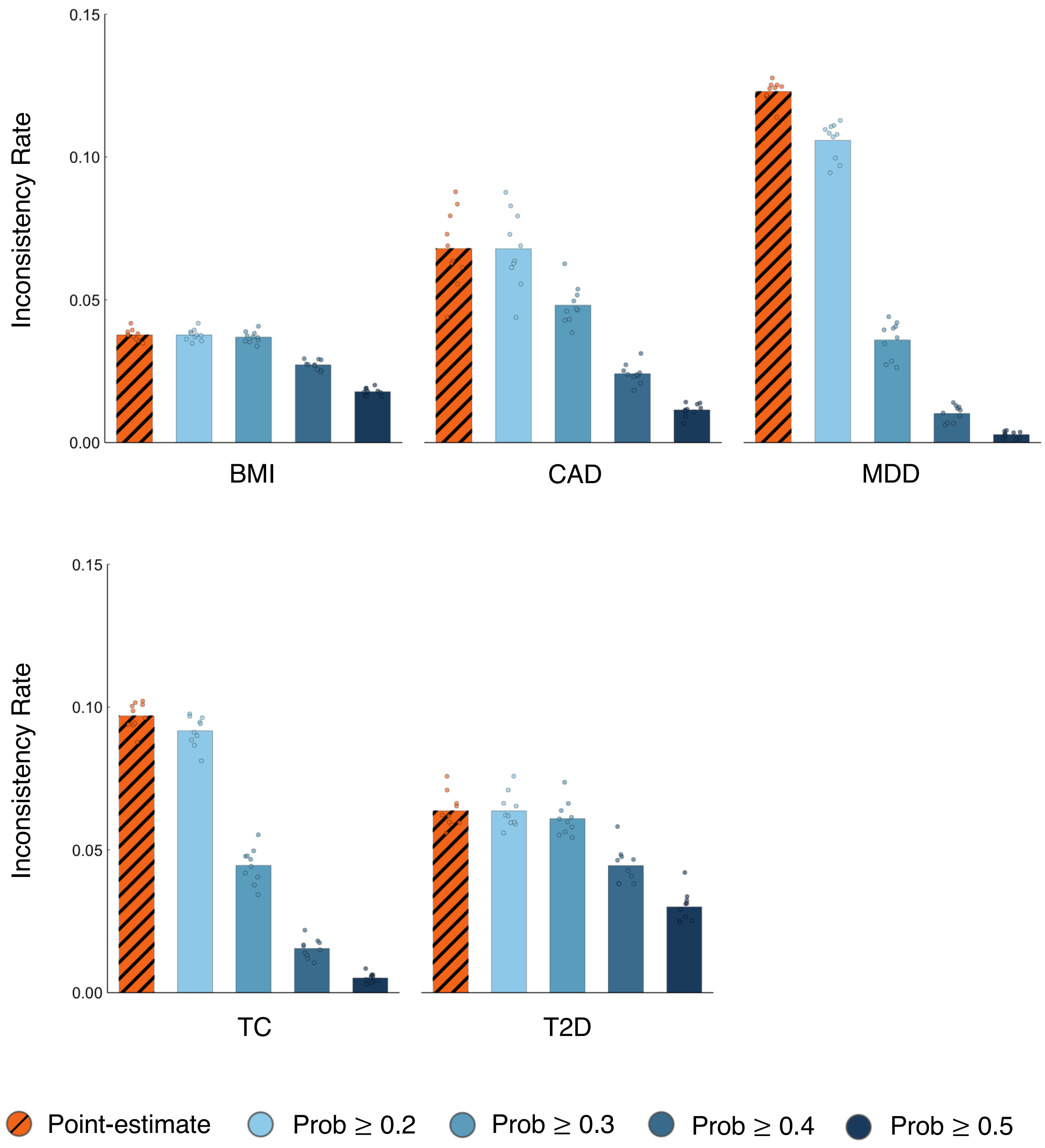


**Supplementary Figure 38:** Comparison of observed classification inconsistency rates for fully integrated PRSs of five representative complex traits and diseases under point-estimate thresholding versus uncertainty-aware probabilistic thresholding across a range of posterior probability cutoffs.

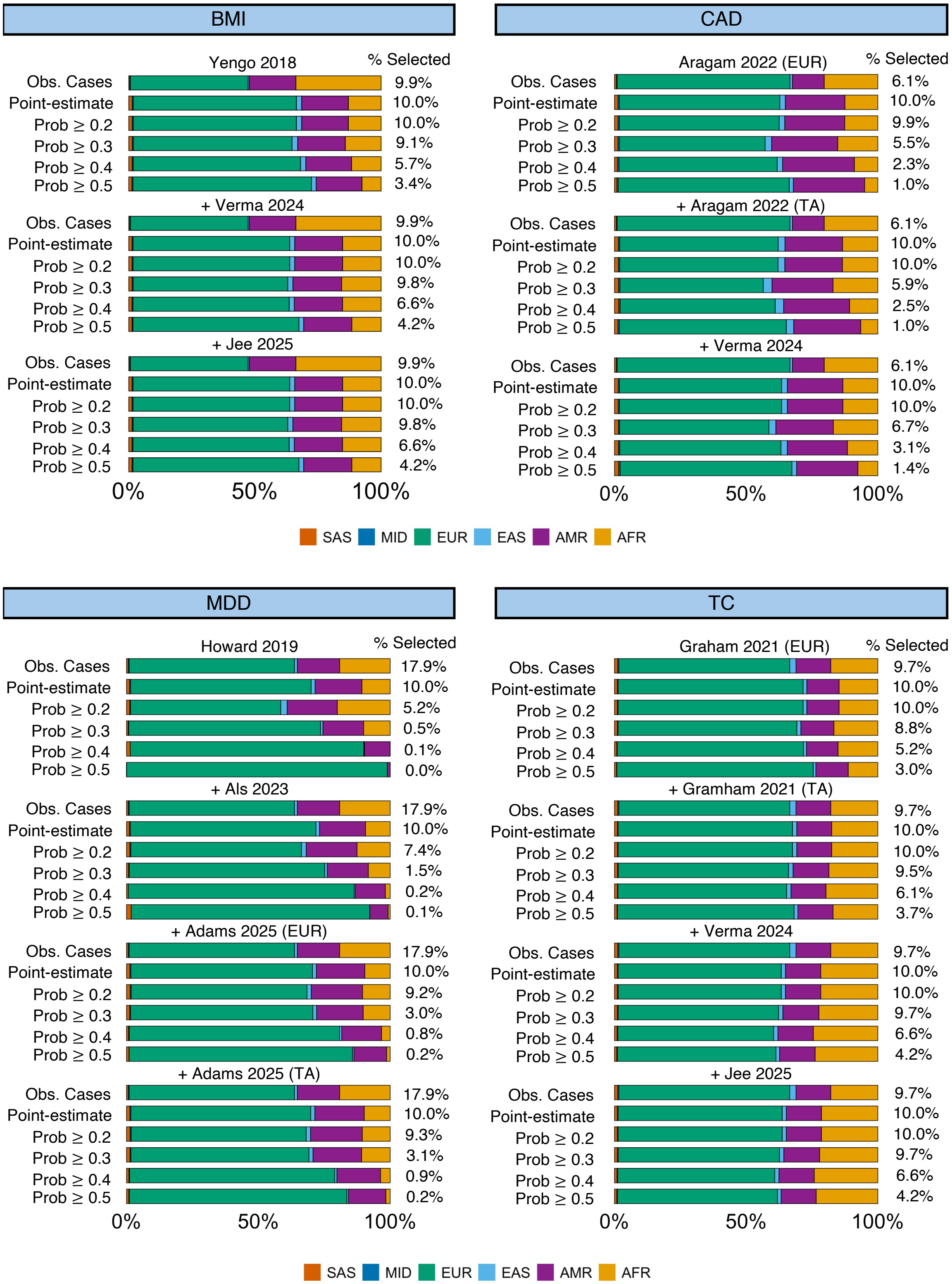


**Supplementary Figure 39:** Genetically inferred ancestry composition of high-risk individuals selected under point-estimate and uncertainty-aware probabilistic thresholding (across varying cutoffs), compared with the ancestry composition of observed cases for sequentially integrated PRSs of BMI, CAD, MDD, and TC.

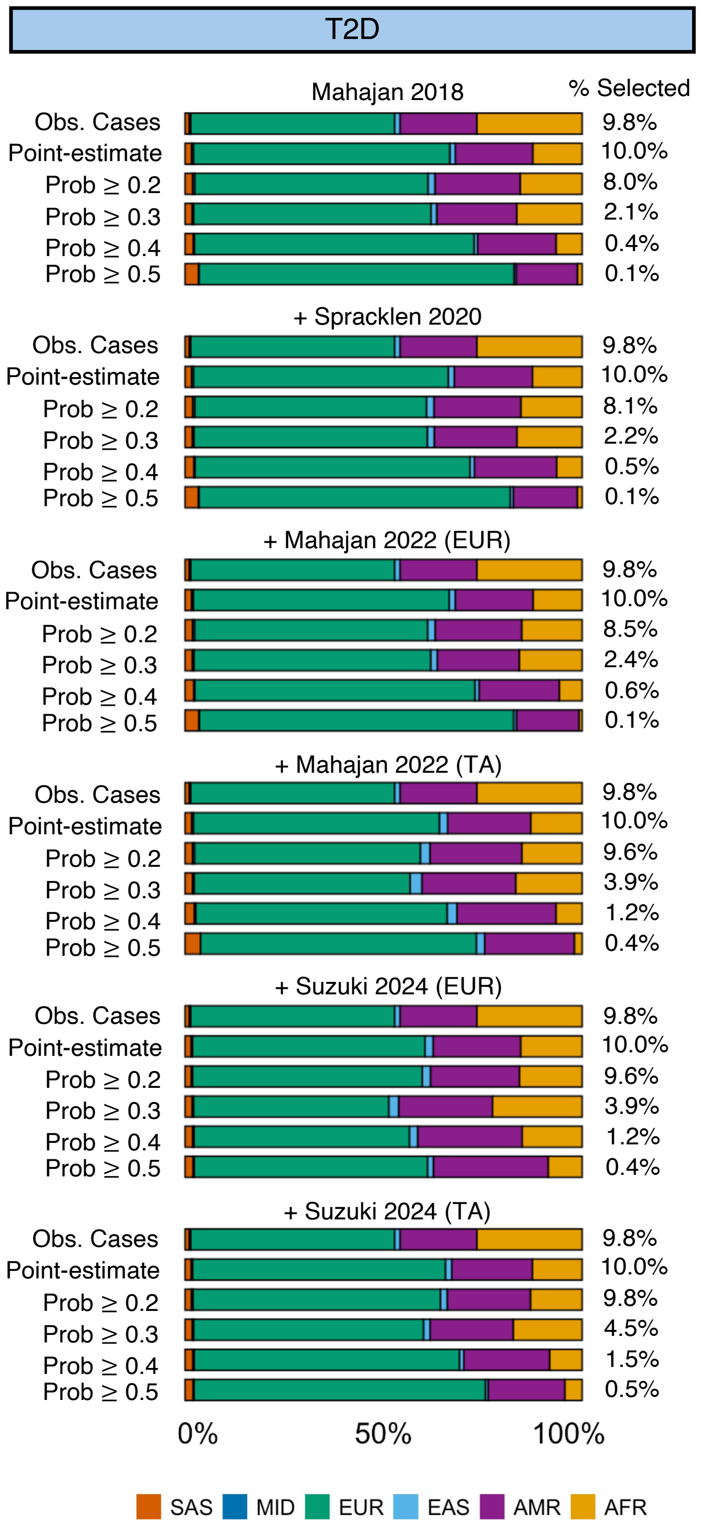


**Supplementary Figure 40:** Genetically inferred ancestry composition of high-risk individuals selected under point-estimate and uncertainty-aware probabilistic thresholding (across varying cutoffs), compared with the ancestry composition of observed cases for sequentially integrated PRSs of T2D.

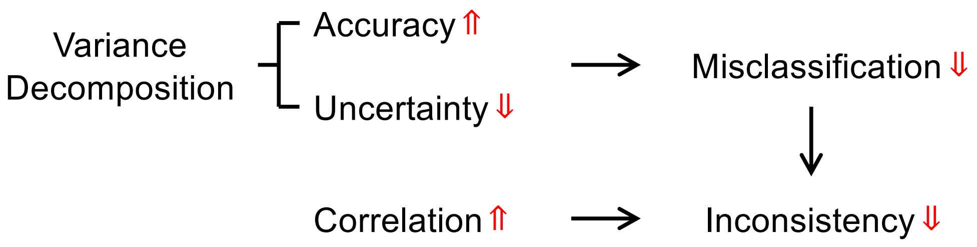


**Supplementary Figure 41:** A summary of the relationships linking PRS accuracy, uncertainty, correlation, misclassification, and inconsistency. For calibrated PRSs, accuracy and uncertainty are complementary components of the total genetic variance. The probability of misclassifying high-risk individuals decreases monotonically as PRS accuracy increases or uncertainty decreases. The probability of (with-truth) classification inconsistency across multiple PRSs decreases monotonically with their accuracies and pairwise correlations.
